# How well does societal mobility restriction help control the COVID-19 pandemic? Evidence from real-time evaluation

**DOI:** 10.1101/2020.10.29.20222414

**Authors:** Juhwan Oh, Hwa-Young Lee, Khuong Quynh Long, Jeffrey F Markuns, Chris Bullen, Osvaldo Enrique Artaza Barrios, Seung-sik Hwang, Young Sahng Suh, Judith McCool, S. Patrick Kachur, Chang-Chung Chan, Soonman Kwon, Naoki Kondo, Hoang Van Minh, J. Robin Moon, Mikael Rostila, Ole F. Norheim, Myoungsoon You, Mellissa Withers, Li Mu, Eun-Jeung Lee, Caroline Benski, Soo Kyung Park, Eun-Woo Nam, LLM Katie Gottschalk, Matthew M. Kavanagh, Tran Thi Giang Huong, Jong-Koo Lee, S.V. Subramanian, Lawrence O. Gostin, Martin McKee

## Abstract

**Objectives:** To determine the impact of restrictions on mobility on reducing transmission of COVID-19.

**Design:** Daily incidence rates lagged by 14 days were regressed on mobility changes using LOESS regression and logit regression between the day of the 100th case in each country to August 31, 2020.

**Setting:** 34 OECD countries plus Singapore and Taiwan.

**Participants:** Google mobility data were obtained from people who turned on mobile device-based global positioning system (GPS) and agreed to share their anonymized position information with Google.

**Interventions:** We examined the association of COVID-19 incidence rates with mobility changes, defined as changes in categories of domestic location, against a pre-pandemic baseline, using country-specific daily incidence data on newly confirmed COVID-19 cases and mobility data.

**Results:** In two thirds of examined countries, reductions of up to 40% in commuting mobility (to workplaces, transit stations, retailers, and recreation) were associated with decreased COVID-19 incidence, more so early in the pandemic. However, these decreases plateaued as mobility remained low or decreased further. We found smaller or negligible associations between mobility restriction and incidence rates in the late phase in most countries.

**Conclusion:** Mild to moderate degrees of mobility restriction in most countries were associated with reduced incidence rates of COVID-19 that appear to attenuate over time, while some countries exhibited no effect of such restrictions. More detailed research is needed to precisely understand the benefits and limitations of mobility restrictions as part of the public health response to the COVID-19 pandemic.

**WHAT IS ALREADY KNOWN ON THIS TOPIC:** Since SARS-CoV-2 became a pandemic, restrictions on mobility such as limitations on travel and closure of offices, restaurants, and shops have been imposed in an unprecedented way in both scale and scope to prevent the spread of COVID-19 in the absence of effective treatment options or a vaccine. Although mobility restriction has also brought about tremendous costs such as negative economic growth and other collateral impacts on health such as increased morbidity and mortality from lack of access to other essential health services, little evidence exists on the effectiveness of mobility restriction for the prevention of disease transmission. A search of PUBMED and Google Scholar for publications on this topic through Sep 20, 2020 revealed that most of the evidence on the effectiveness of physical distancing comes from mathematical modeling studies using a variety of assumptions. One study investigated only the combined effect of several interventions, including physical distancing, among SARS-CoV-2 infected patients.

**WHAT THIS STUDY ADDS:** This is the first study to investigate the association between change in mobility and incidence of COVID-19 globally using real-time measures of mobility at the population level. For this, we used Google Global Mobility data and the daily incidence of COVID-19 for 36 countries from the day of 100^th^ case detection through August 31, 2020. Our findings from LOESS regression show that in two-thirds of countries, reductions of up to 40% in commuting mobility were associated with decreased COVID-19 incidence, more so early in the pandemic. This decrease, however, plateaued as mobility decreased further. We found that associations between mobility restriction and incidence became smaller or negligible in the late phase of the pandemic in most countries. The reduced incidence rate of COVID-19 cases with a mild to moderate degree of mobility restriction in most countries suggests some value to limited mobility restriction in early phases of epidemic mitigation. The lack of impact in some others, however, suggests further research is needed to confirm these findings and determine the distinguishing factors for when mobility restrictions are helpful in decreasing viral transmission. Governments should carefully consider the level and period of mobility restriction necessary to achieve the desired benefits and minimize harm.

## INTRODUCTION

One of the most widely implemented policy response to the novel coronavirus (SARS-CoV-2) pandemic has been the imposition of restrictions on mobility.^1^ These restrictions have included both incentives, encouraging working from home, supported by a wide range of online activities such as meetings, lessons, and shopping, and sanctions, such as stay at home orders, restrictions on travel, and closure of shops, offices, and public transport.^2-5^ The measures constitute a major component of efforts to control the COVID-19 pandemic. Compared to previous epidemic responses, they are unprecedented in both scale and scope.^6^

The rationale underpinning these public health measures is that restricting normal activities decreases the number, duration, and proximity of interpersonal contacts and thus the potential for viral transmission. Transmission simulations using complex mathematical modelling have built on past experience such as the 1918 influenza epidemic,^7^ as well as assumptions about the contemporary scale and nature of contact in populations.^8^ However, the initial models were not always founded on empirical evidence from behavioral scientists on the feasibility or sustainability of mass social and behavior change in contemporary society. While reductions in interpersonal contact and increases in physical distancing are known to decrease respiratory infection spread,^9^ the paucity of recent examples of large-scale restrictions on mobility has limited the scope for research on their impact on transmission. Where restrictions have been imposed, as with Ebola, they have involved diseases with a different mode of transmission. Nonetheless, the rapidity of progression of this pandemic has forced many governments into trialing various approaches to containment with limited evidence of effectiveness.^10^

More conventional public health prevention measures (such as quarantine of contacts, isolation of infected individuals and contact tracing) and control measures in health systems (such as patient flow segregation, negative pressure ventilation, and use of personal protective equipment),^11-14^ have been applied widely to control the epidemic in many countries as part of a portfolio of policy responses. However, mobility restriction as a new large-scale mass behavioral and social prescription has incurred considerable costs.^15 16^ Estimates suggest global GDP growth has fallen by as much as 10%,^17^ at least in part due to mobility restriction policies. Although views differ, not least because of the lack of information of what would happen if the disease was unchecked and the emerging evidence of persisting disability in survivors, some have argued that this is greater than would be accounted for by the economic impact of direct illness and deaths from COVID-19.^18 19^ More recently, a study in the U.S. looking at human mobility patterns suggested that mobility restrictions were associated with reductions in viral transmission,^20^ while another recent analysis of GDP and death rates suggested poorer pandemic control tends to be associated with greater economic loss.^21^

To inform decisions on large scale restrictions of mobility, there is an urgent need to assess their effectiveness in limiting pandemic spread. To this end, we examined the association of mobility with COVID-19 incidence in Organization of Economic Cooperation and Development (OECD) countries and equivalent economies such as Singapore and Taiwan.

## METHODS

### Study population and data

The study population consisted of the populations of 36 countries (all 34 OECD countries plus Singapore and Taiwan). Two sets of publicly available data were pooled into one dataset: country-specific newly confirmed COVID-19 cases per day retrieved from the dataset curated by *Our World in Data* (https://ourworldindata.org/coronavirus),^22^ and country-specific mobility change data obtained from the *Google Global Mobility Data Source*.^23 24^

### Variables

#### Independent variables

We used country-specific mobility data, collected from mobile device-based global positioning system (GPS) information from people who agreed to share their anonymized position information with Google, starting from the date the 100^th^ case was detected in a country. The average mobility around specific categories of locations during the reference period (between January 3 and February 6, 2020, a period before COVID-19 was declared a global pandemic) was applied as a baseline mobility value for each country to calculate the percent mobility change on each day during the study interval for each country. The data were categorized by Google into 6 different types of locations visited (workplaces, transit stations, retailer and recreational places, residential areas, groceries and pharmacies, and parks). To reduce multicollinearity, three highly correlated variables (workplace mobility, transit station mobility, and retailer and recreation mobility) were averaged into a combined latent variable (“commuting mobility”) as the main mobility variable for this study, as this is the activity that is most likely to bring large groups on people together. The correlation analysis of different types of mobility is shown in Supplementary Fig 1.

#### Outcome variables

The outcome variable chosen was the daily national incidence rate of new COVID-19 cases (daily new cases per accumulated cases over the prior 14 days), intended to reflect the total number of local active infections on that day, starting from the 14^th^ day from the date the 100^th^ case was detected in each country through August 31, 2020.

### Analysis

First, daily incidence rates of COVID-19 and mobility changes during the study period were described. Second, the daily national COVID-19 incidence rates were compared to changes in mobility. The analysis sought any country-specific associations between COVID-19 daily incidence rate and the percentage change in mobility for each of the 36 countries with a 14-day lag using LOESS regression, a nonparametric technique that uses local weighted regression to fit a smooth curve through points in a scatter plot.^25^ Based on the range and frequency of commuting mobility values observed across countries, we grouped values into mild (up to 20%), moderate (21-40%) and extensive (greater than 40%) degrees of mobility change, when describes various degree of associations per each mobility change. Logit-transformed incidence rates were regressed to mobility changes to compare the country-specific association using a single regression coefficient per each country. We performed additional analysis by designating an early phase versus late phase using the median date (between the day of the 100^th^ case and the last day of study period–August 31) for each country; and by using different mobility location categories (i.e. residential areas, parks, or groceries and pharmacies) beyond the main analysis focused on commuting mobility. The unit of analysis was country level. All analyses used R (version 4.0.0, R Foundation for Statistical Computing, Vienna, Austria).

#### Sensitivity Analysis

We performed sensitivity analyses by varying the lag interval (7 or 21 days) between the change in mobility and incidence of infection to apply various incubation period possibilities.

## PATIENT AND PUBLIC INVOLVEMENT

Patients or the public were not involved in any phase of the design, conduct, reporting, or dissemination plans of our research other than when they decided to share their data with Google. All data used were obtained from existing public sources. We plan to share these findings on social media, Twitter, and blogs.

## RESULTS

There were 5,766 observations with data on daily incidence of COVID-19 infections and mobility changes, from 36 countries, starting with the 100^th^ case in each country, which ranged from February 21 (South Korea) to March 23 (New Zealand), and continuing until August 31, 2020. The changes in commuting mobility were pronounced, from a 91.8% decrease (Spain on Apr 10) to a 60.8% increase (Greece on Aug 16). National daily incidence rate of COVID-19 cases was as high as 0.64 (New Zealand on Mar 10). There was a reduction in incidence in most countries as mobility decreased by up to 20% or 40% but, in most cases, then reached a plateau with little further change after additional reductions (Fig. 1 and 2). The degree of association varied, with some countries experiencing relatively small changes in mobility. The magnitude of the association tended to be larger in the earlier period, as shown in Fig. 3 where the country- and phase-specific slopes of the curves (point estimation with 95% confidence interval) are shown using logit regression.

Of note, small increases in mobility in residential areas, seen throughout most of the study period in all countries, were associated with reductions in COVID-19 incidence, but larger increases had no additional effect in the early phase, and there were no associations in any ranges in the late phase (Fig. 4, Fig S2). In the early phase, increased mobility in parks was associated with increased incidence in 5 countries (US. Spain, Japan, Estonia, and Latvia) whereas it was not associated in other countries. In late phase, there was no association in most countries (Fig. 4 and Fig. S2). Mobility around groceries and pharmacies, generally viewed as essential visits, was typically associated with the highest incidence of infection at a time when mobility was close to baseline, during the early phase (Fig. S3). In the late phase, however, this finding no longer held and there was no association between infection rate and mobility, regardless of the degree of mobility change. In fact, results from the late phase show little association between incidence and mobility changes in any locations in most countries, except some for a reduction in incidence with moderate restriction in commuting mobility in Ireland, Australia, Italy, Spain Estonia, and Hungary; and an increased incidence with increased mobility around parks in Estonia, Hungary, and Spain (Fig. 1, 2, 3 and 4; Fig. S3 and S4). The findings were mostly consistent in the sensitivity analysis, when we used alternative lag periods of 7 or 21 days in the model (Fig. S4).

## DISCUSSION

Overall, the expectation that reduced mobility would be associated with reduced incidence of COVID-19 cases was confirmed, with a minimum reduction of 20-40%. However, the decrease plateaued as mobility remained low or decreased further. Mobility restrictions were most effective in the early phase of the pandemic in most countries with a much smaller effect in the later phase.

This study has several limitations. First, Google data are only generated by Android users who have location services switched on. These individuals may be an atypical minority of the population in some countries, or there may be changes in the type of users over time, such as tourists (as in Greece), although in most cases numbers will be small. Second, reported incidence rates are influenced by availability of testing and quality of reporting. Third, the use of an ecological design introduces scope for confounding and imprecision: our data do not allow us to isolate the impact of mobility restrictions from the many other variables that intervene in a pandemic response, such as the degree of inter-household mixing, the ability to detect and rapidly control an outbreak, and behavioral characteristics, such as use of face coverings and adherence to physical distancing guidelines, themselves influenced by clarity of messaging and trust in official advice. A related limitation is that, in federal countries such as the United States, there may be substantial sub-national differences in implementation of these characteristics and in other policies. However, given the complex nature of these relationships, influenced by starting conditions, feedback loops, and non-linear relationships, the analytic challenges of disentangling these factors are formidable even if data were available.

There are a variety of possible explanations for our findings. Mandatory restrictions on mobility may reduce both the frequency and/or duration of interpersonal interactions and, by reinforcing official advice, the nature of those interactions, with greater distancing and hygiene precautions. As restrictions are lifted, protective behaviors such as distancing may have become normalized, mitigating the effects of greater mobility. When this is coupled with improved contract-tracing, quarantine, and support for isolation, the relative benefits of mobility restrictions may decrease as the population adopts additional risk mitigation measures, allowing a return to greater mobility (adapting to the “new normal”). These behavioral changes may also be enhanced through social learning of these practices promoted by trusted and authoritative public health messaging. While this finding supports the role of mobility restrictions as a critical strategy early in a pandemic or when a country has yet to implement more comprehensive and meticulous mitigation strategies, it suggests that rapid scale-up of a suite of other coordinated mitigation strategies simultaneously as part of a strategy to drive transmission as low as possible may maximize health gains while enabling a gradual reopening of the economy.^26-31^

These findings are consistent with other research suggesting benefits of early mobility restrictions.^20^; for example, an earlier study of government-imposed physical distancing interventions demonstrated an association with larger reductions in COVID-19 incidence, although this did not directly assess the degree of mobility reduction nor the impact of timing of the interventions.^32^ Modeling in China using mobile phone data in the initial phase of pandemic provides further support for this view.^33^

Our study uses mobile phone tracking to analyze real-time changes in mobility. In at least some jurisdictions the public seems to have pre-empted official instructions, reacting to emerging data (28)^34^. One study in the United States, where implementation of other public health mitigation strategies has varied, found a strong correlation between decreased mobility patterns and lower COVID-19 case growth rates using similar phone tracking data.^34^

Our analysis focused on changes in commuting mobility. However, this is only one way in which people mix and spread infections, with inter-household spread important in some settings.^35^ Although we did find some associations between incidence and, for example, mobility in residential areas, these are difficult to interpret and investigation of these phenomena will require other data sources, such as apps that record close contacts. Finally, our finding of little difference using different lag times (7, 14 or 21 days) is consistent with prior research suggesting a typical lag between interventions and changes in infection rate of around 14 days.^34 36^

While additional work is needed to elaborate on these findings, they may be useful to policymakers as they adapt responses to the pandemic over time and, in particular, point to the importance of including mobility data when developing comprehensive strategies.

Severe restrictions on mobility, or lockdowns, are blunt but necessary instruments at the beginning of a pandemic. If sufficiently stringent to get the R number below 1 they will drive down the level of infection but at considerable human and economic cost. The time bought must be used to establish effective testing and tracing systems. If both are done, and other non-pharmacological interventions such as wearing of face coverings and social distancing are adhered to, cit should be possible to slowly open up society in the knowledge that local resurgences can be contained. However, if these systems fail and cases rise rapidly, they may be required again, but this should be seen as a last resort and evidence of policy failure.

## CONCLUSION

Our analysis extends the understanding of the complex dynamics at play when mobility is restricted at a population level in response to a pandemic caused by a respiratory virus. Societal mobility restrictions appear to have reduced COVID-19 spread in many countries, particularly in the early phase of the pandemic, but in the late phase, when other measures have been adopted, the magnitude of impact is attenuated. It is critical for policymakers to consider the effectiveness of mobility restriction to COVID 19 response and the economic impacts imposed on their society as this pandemic is far from an end and the society may need to adjust to the “new normal” way of life. For this, additional evidence, including the relationship with other non-pharmacological interventions, is needed to fully understand the role of mass restrictions on mobility in containing COVID-19 and future infectious diseases with a similar mode of transmission. As the pandemic progresses, governments must develop strategies that bear down on the amount of circulating virus and allow rapid responses to outbreaks. The pandemic has brought enormous changes to working and living, some of which will likely persist even if and when a vaccine is made available. Surveillance that goes beyond incidence of infection, to include risk factors such as mobility, can only improve our ability to develop effective public health responses.

## Data Availability

All data used are publicly available

https://www.google.com/covid19/mobility/

https://ourworldindata.org/coronavirus

## Contributors

JO conceptualized the study. JFM, YS, and KL performed data curation. KL, JO, HWL and YS performed formal analyses. JO and SVS supervised methodology. SVS validated the analysis. JO and HWL drafted the manuscript. All authors interpreted the results and reviewed and revised the paper. HYL, SVS,MM, and LG supervised all the work.

## Ethical approval

not applicable

## Data sharing

All data used are publicly available

---

Legend of figures

Figure 1

Association between new daily incidence rates of COVID-19 and mobility changes for 36 countries, by pandemic phase.

Footnote 1. The mobility change measurement period was from the day of the 100th case in each country through August 31, 2020.

Footnote 2. Pandemic phase was defined for each country by the median of the date when the 100^th^ case was detected to the end of the study period: early phase for the period before the median date and late phase for the period after the median date.

Figure 2

Association between new daily incidence rates of COVID-19 and mobility changes in each of 36 countries, by measurement period.

Figure 2A. Western Europe, North America, and Oceania Figure 2B. Asia, Eastern Europe, Latin America, and Caribbean

Footnote 1. The mobility change measurement period was from the day of the 100th case in each country through August 31, 2020.

Footnote 2. Pandemic phase was defined for each country by the median of the date when the 100^th^ case was detected to the end of the study period: early phase for the period before the median date and late phase for the period after the median date.

Figure 3

Forest plot showing unadjusted estimate for the association of COVID-19 incidence rate logit-transformed with mobility changes

Footnote 1. The mobility change measurement period was from the day of the 100th case in each country through August 31, 2020.

Footnote 2. Pandemic phase was defined for each country by the median of the date when the 100^th^ case was detected to the end of the study period: early phase for the period before the median date and late phase for the period after the median date.

Figure 4

Association between new daily incidence rates of COVID-19 and mobility changes for 36 countries, early and late phase, for parks and residential areas.

Footnote 1. Pandemic phase was defined for each country by the median of the date when the 100^th^ case was detected to the end of the study period: early phase for the period before the median date and late phase for the period after the median date.

Footnote 2. (1) Parks and (2) Residential areas.

Legend of Supplementary Figures: 1-4

Supplementary Figure 1

Correlation matrix between six categories of mobility locations: workplaces; transit stations; retailer and recreational places; residential areas; groceries and pharmacies; parks; and commuting (average of workplaces; transit stations; retailer and recreational places)

Footnote 1. The mobility change measurement period was from the day of the 100th case in each country through August 31, 2020.

Supplementary Figure 2

Association between new daily incidence rates of COVID-19 and mobility changes in 36 countries by early and late phase and other places (parks and residential areas) visited.

Supplementary Figure 2A. Western Europe, North America, and Oceania Supplementary Figure 2B. Asia, Eastern Europe, Latin America, and Caribbean

Footnote 1. The mobility change measurement period was from the day of the 100th case in each country through August 31, 2020.

Footnote 2. Pandemic phase was defined for each country by the median of the date when the 100^th^ case was detected to the end of the study period: early phase for the period before the median date and late phase for the period after the median date.

Supplementary Figure 3

Association between new daily incidence rates of COVID-19 and mobility changes in 36 countries, for grocery and pharmacy visits.

Footnote 1. The mobility change measurement period was from the day of the 100th case in each country through August 31, 2020.

Footnote 2. Pandemic phase was defined for each country by the median of the date when the 100^th^ case was detected to the end of the study period: early phase for the period before the median date and late phase for the period after the median date.

Supplementary Figure 4

Association between new daily incidence rates of COVID-19 and mobility changes for 36 countries based on alternative lag days

Footnote 1. The mobility change measurement period was from the day of the 100th case in each country through August 31, 2020.

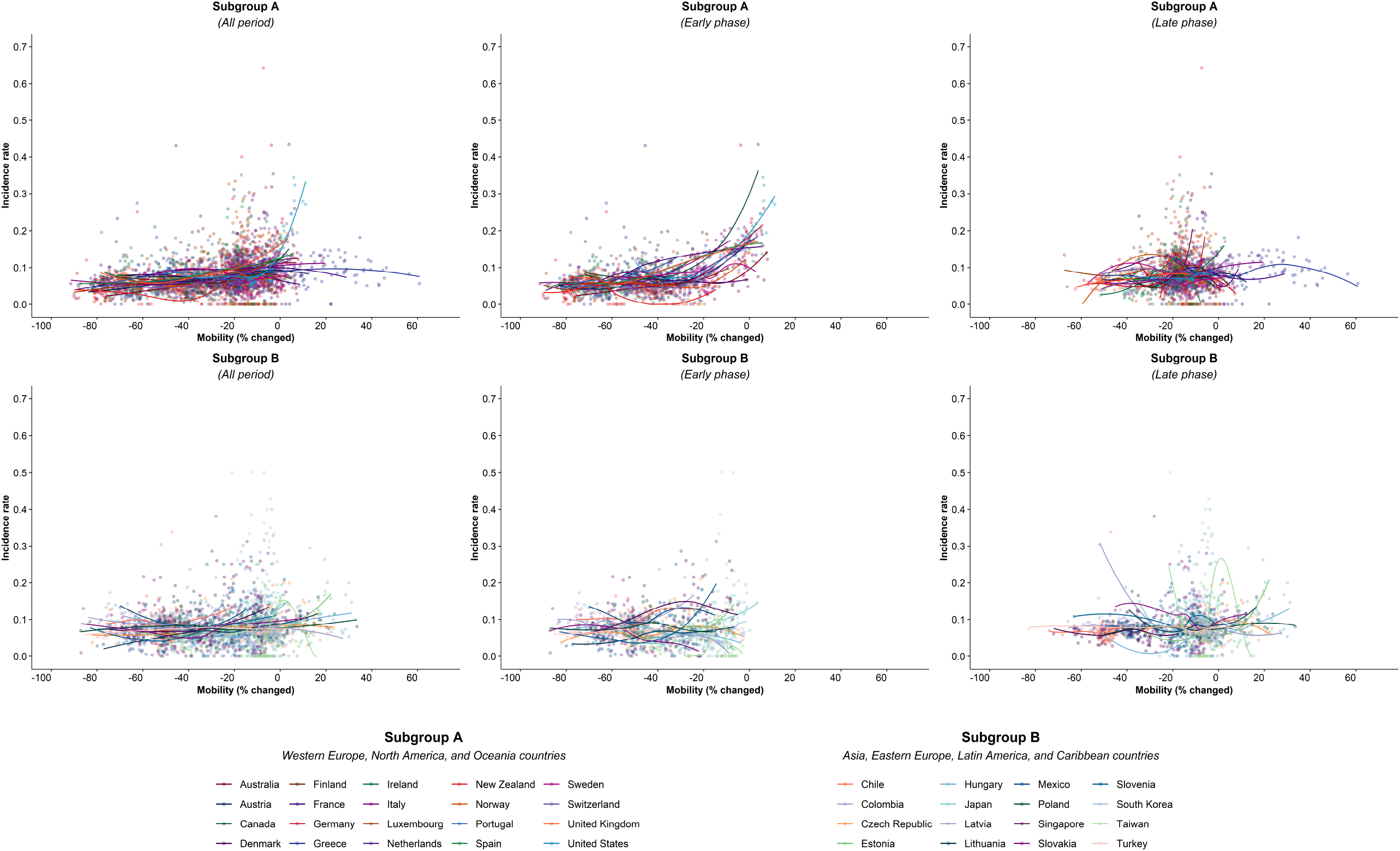

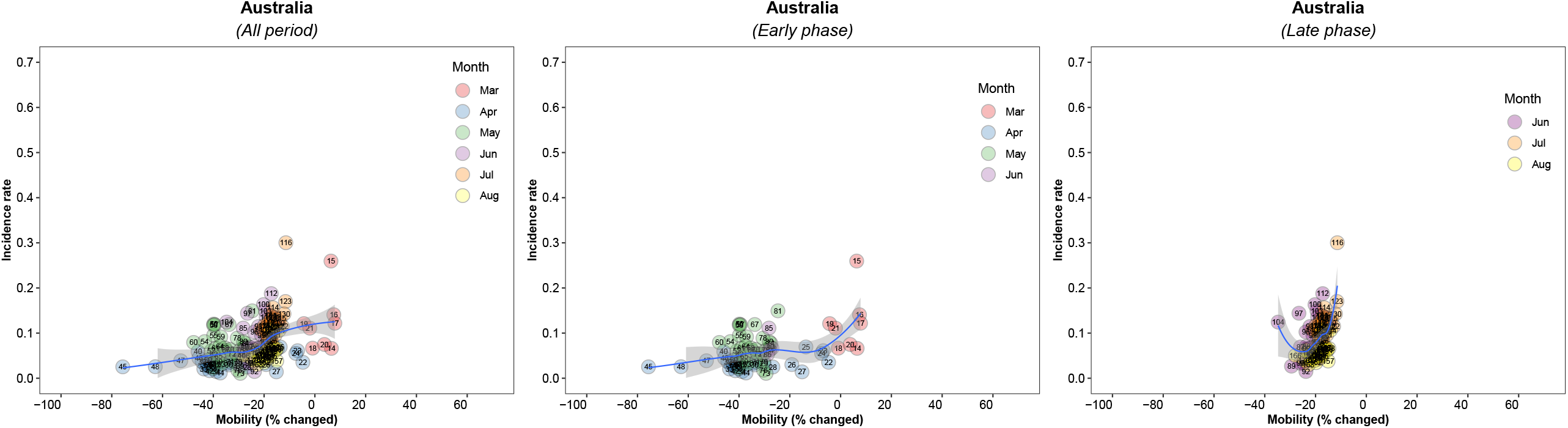

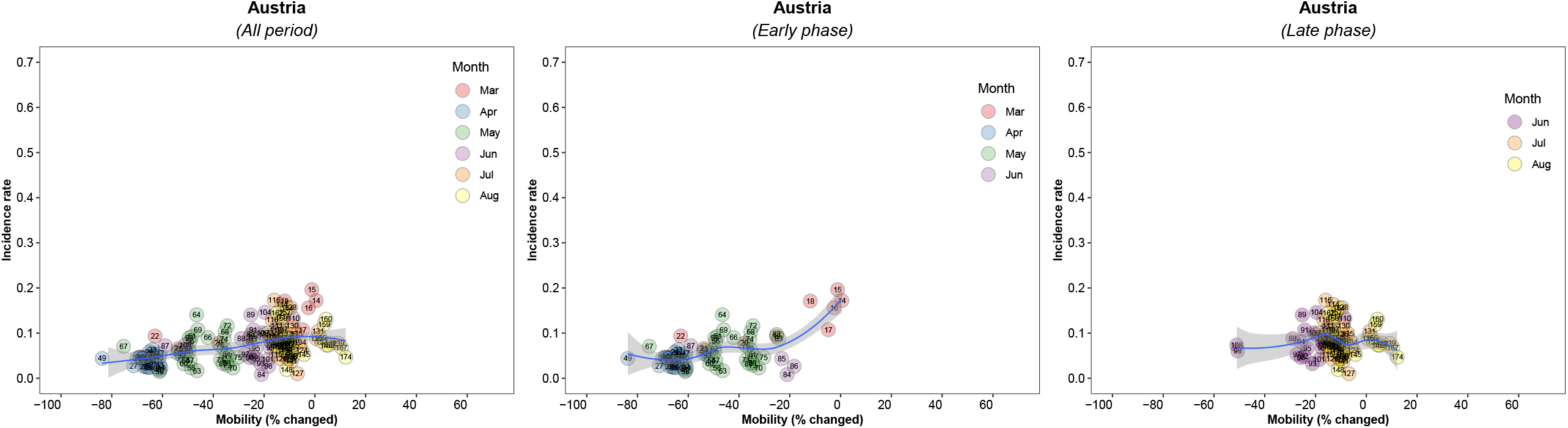

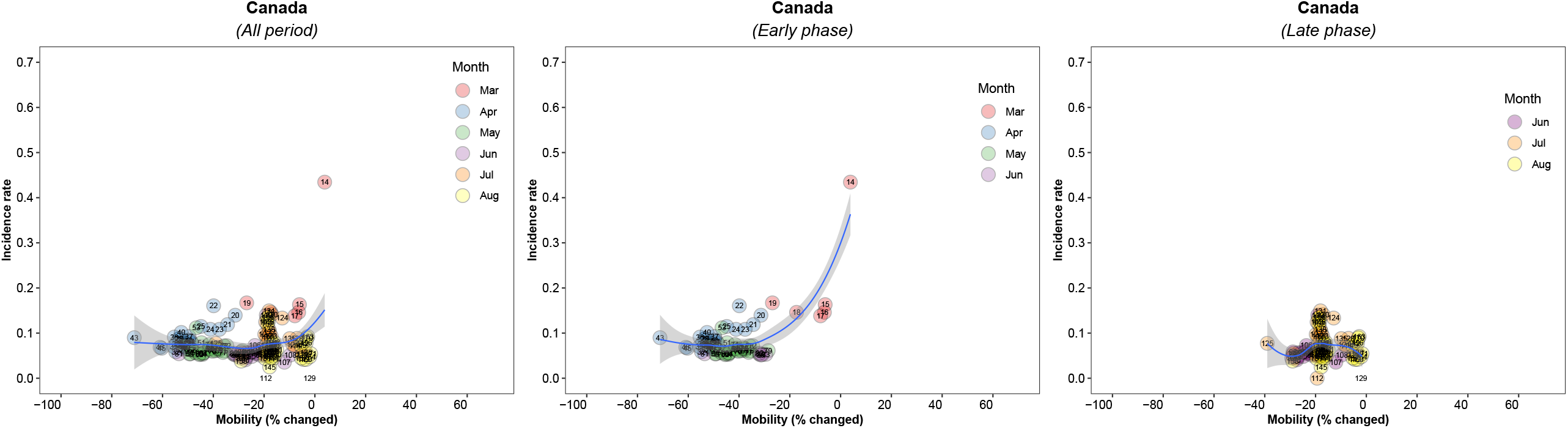

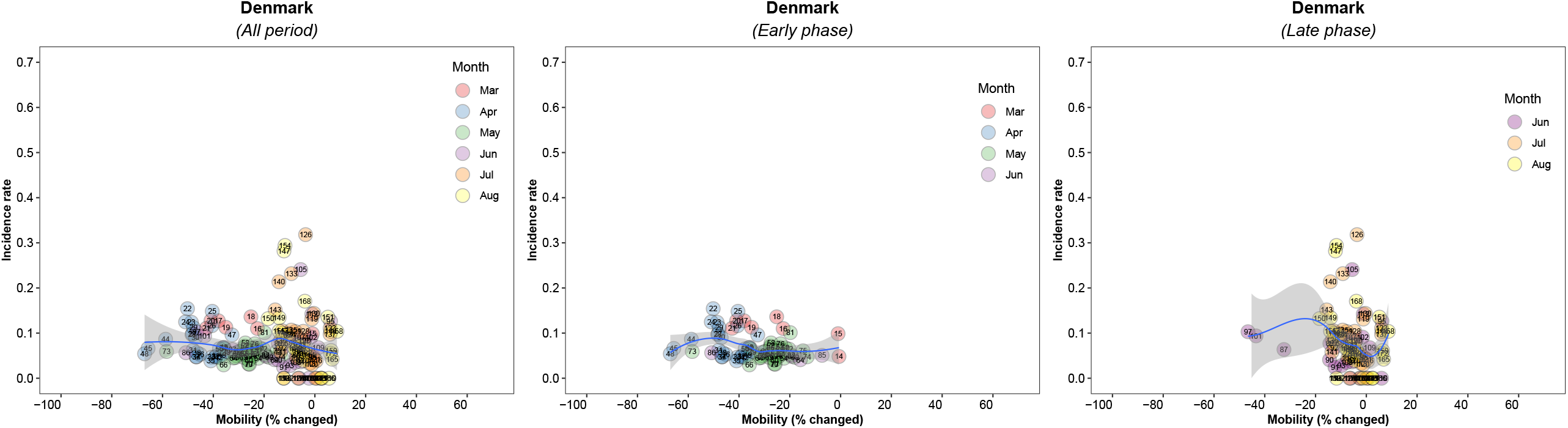

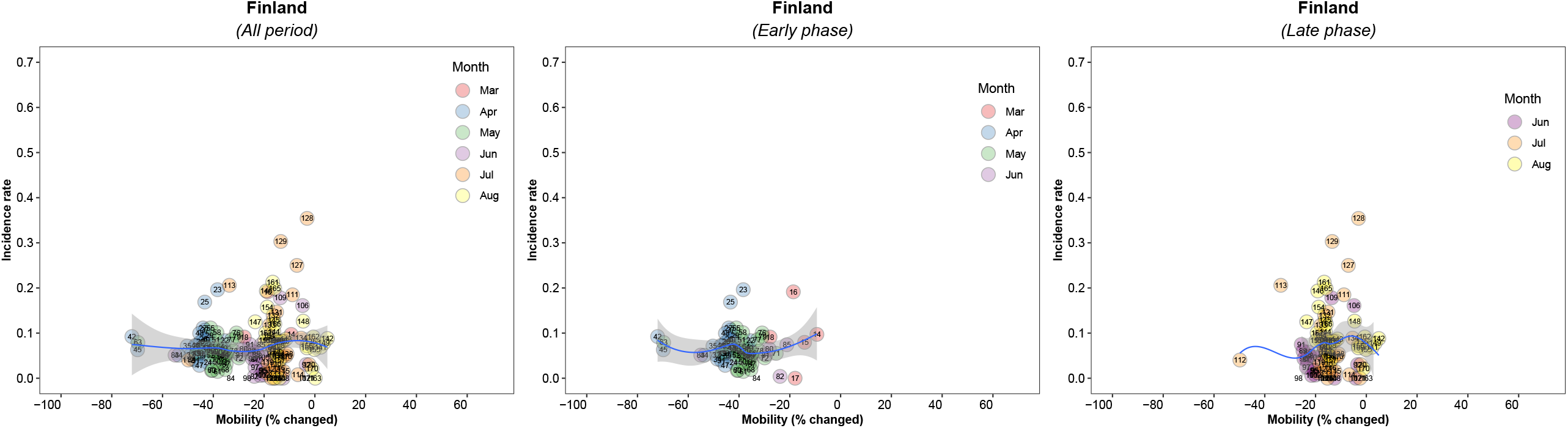

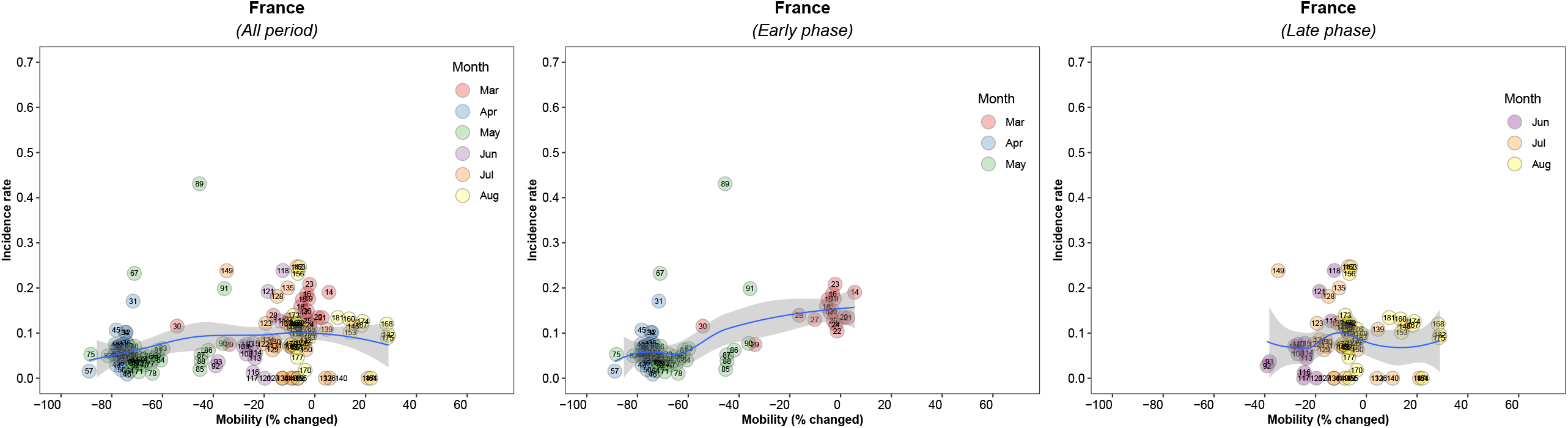

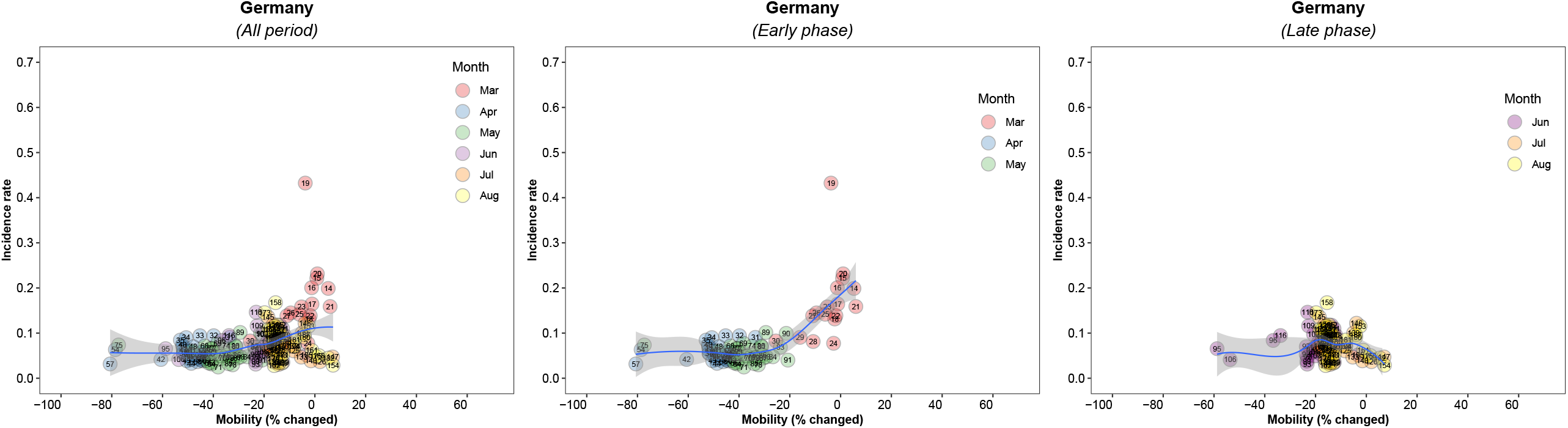

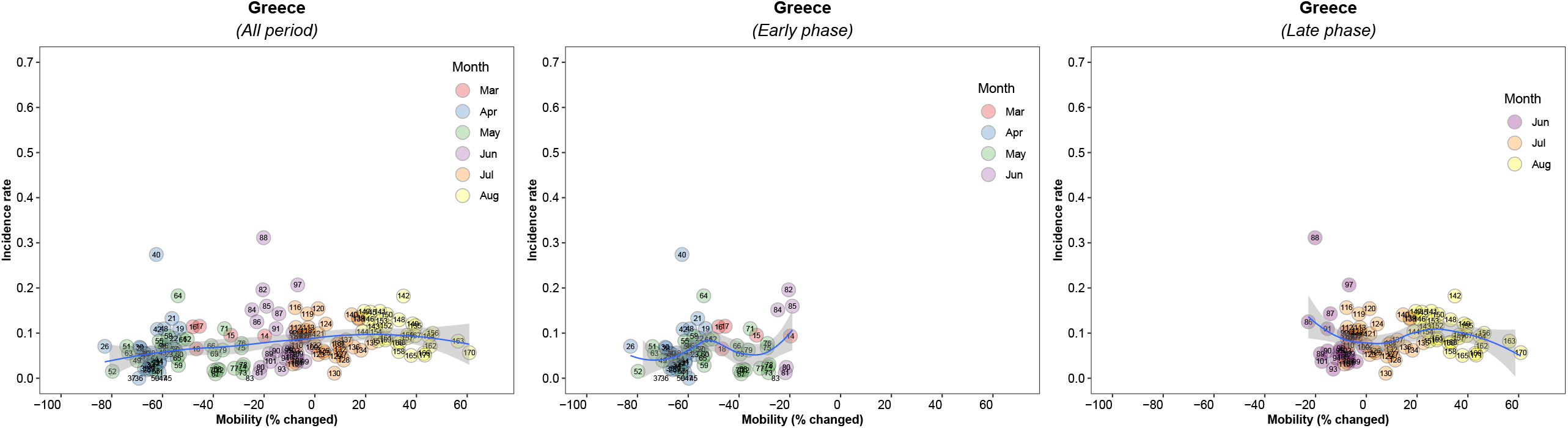

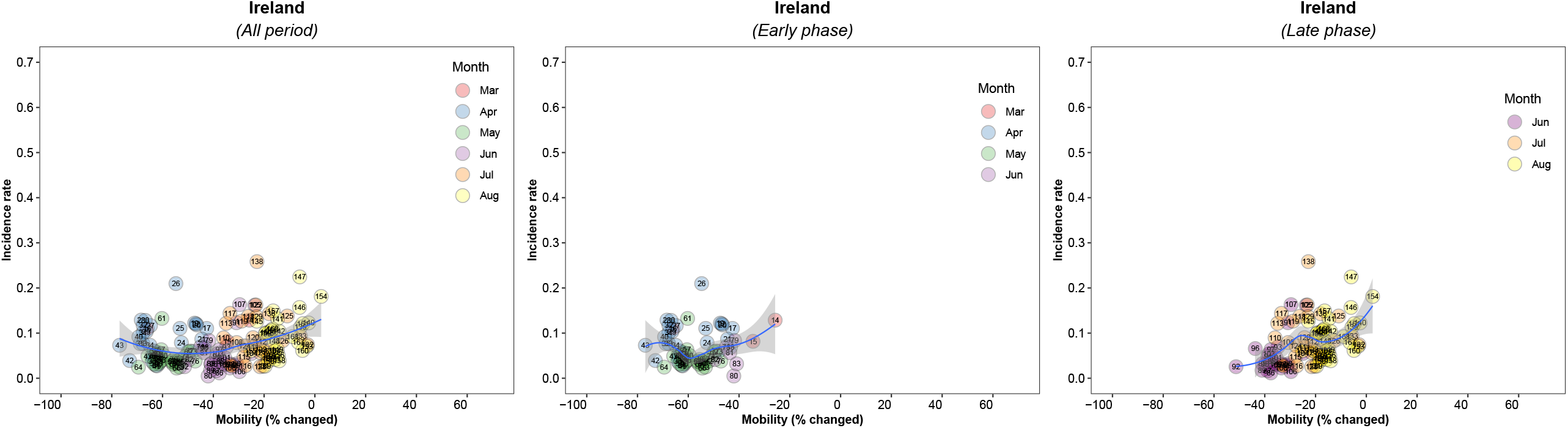

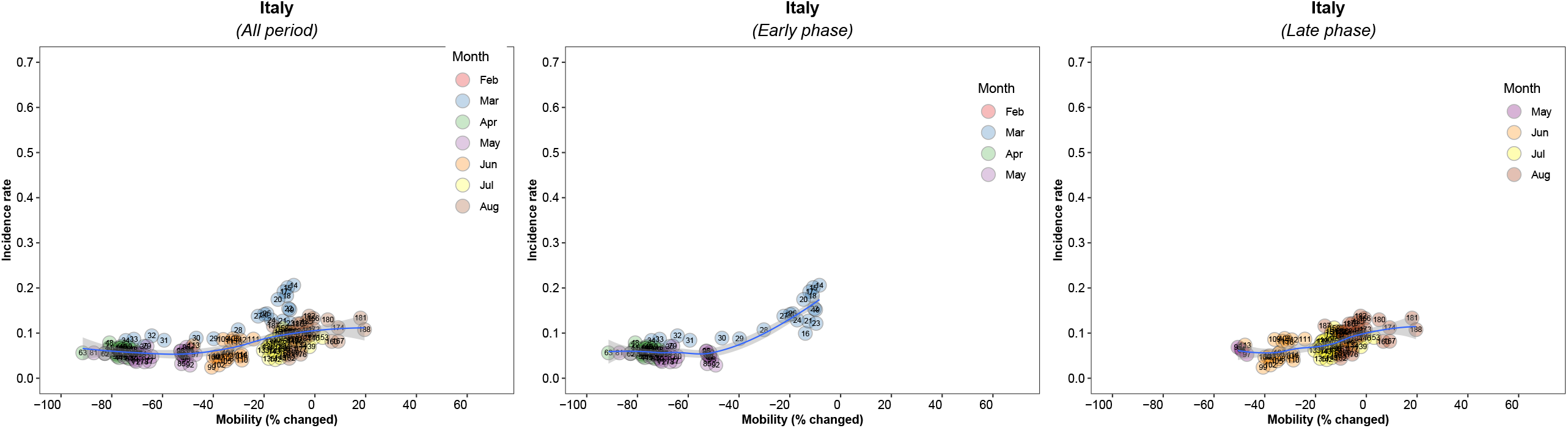

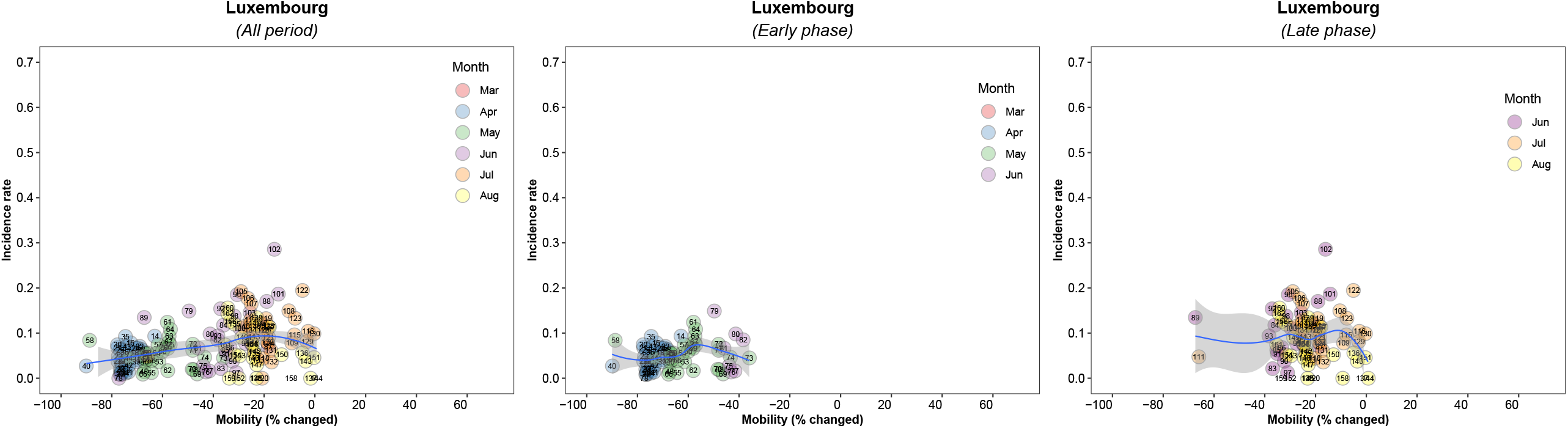

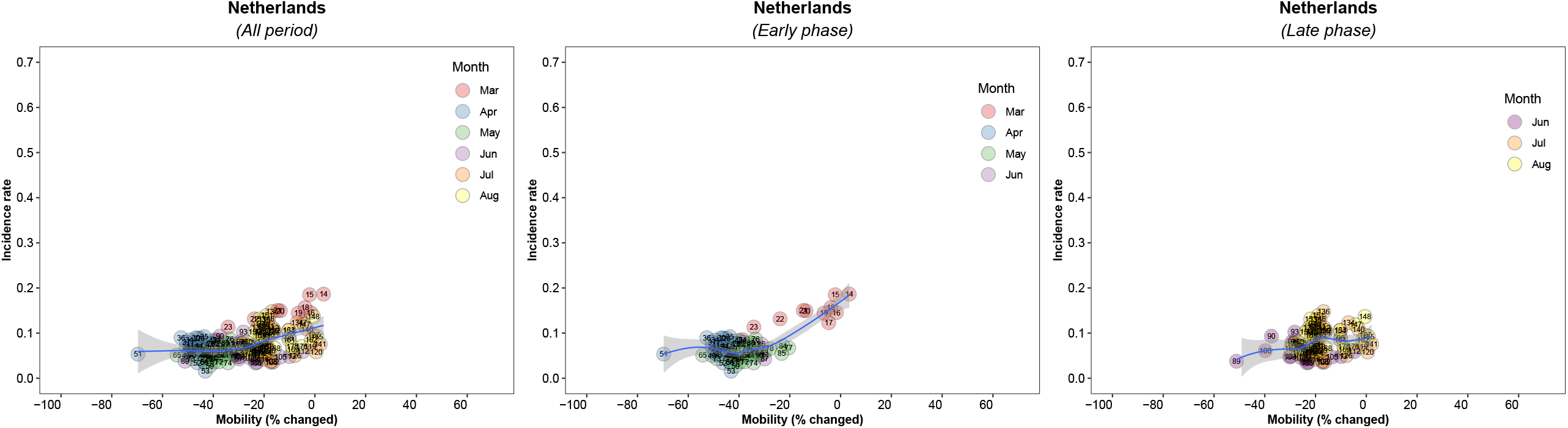

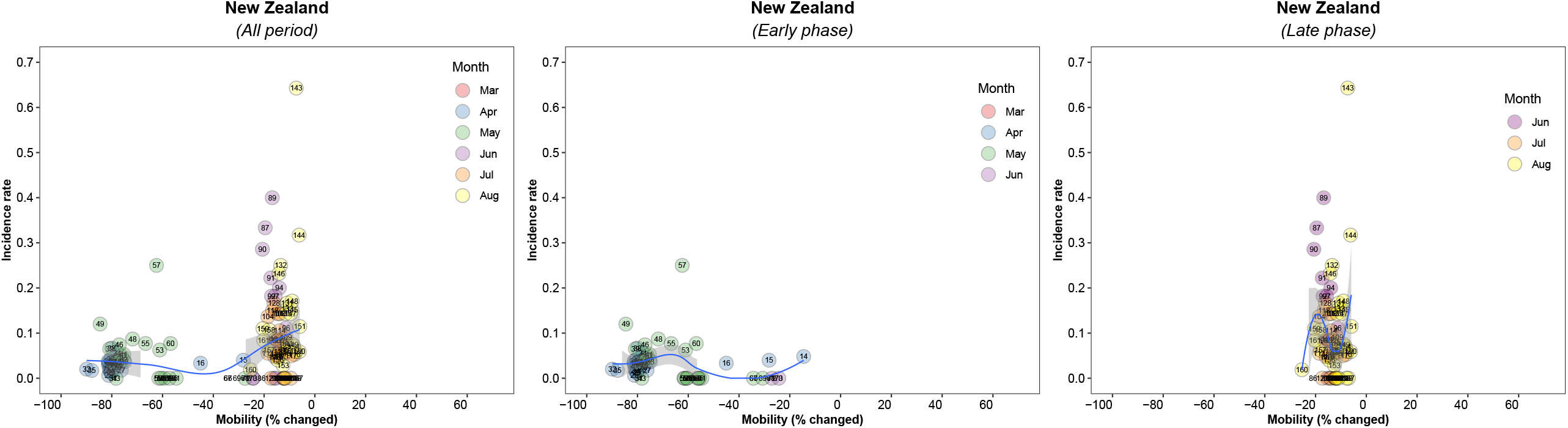

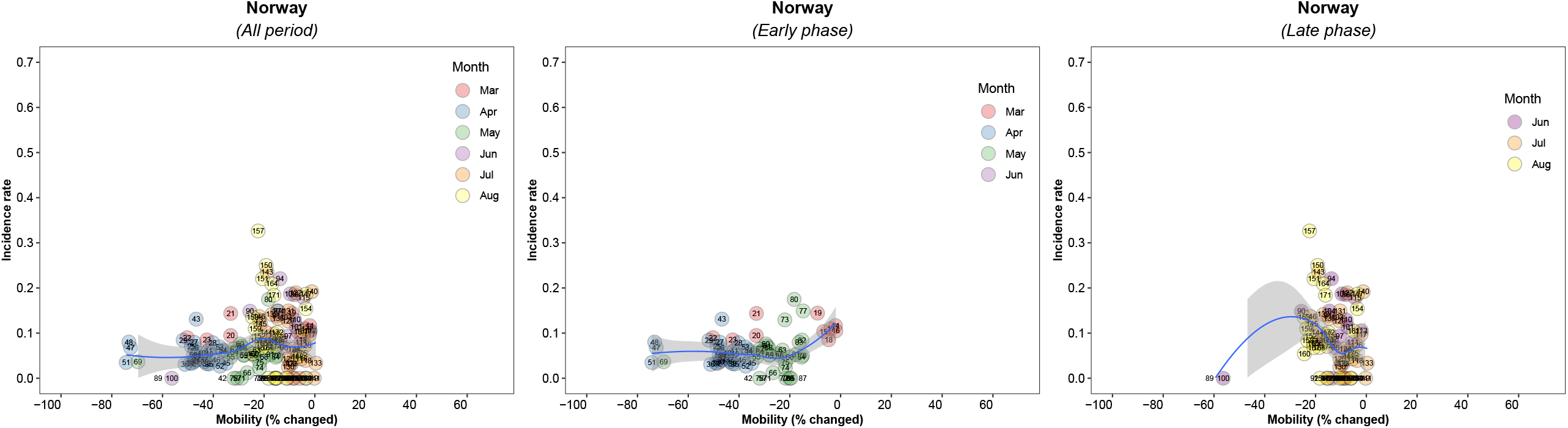

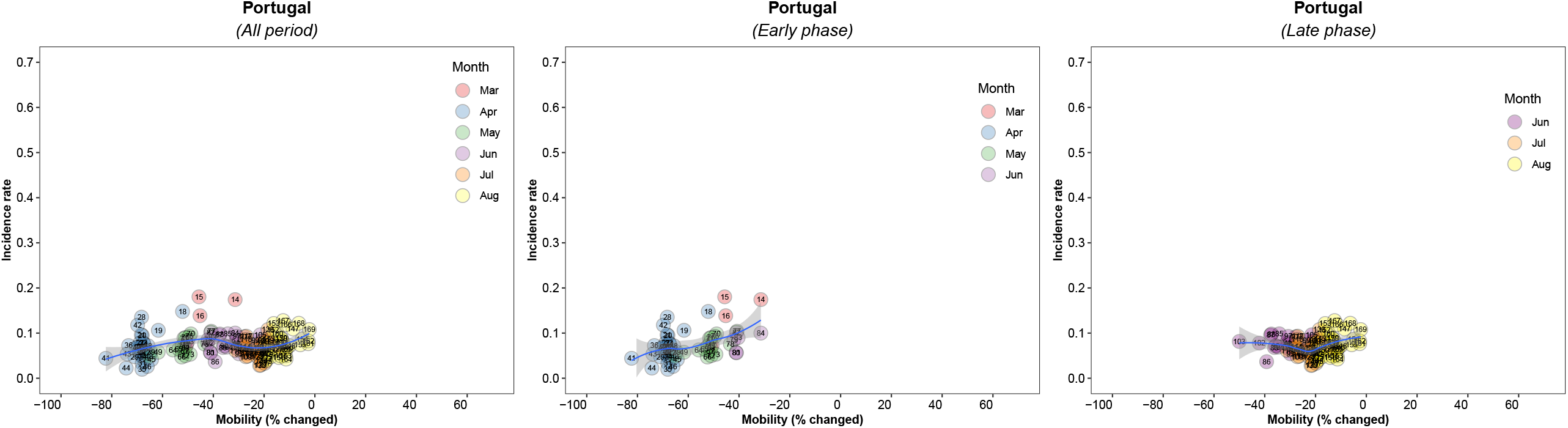

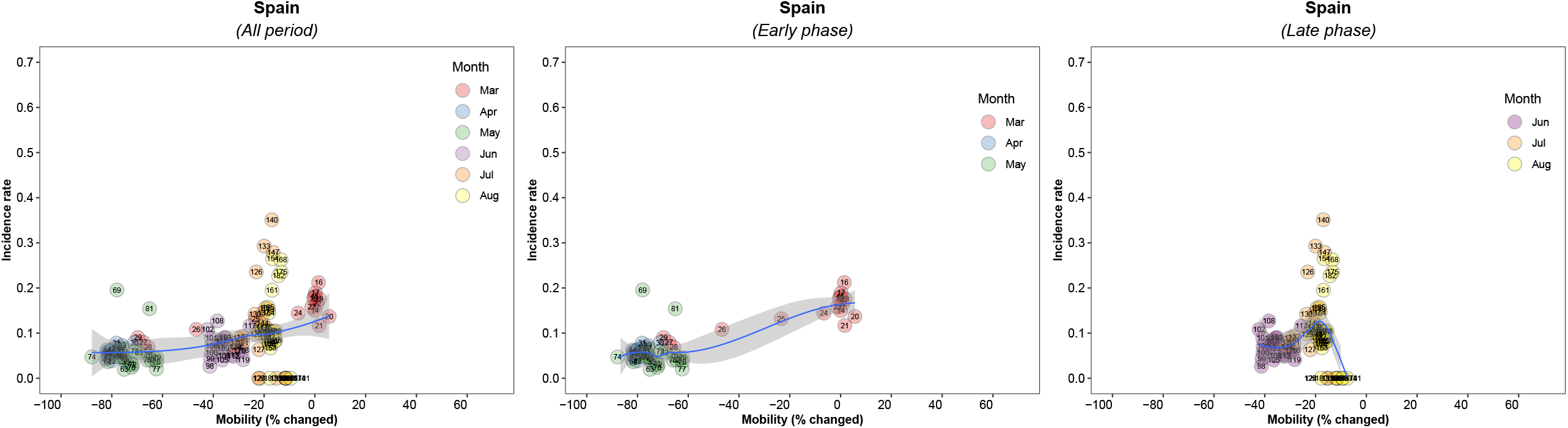

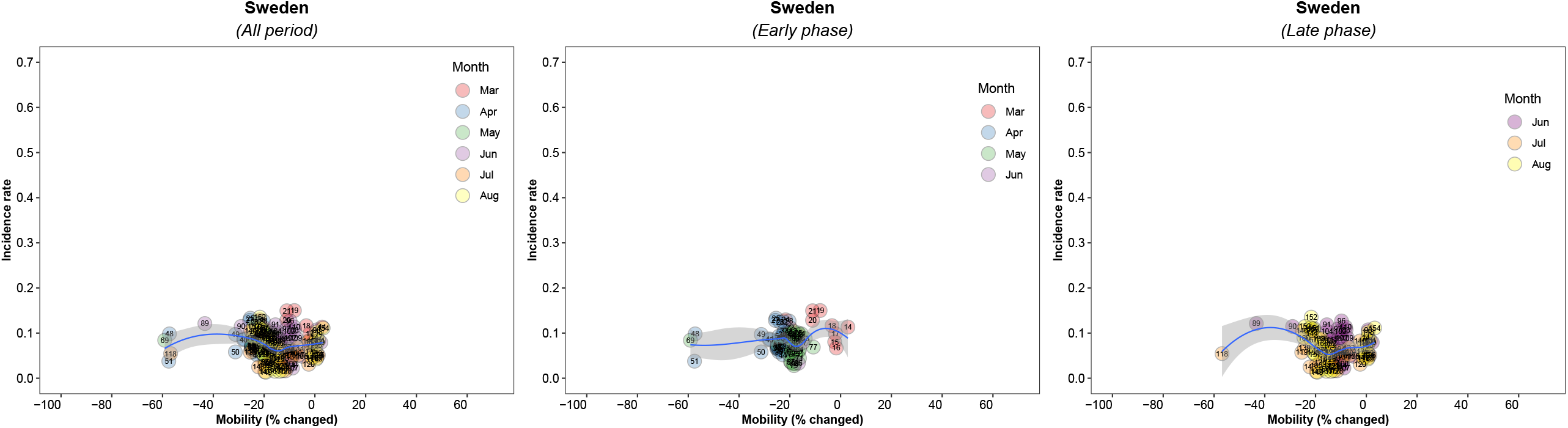

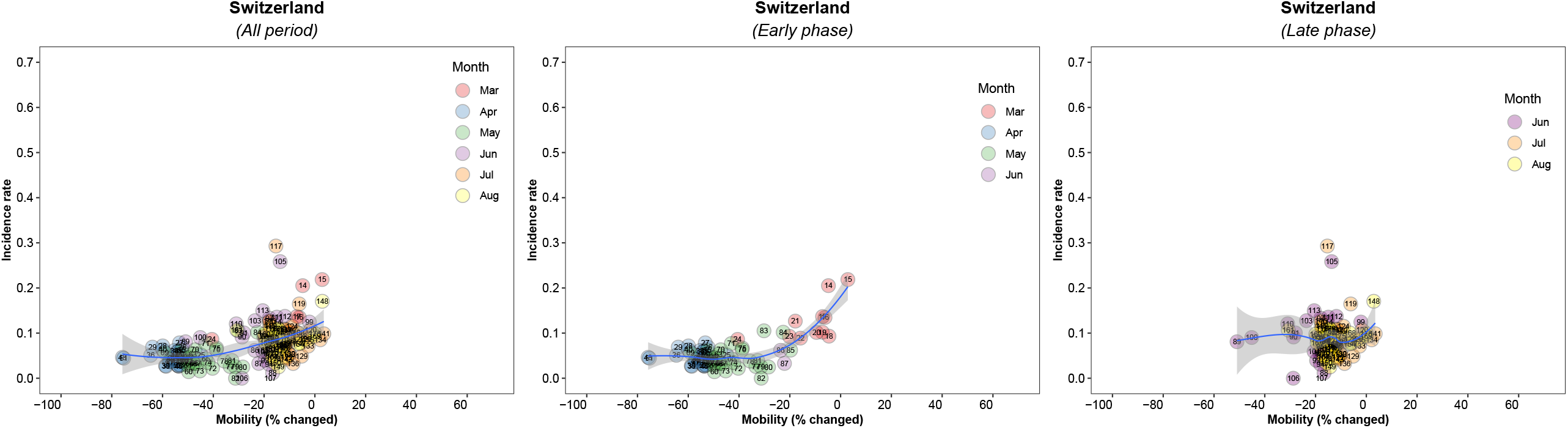

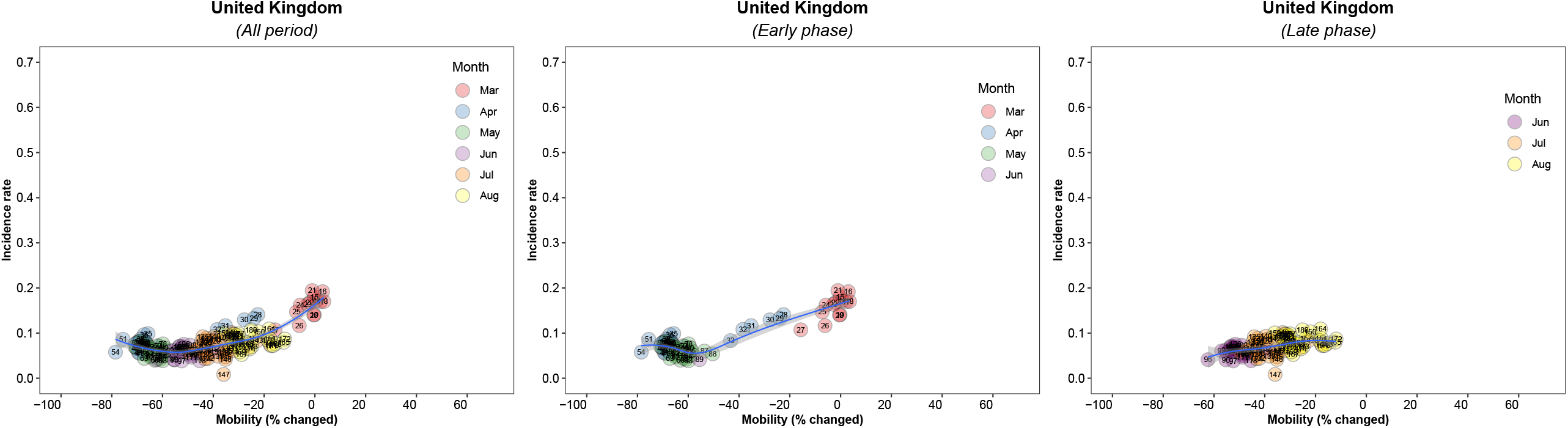

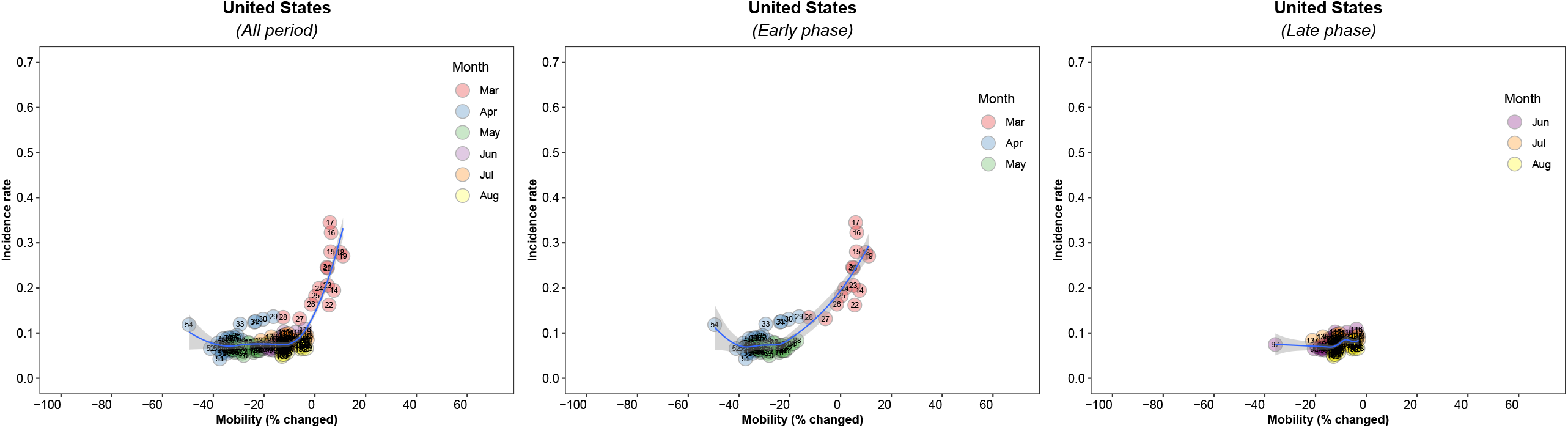

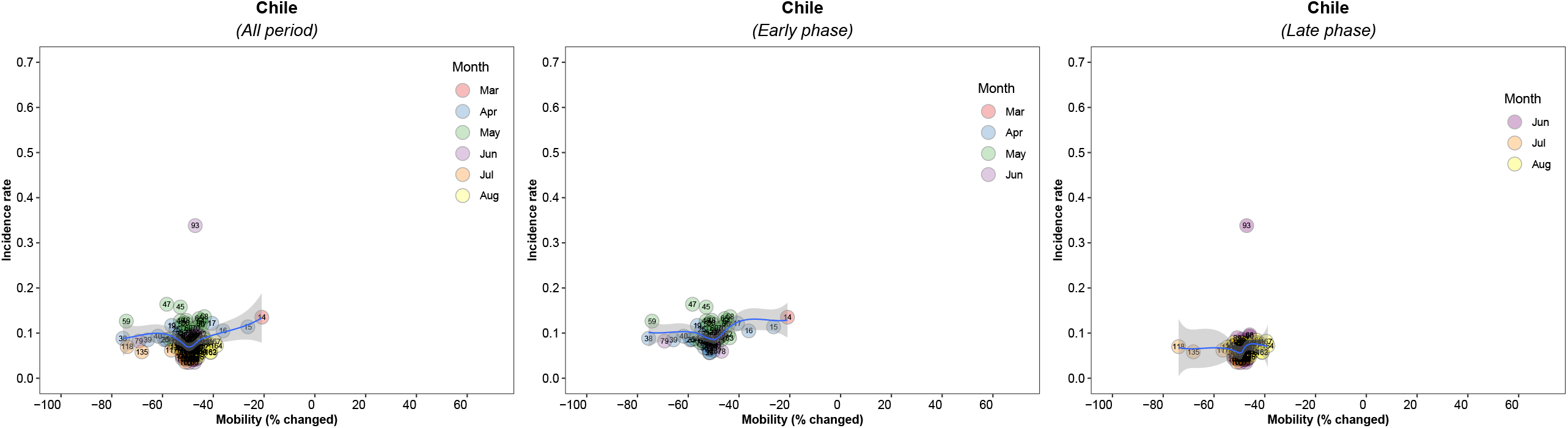

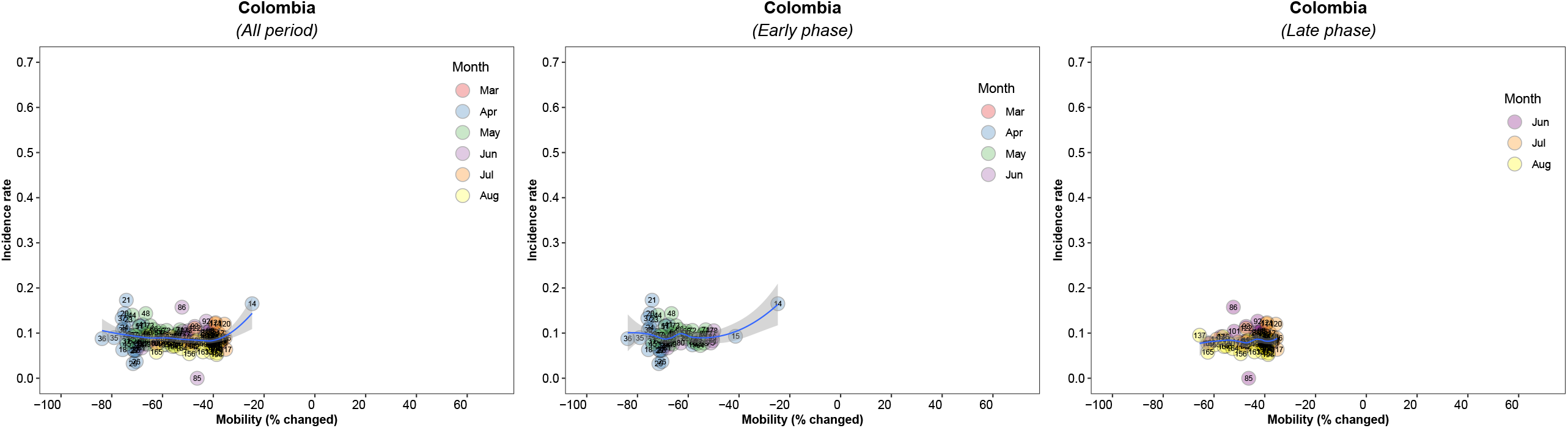

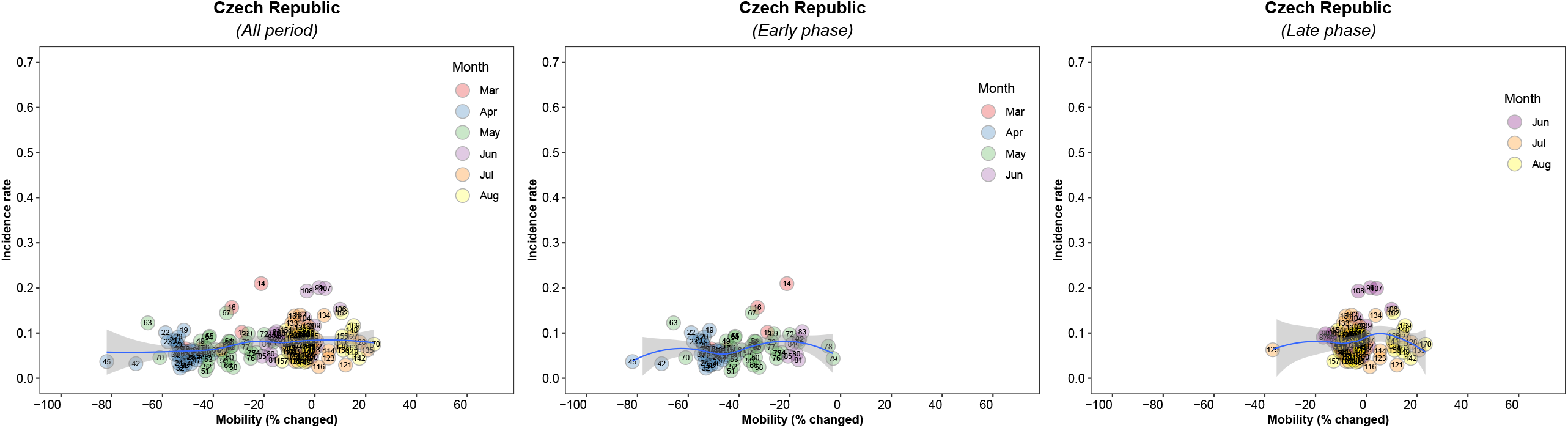

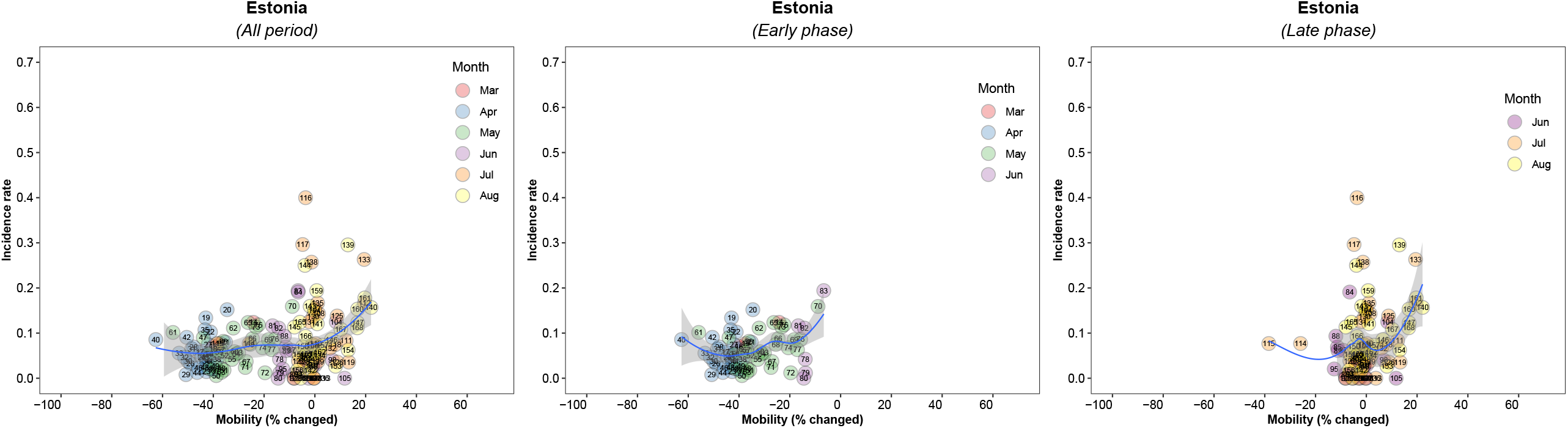

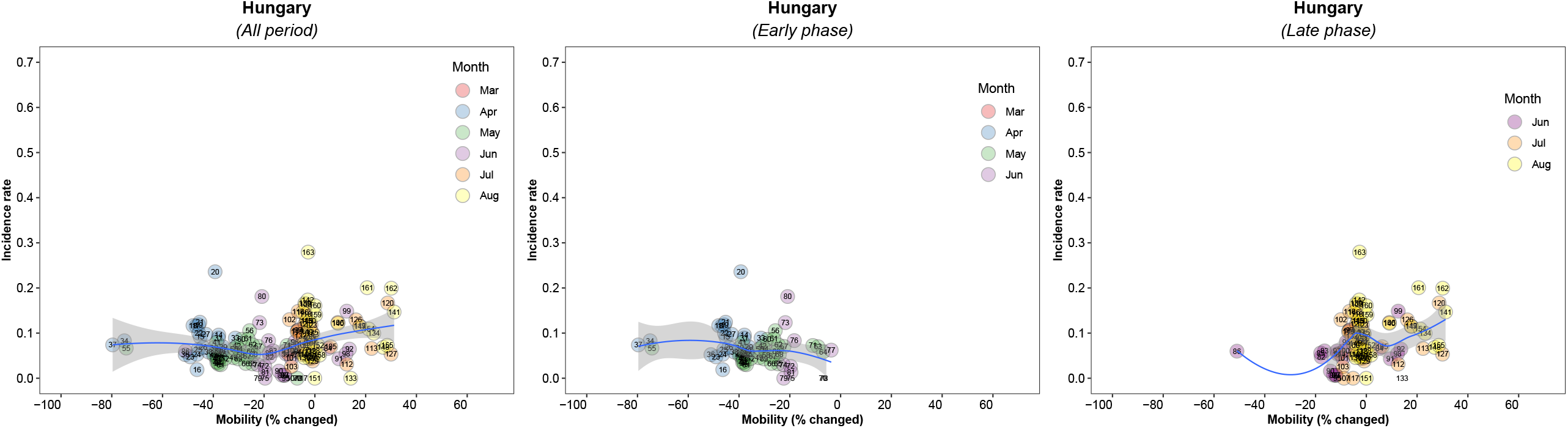

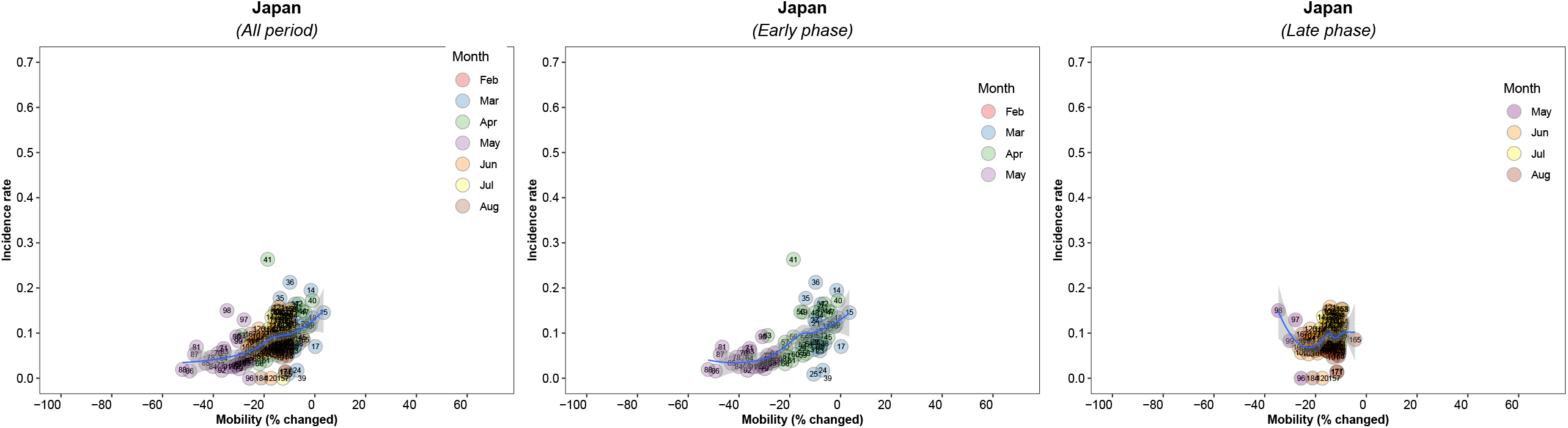

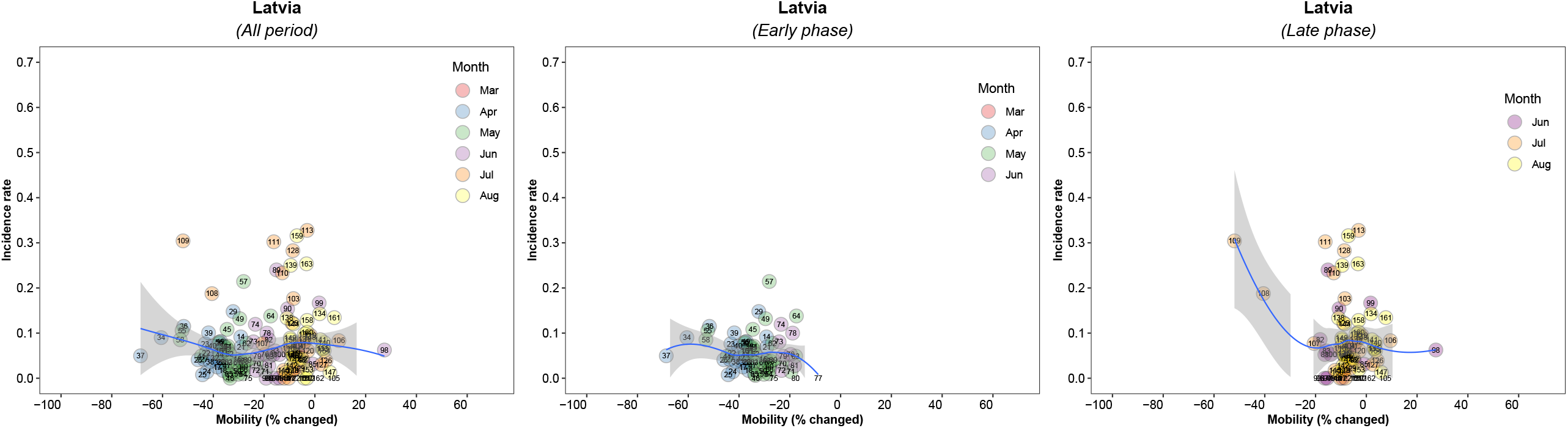

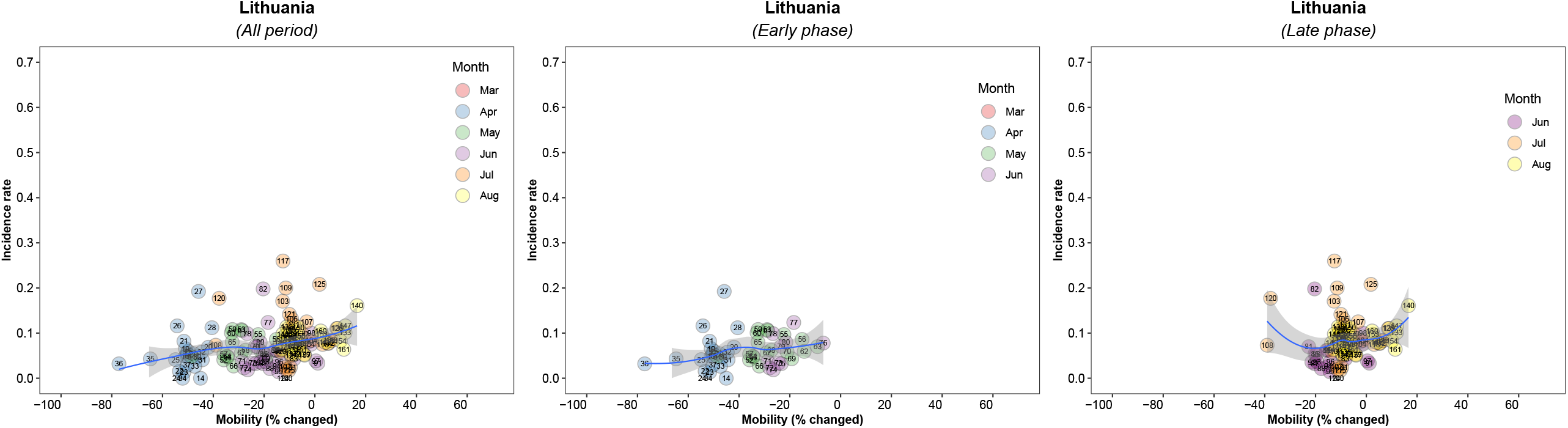

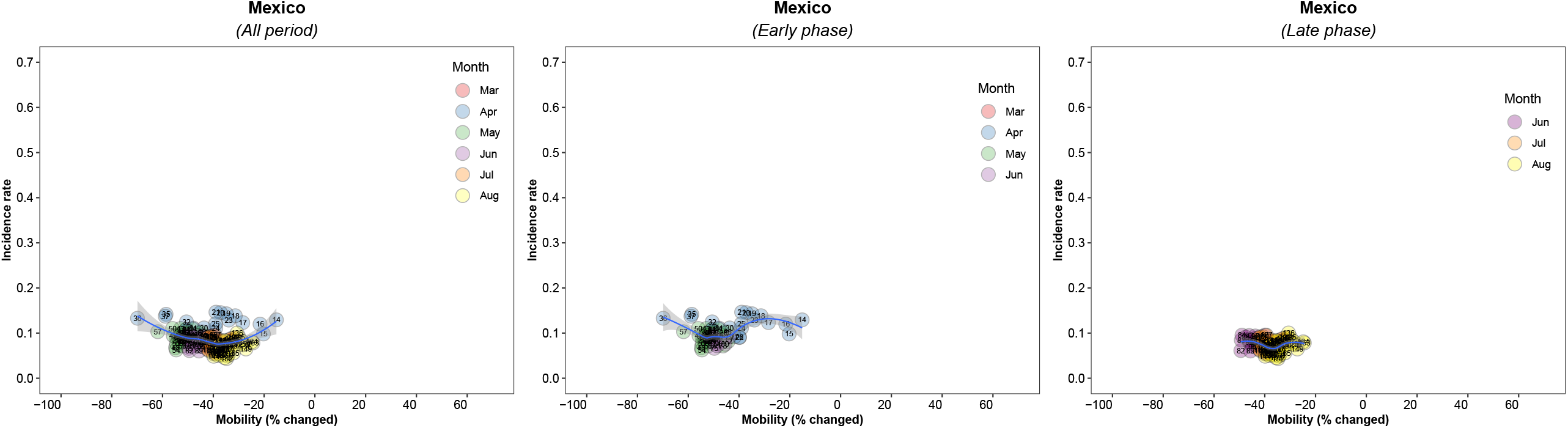

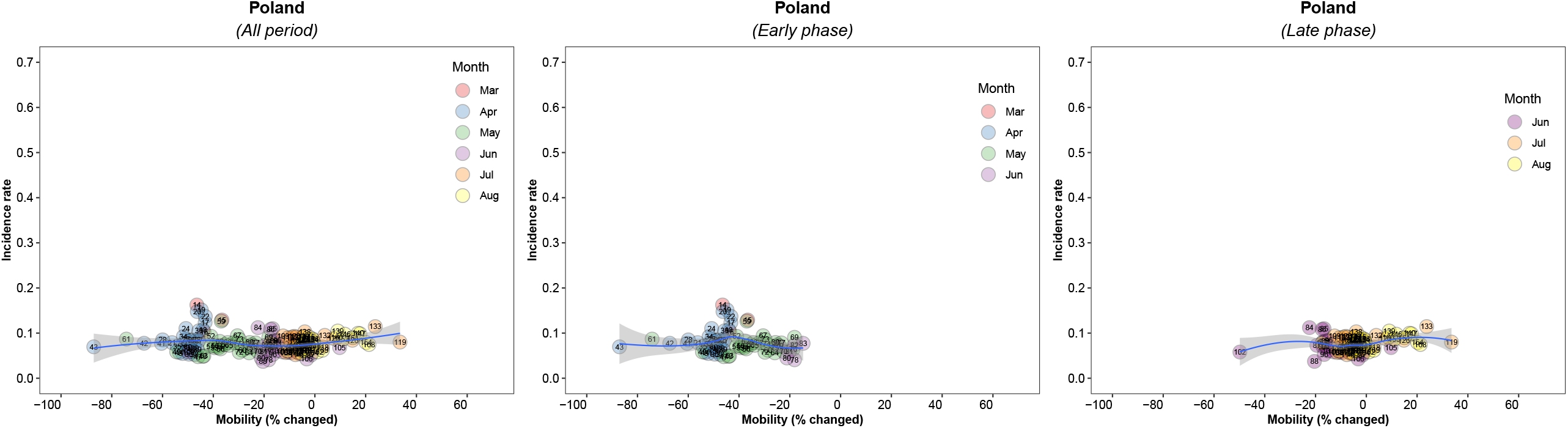

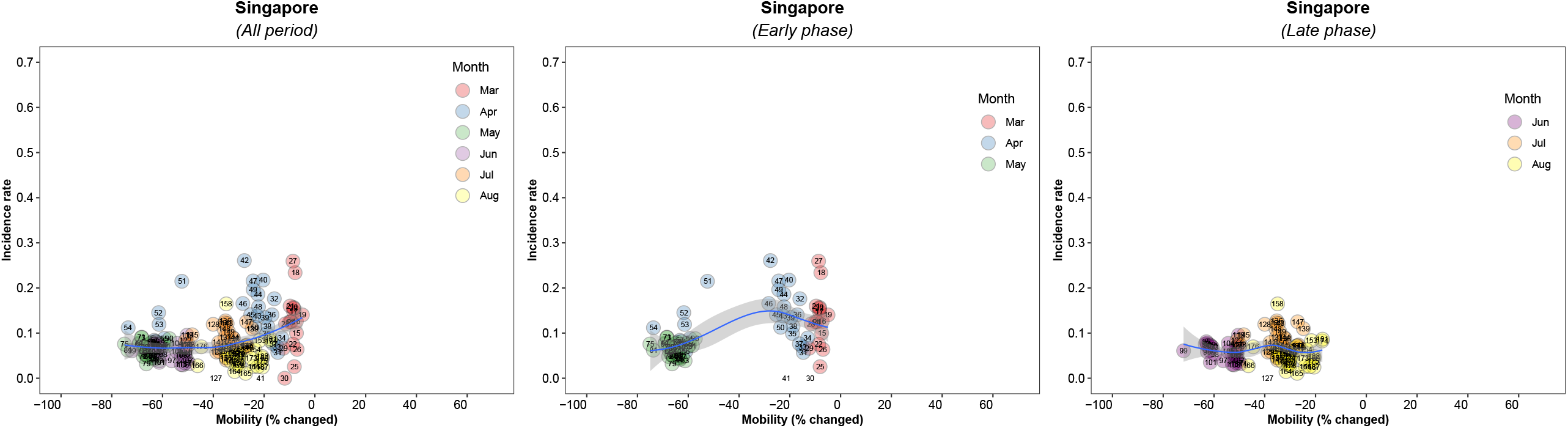

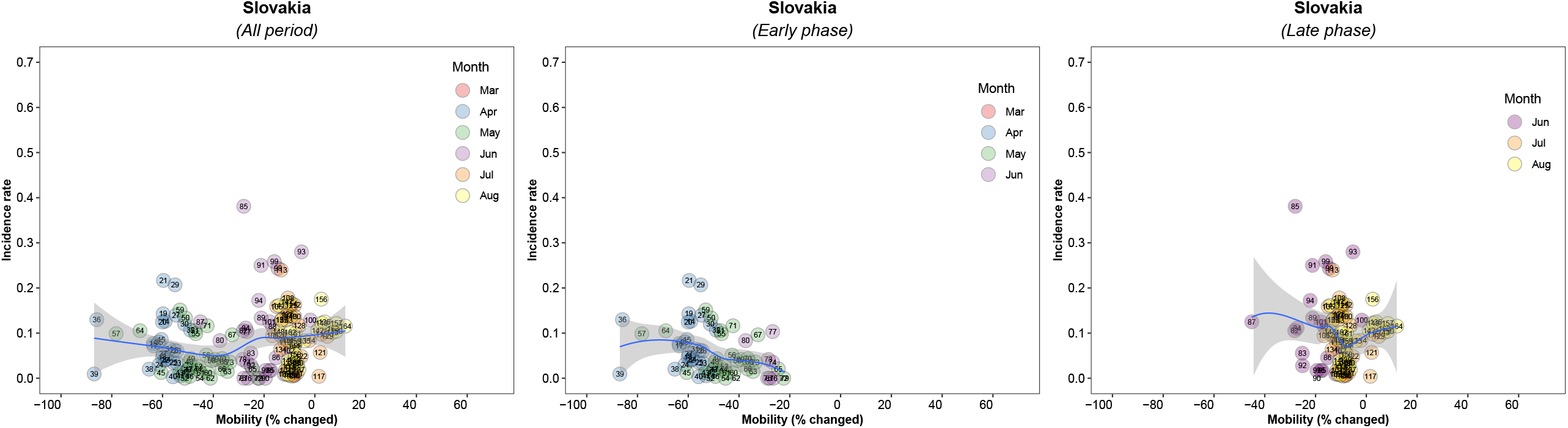

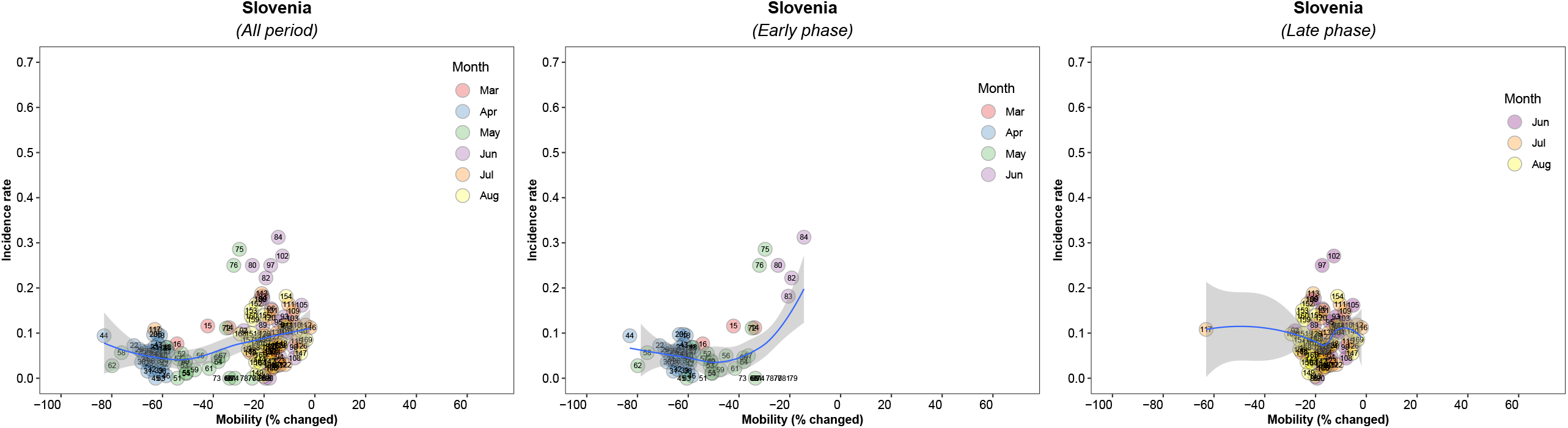

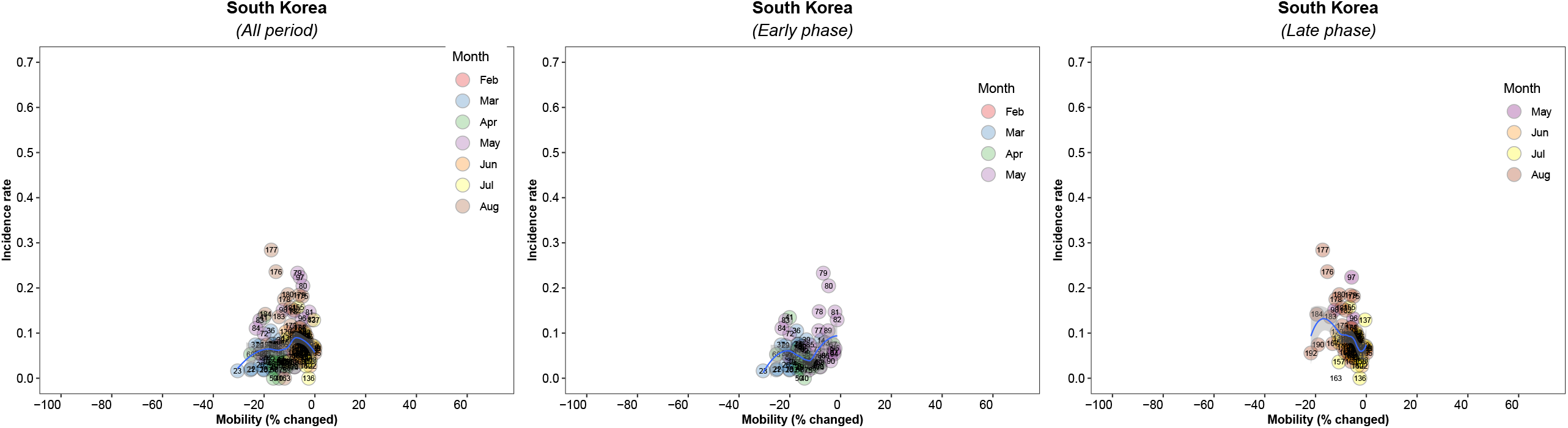

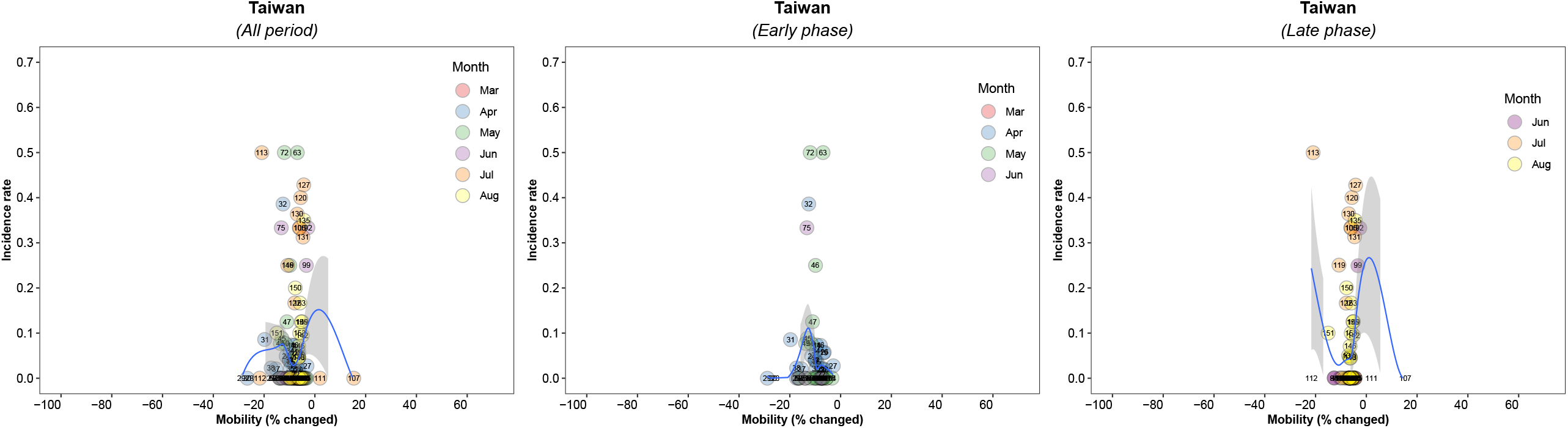

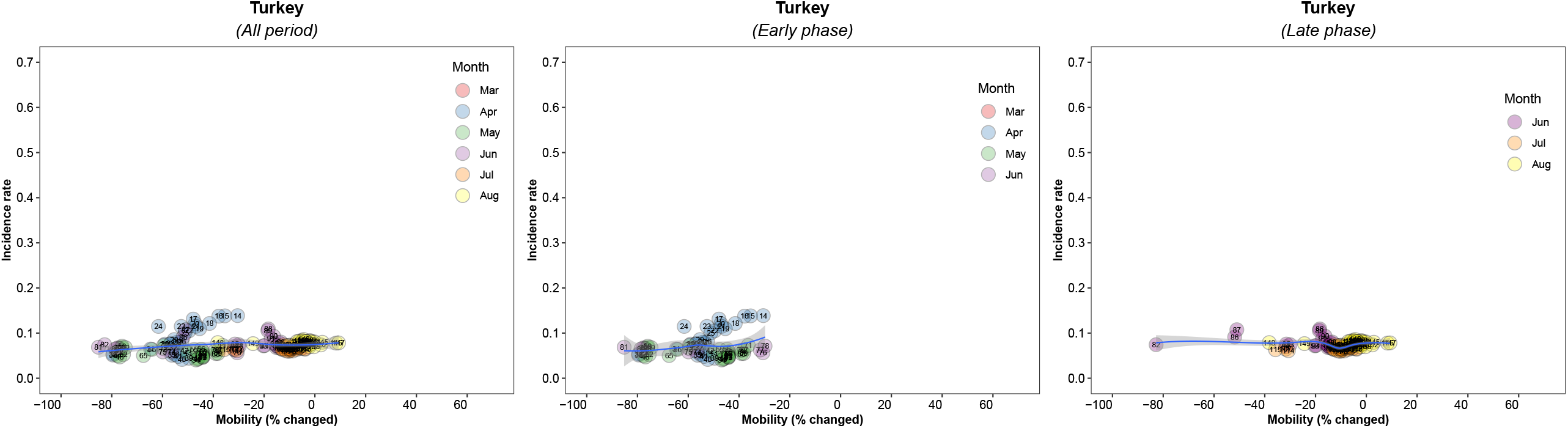

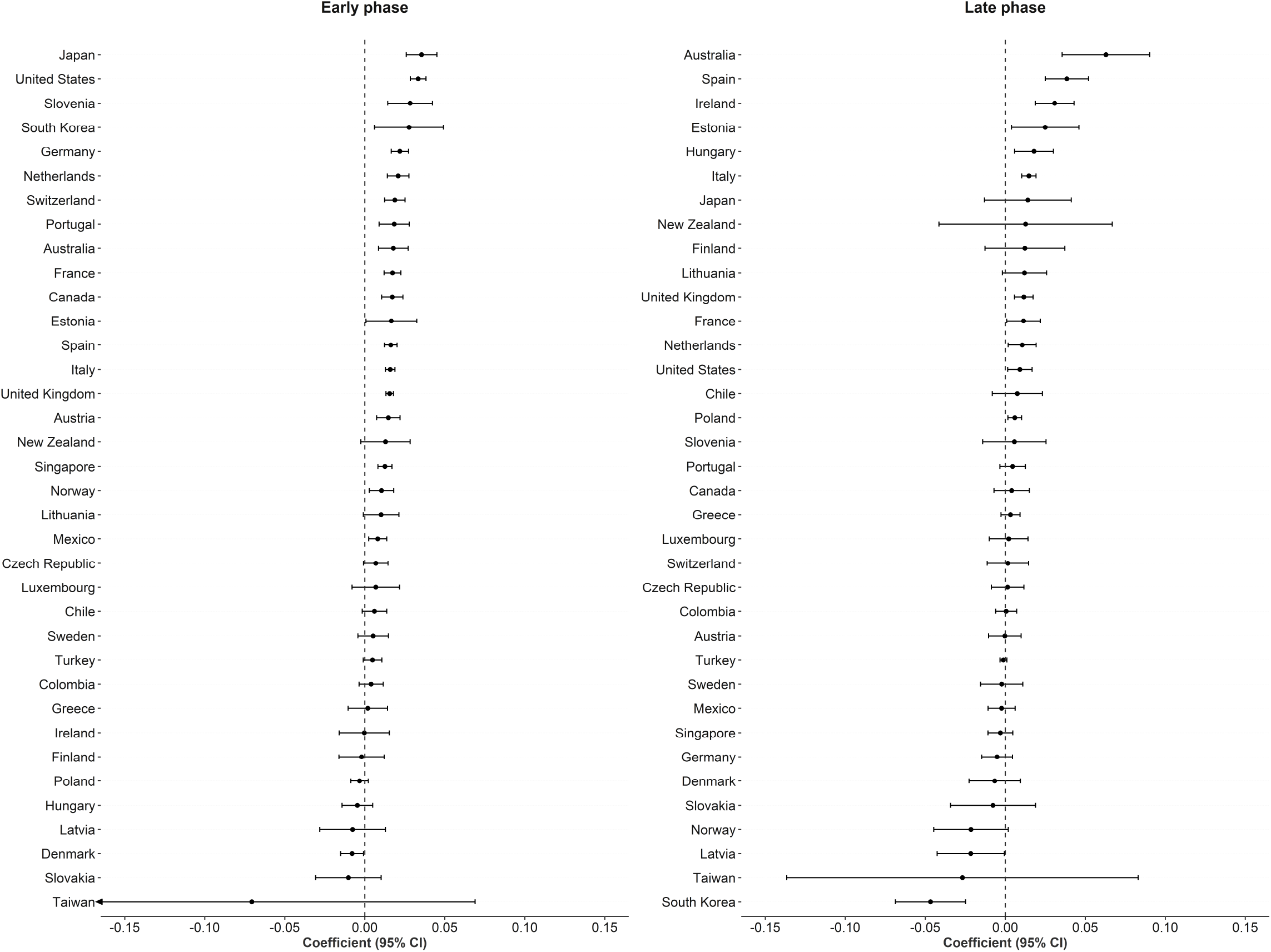

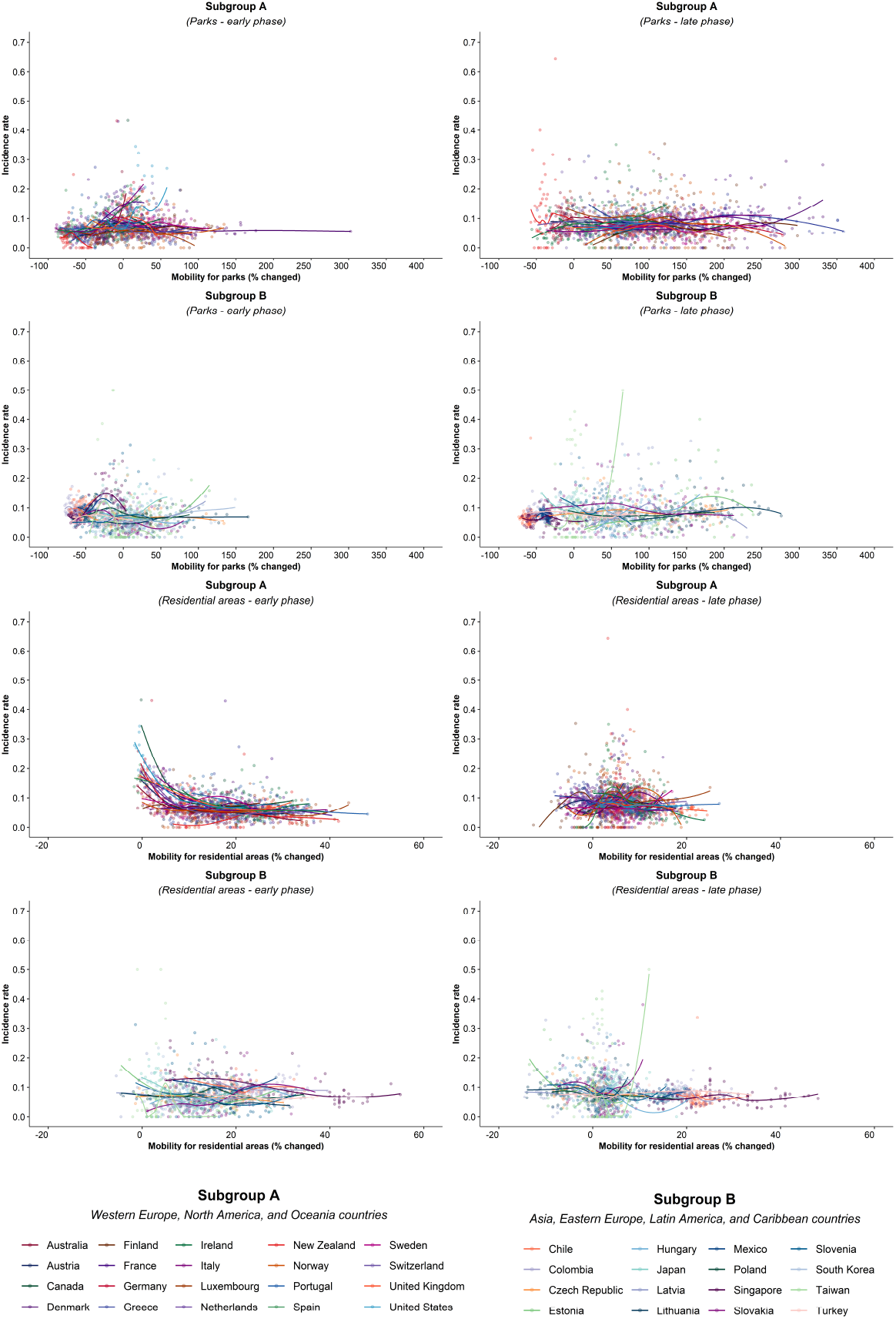

