## Supplemental figure 1~4 for "How well does societal mobility restriction help control the COVID-19 pandemic? Evidence from real-time evaluation"

Supplementary Figure 1. Correlation matrix between six categories of mobility locations: workplaces; transit stations; retailer and recreational places; residential areas; groceries and pharmacies; parks; and commuting (average of workplaces; transit stations; retailer and recreational places)

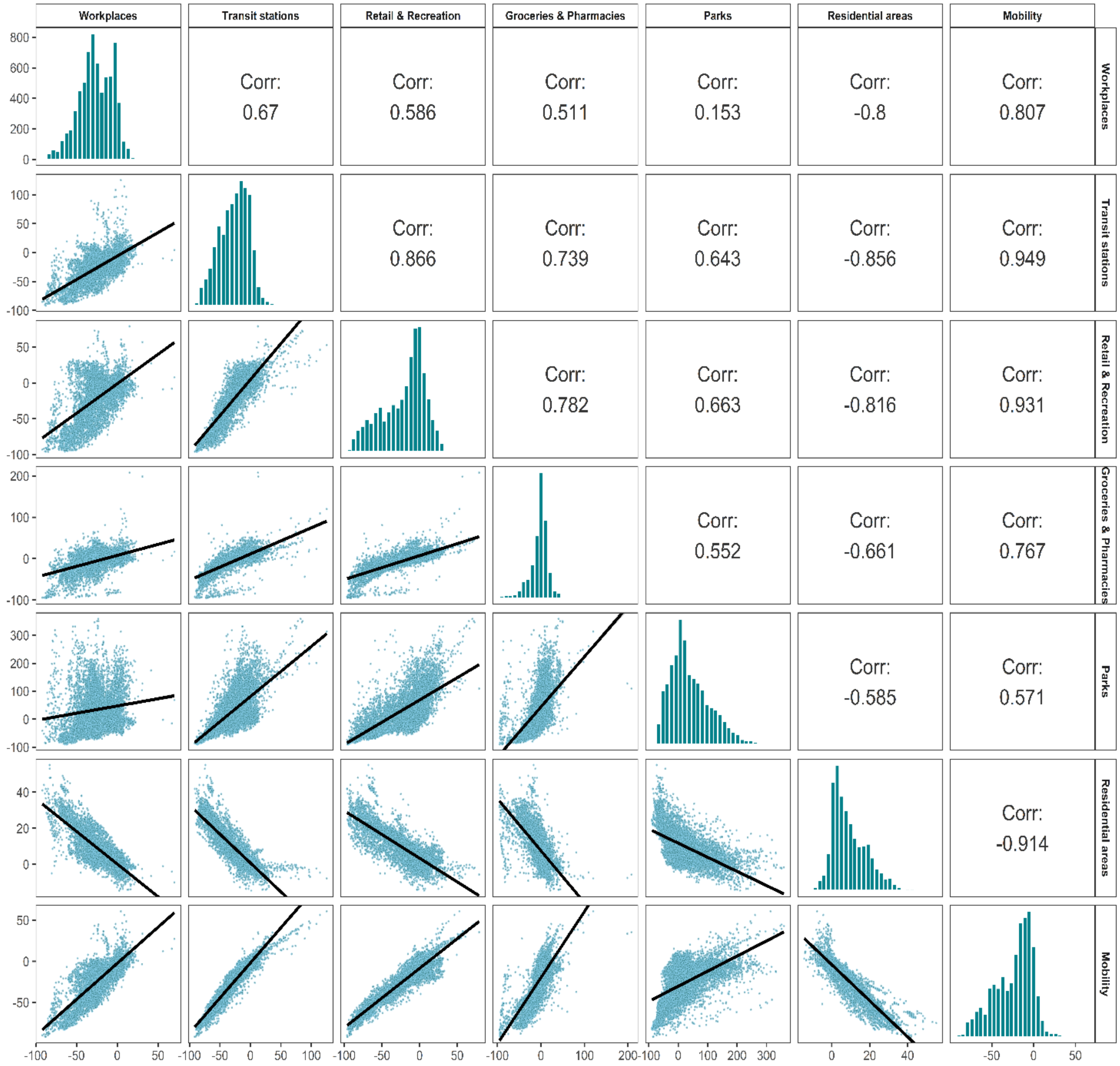

Supplementary Figure 2A. Association between new daily incidence rates of COVID-19 and mobility changes in 36 countries by early and late phase and other places (parks and residential areas) visited. Western Europe, North America, and Oceania

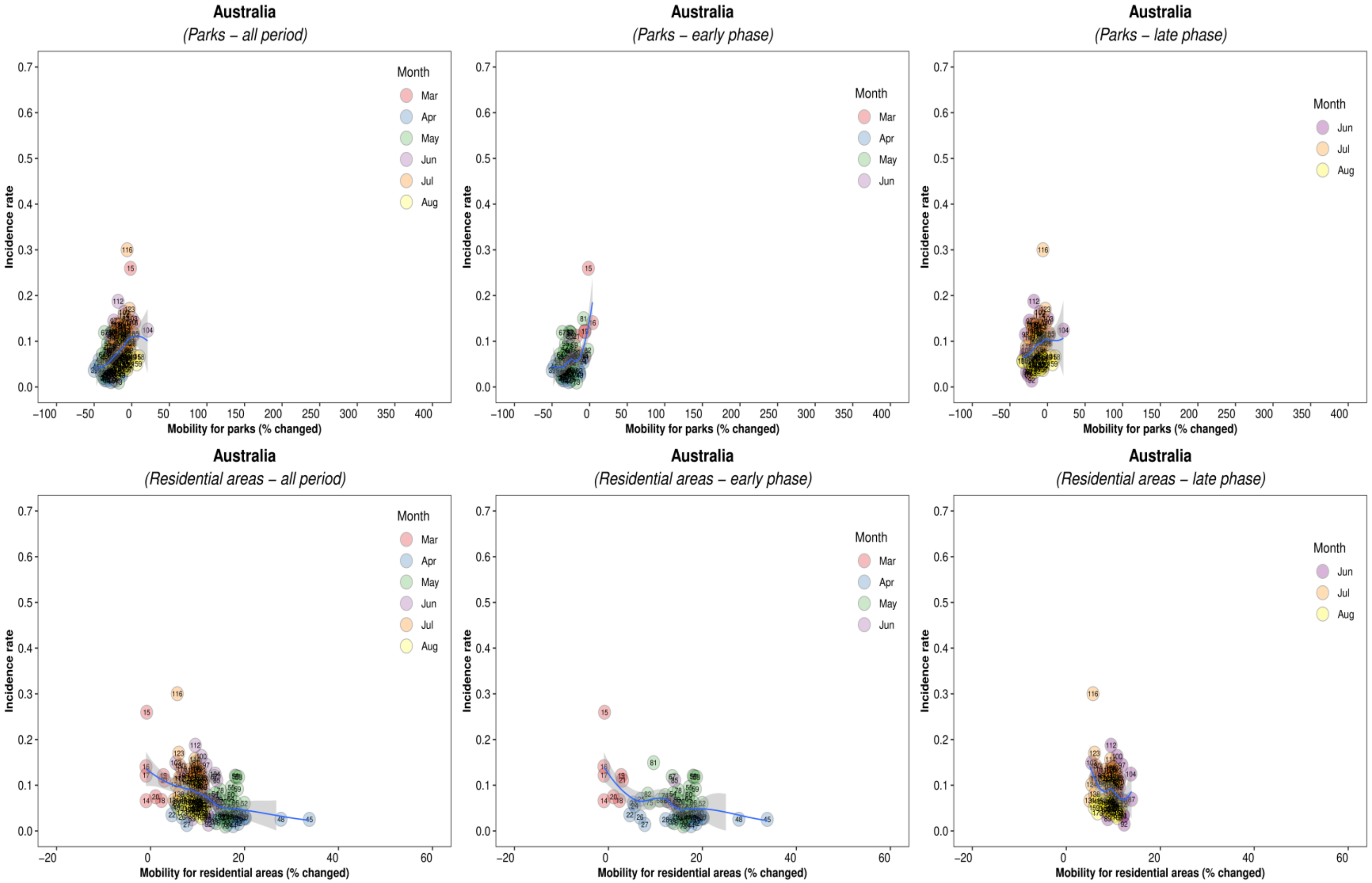

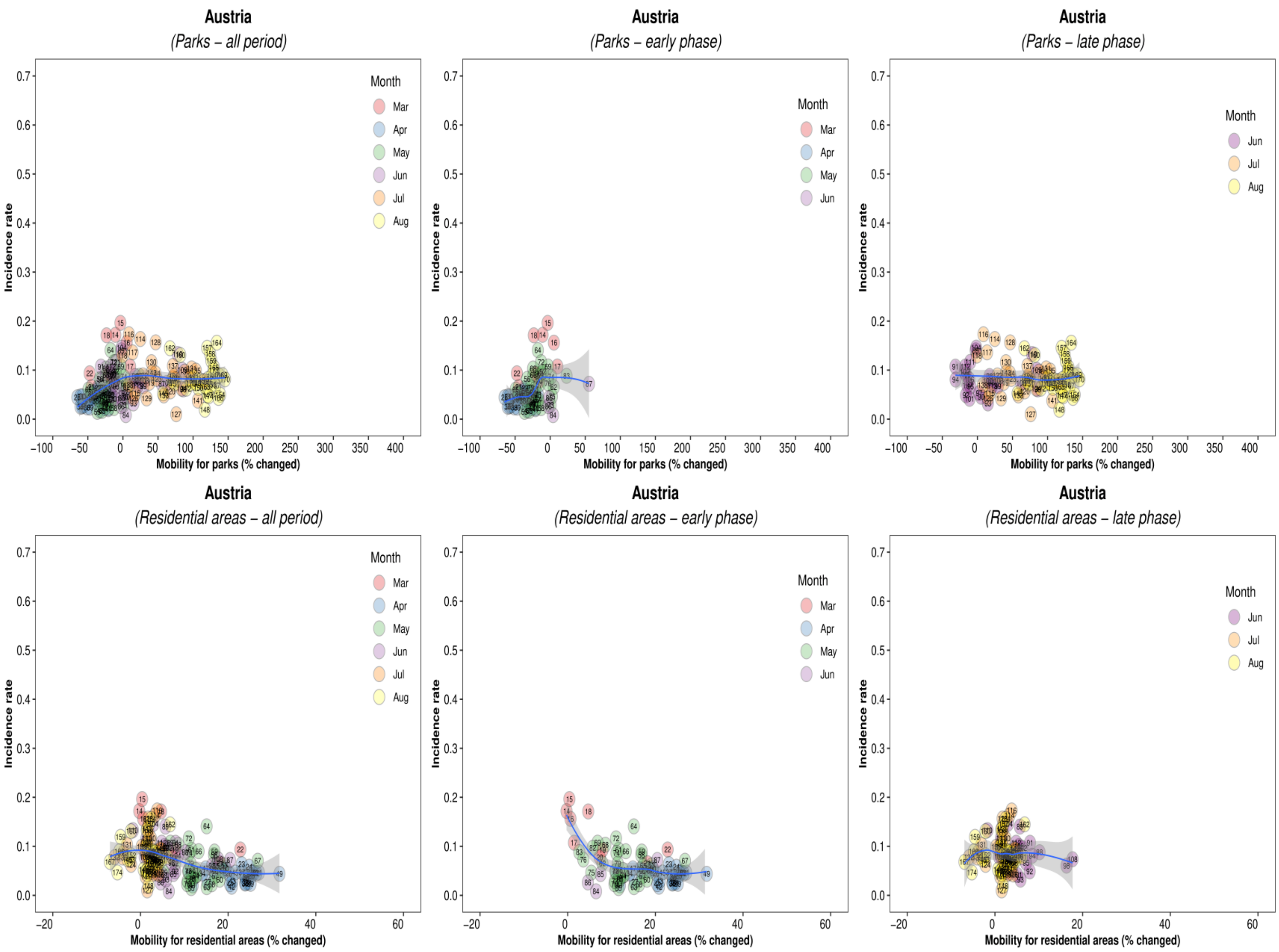

Austria
(Parks – all period)
Austria
(Parks – early phase)
Austria
(Parks – late phase)
Austria
(Residential areas – all period)
Austria
(Residential areas – early phase)
Austria
(Residential areas – late phase)
Incidence rate
Mobility for parks (% changed)
Mobility for residential areas (% changed)
Month
Mar
Apr
May
Jun
Jul
Aug

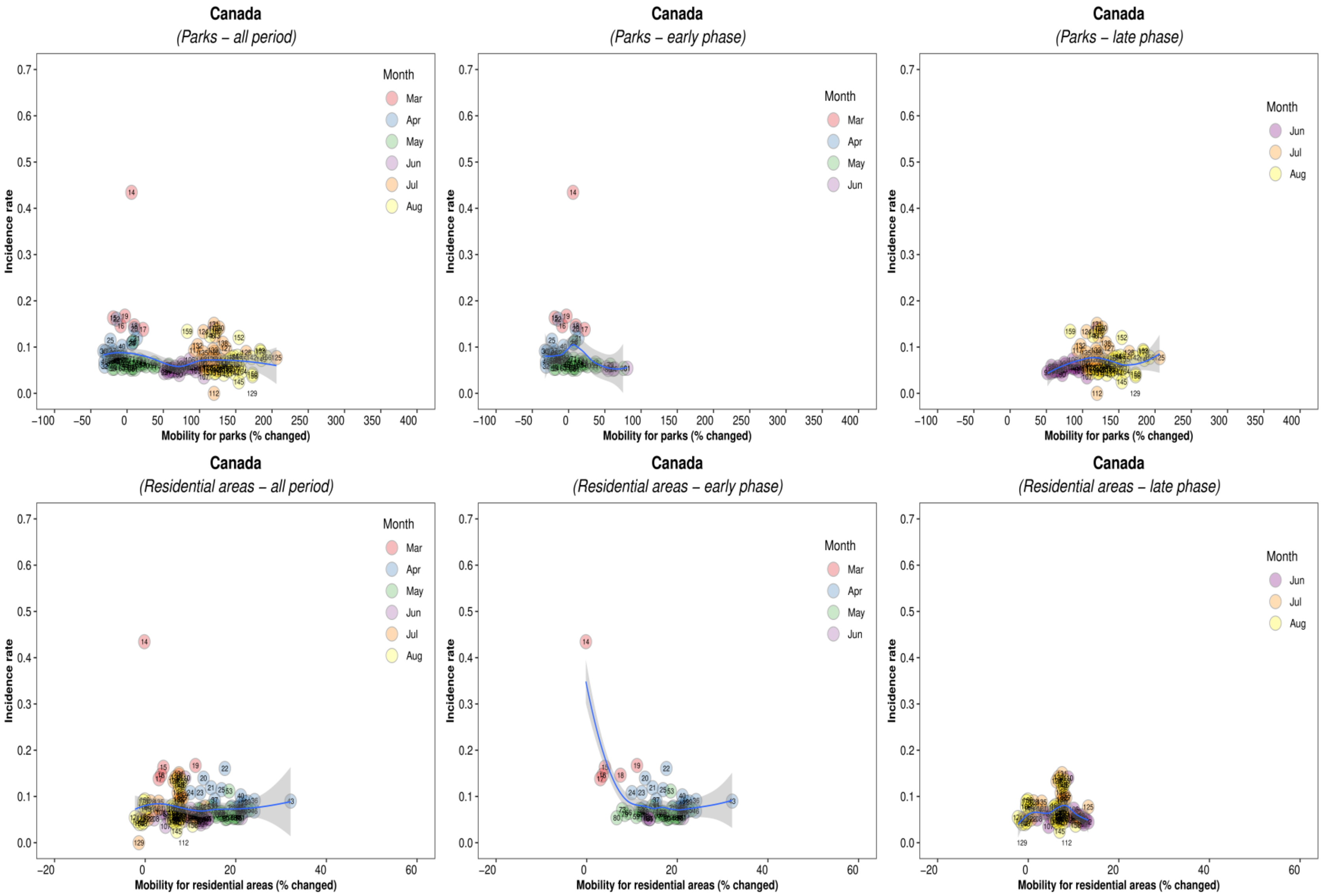

Canada
(Parks – all period)
Canada
(Parks – early phase)
Canada
(Parks – late phase)
Canada
(Residential areas – all period)
Canada
(Residential areas – early phase)
Canada
(Residential areas – late phase)
Incidence rate
Mobility for parks (% changed)
Mobility for residential areas (% changed)
Month
Mar
Apr
May
Jun
Jul
Aug

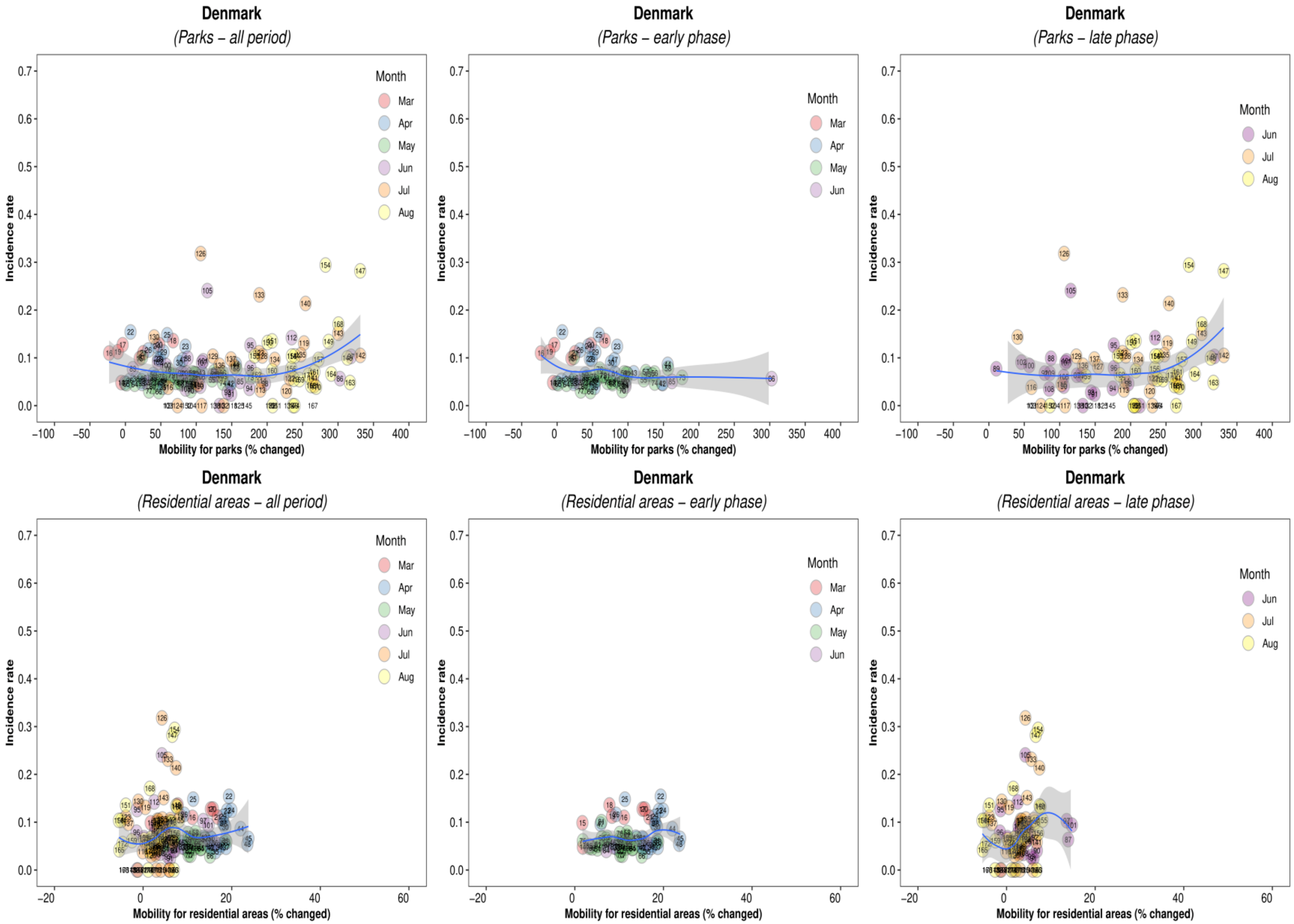
Denmark
(Parks – all period)
Denmark
(Parks – early phase)
Denmark
(Parks – late phase)
Denmark
(Residential areas – all period)
Denmark
(Residential areas – early phase)
Denmark
(Residential areas – late phase)
Month
Mar
Apr
May
Jun
Jul
Aug
Incidence rate
Mobility for parks (% changed)
Mobility for residential areas (% changed)

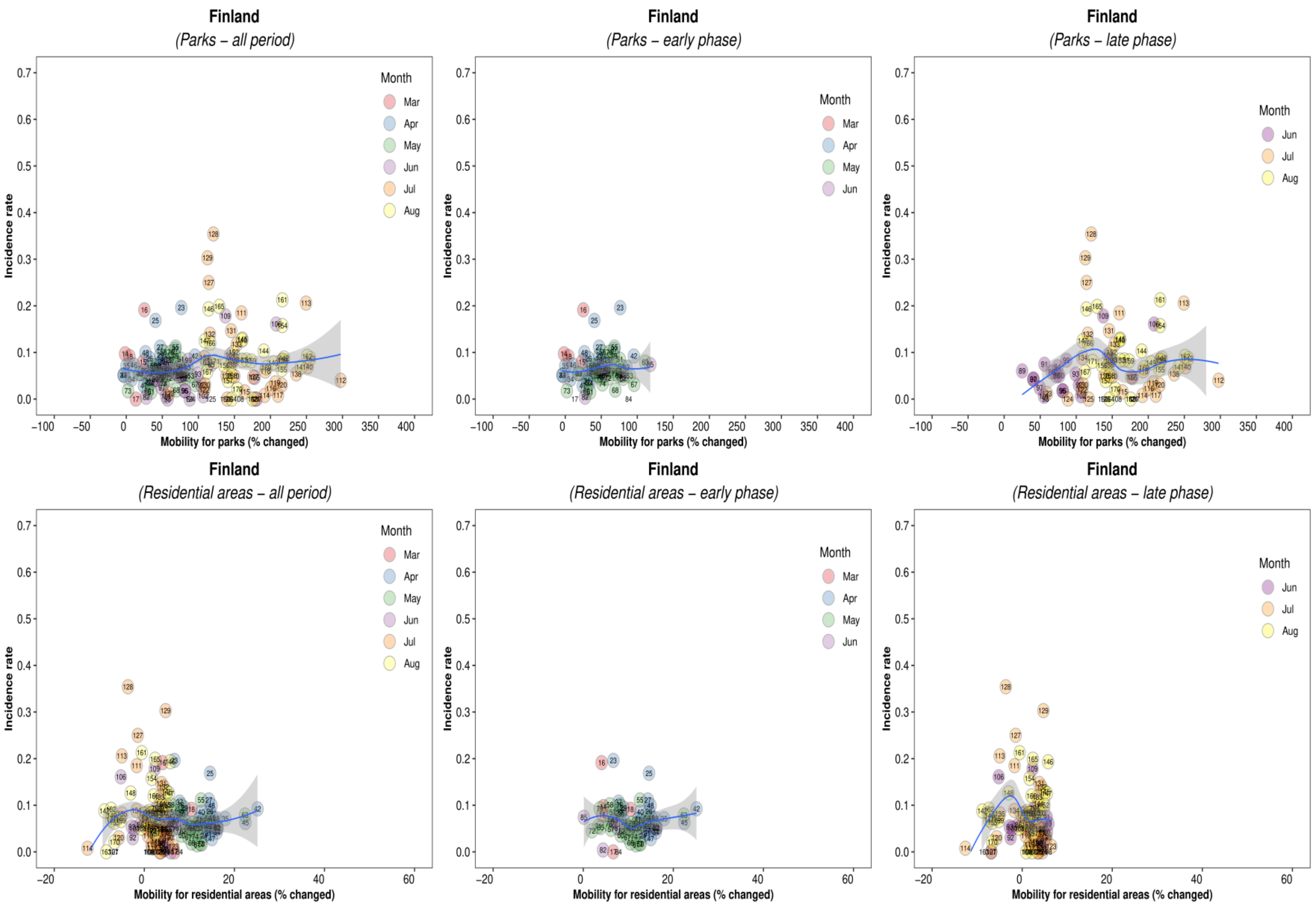
Finland
(Parks – all period)
Finland
(Parks – early phase)
Finland
(Parks – late phase)
Finland
(Residential areas – all period)
Finland
(Residential areas – early phase)
Finland
(Residential areas – late phase)
Incidence rate
Mobility for parks (% changed)
Mobility for residential areas (% changed)
Month
Mar
Apr
May
Jun
Jul
Aug

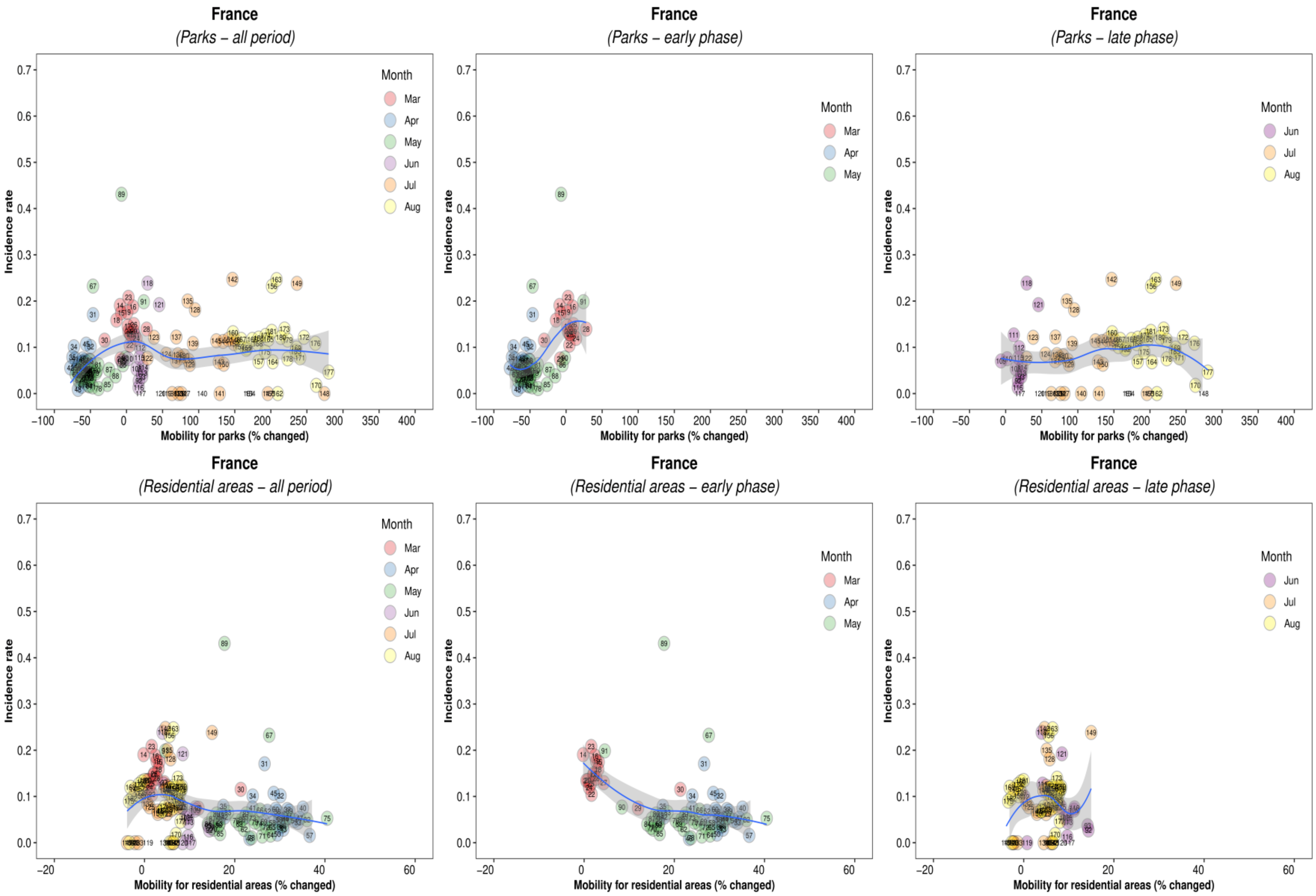
France
(Parks – all period)
Month
Mar
Apr
May
Jun
Jul
Aug
Incidence rate
Mobility for parks (% changed)
France
(Parks – early phase)
Month
Mar
Apr
May
Incidence rate
Mobility for parks (% changed)
France
(Parks – late phase)
Month
Jun
Jul
Aug
Incidence rate
Mobility for parks (% changed)
France
(Residential areas – all period)
Month
Mar
Apr
May
Jun
Jul
Aug
Incidence rate
Mobility for residential areas (% changed)
France
(Residential areas – early phase)
Month
Mar
Apr
May
Incidence rate
Mobility for residential areas (% changed)
France
(Residential areas – late phase)
Month
Jun
Jul
Aug
Incidence rate
Mobility for residential areas (% changed)

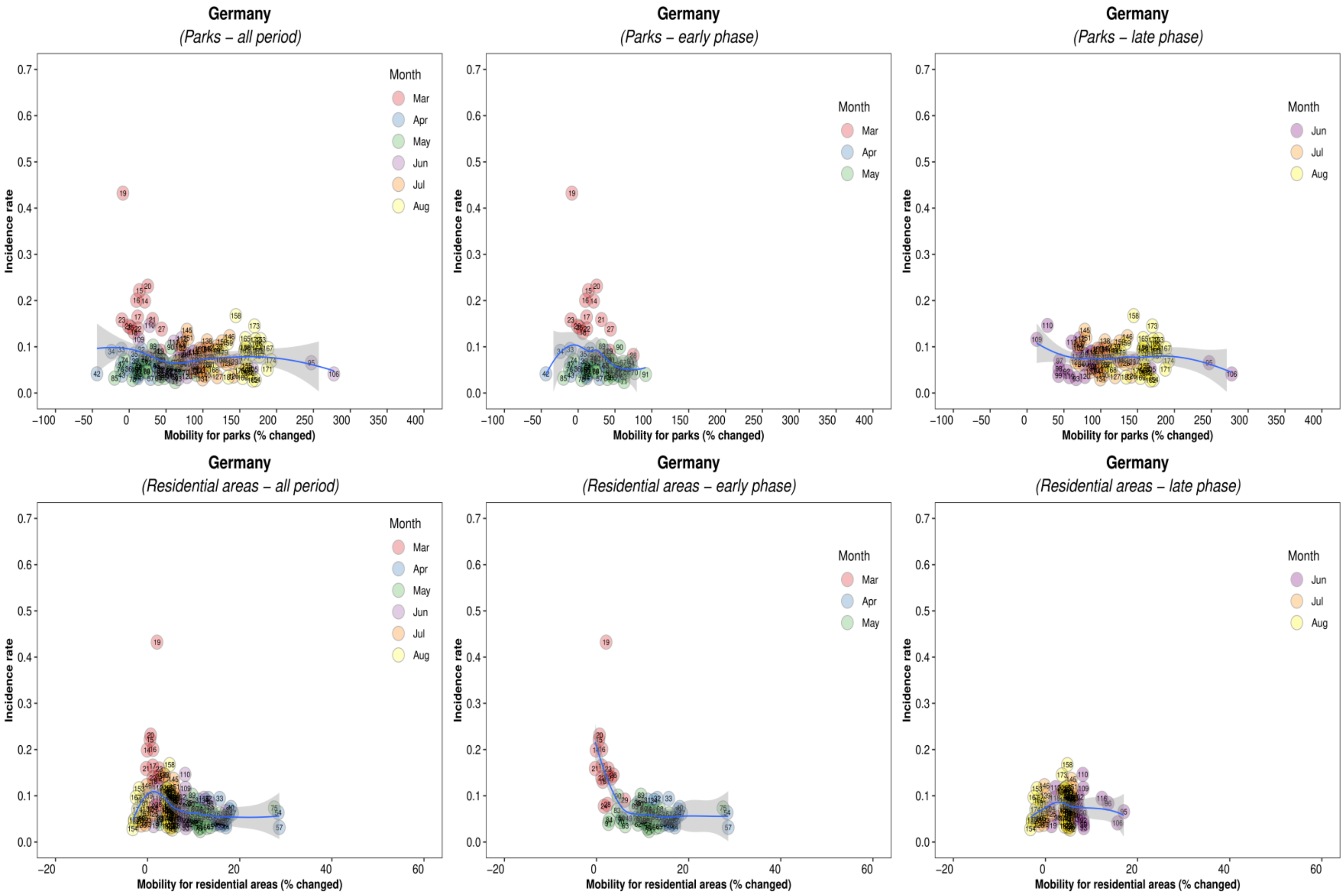
Germany
(Parks – all period)
Germany
(Parks – early phase)
Germany
(Parks – late phase)
Germany
(Residential areas – all period)
Germany
(Residential areas – early phase)
Germany
(Residential areas – late phase)
Incidence rate
Mobility for parks (% changed)
Mobility for residential areas (% changed)
Month
Mar
Apr
May
Jun
Jul
Aug

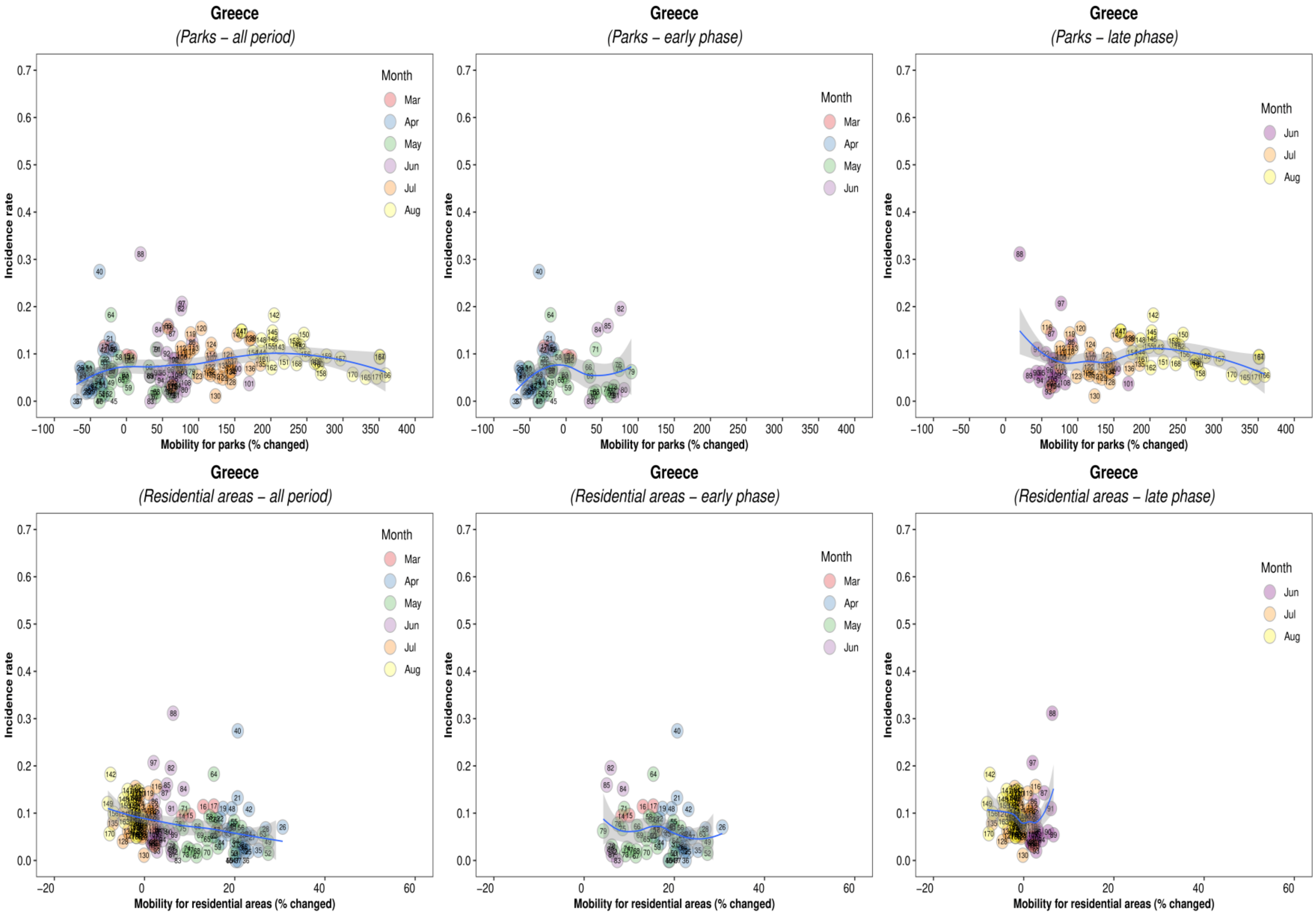
Greece
(Parks – all period)
Greece
(Parks – early phase)
Greece
(Parks – late phase)
Greece
(Residential areas – all period)
Greece
(Residential areas – early phase)
Greece
(Residential areas – late phase)
Incidence rate
Mobility for parks (% changed)
Mobility for residential areas (% changed)
Month
Mar
Apr
May
Jun
Jul
Aug

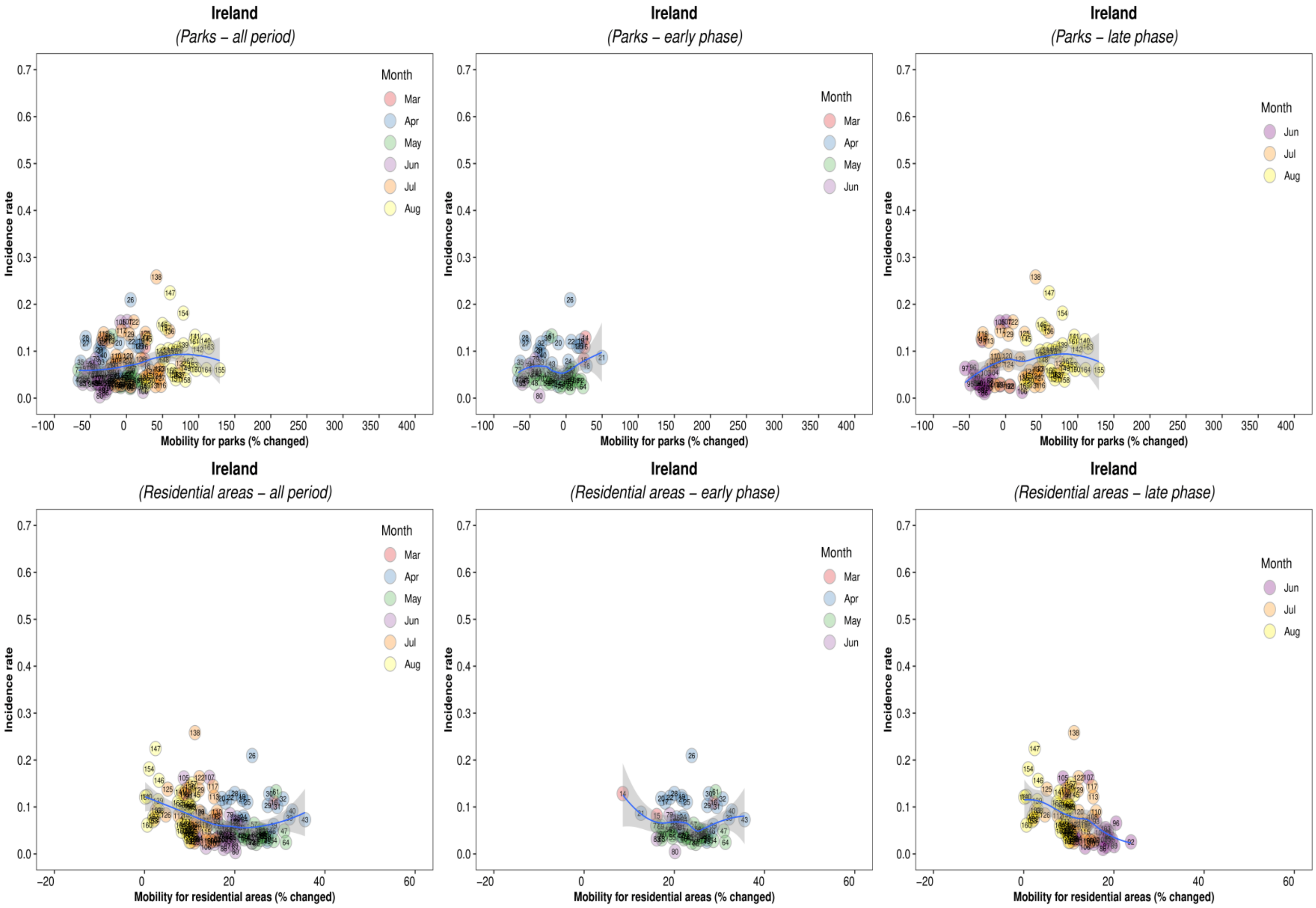

Ireland
(Parks – all period)
Ireland
(Parks – early phase)
Ireland
(Parks – late phase)
Ireland
(Residential areas – all period)
Ireland
(Residential areas – early phase)
Ireland
(Residential areas – late phase)
Month
Mar
Apr
May
Jun
Jul
Aug
Incidence rate
0.0
0.1
0.2
0.3
0.4
0.5
0.6
0.7
Mobility for parks (% changed)
−100
−50
0
50
100
150
200
250
300
350
400
Mobility for residential areas (% changed)
−20
0
20
40
60

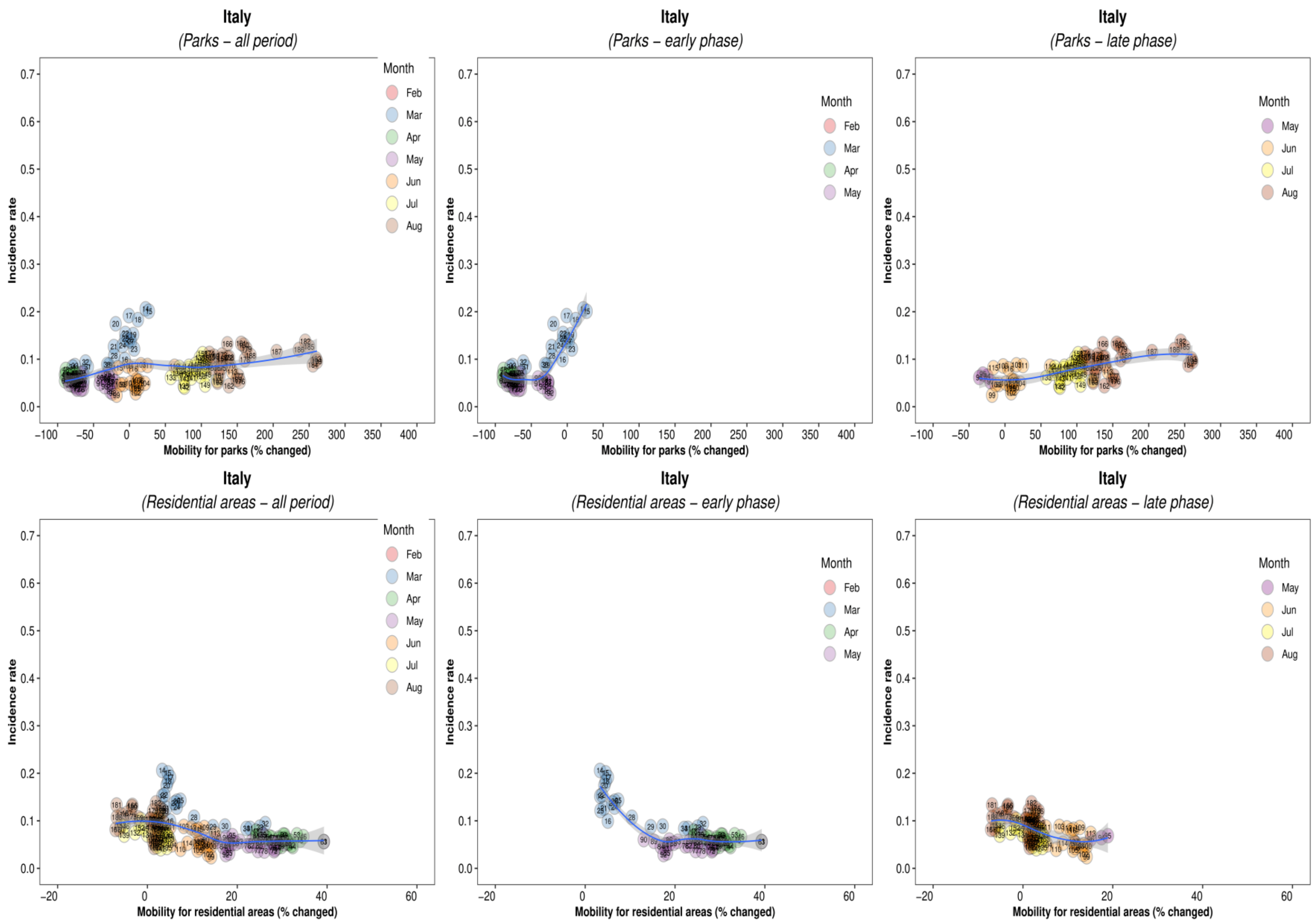

Italy
(Parks – all period)
Italy
(Parks – early phase)
Italy
(Parks – late phase)
Italy
(Residential areas – all period)
Italy
(Residential areas – early phase)
Italy
(Residential areas – late phase)
Incidence rate
Mobility for parks (% changed)
Mobility for residential areas (% changed)
Month
Feb
Mar
Apr
May
Jun
Jul
Aug
0.0
0.1
0.2
0.3
0.4
0.5
0.6
0.7
−100
−50
0
50
100
150
200
250
300
350
400
−20
0
20
40
60

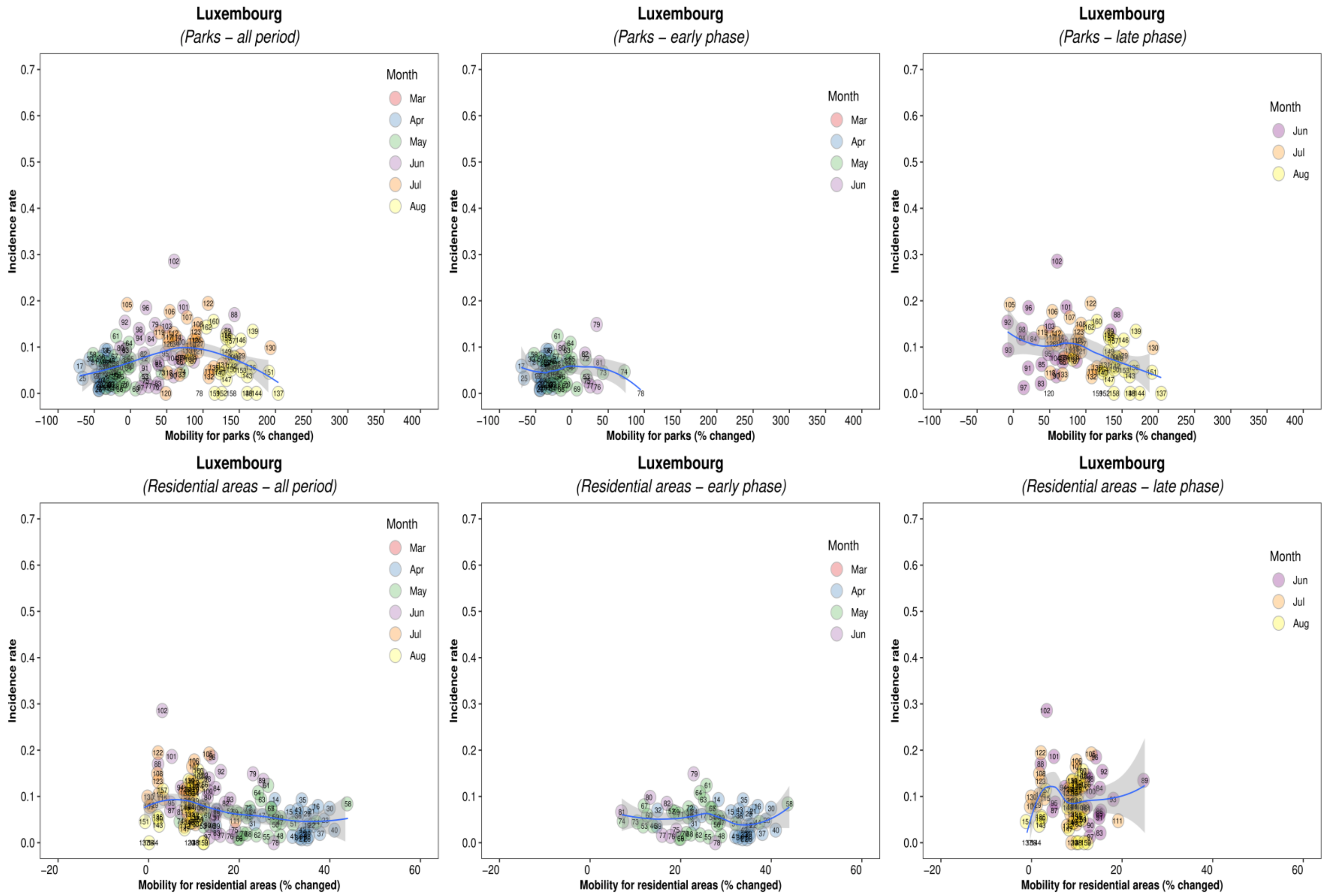
Luxembourg
(Parks – all period)
Incidence rate
Mobility for parks (% changed)
Month
Mar
Apr
May
Jun
Jul
Aug
Luxembourg
(Parks – early phase)
Incidence rate
Mobility for parks (% changed)
Month
Mar
Apr
May
Jun
Luxembourg
(Parks – late phase)
Incidence rate
Mobility for parks (% changed)
Month
Jun
Jul
Aug
Luxembourg
(Residential areas – all period)
Incidence rate
Mobility for residential areas (% changed)
Month
Mar
Apr
May
Jun
Jul
Aug
Luxembourg
(Residential areas – early phase)
Incidence rate
Mobility for residential areas (% changed)
Month
Mar
Apr
May
Jun
Luxembourg
(Residential areas – late phase)
Incidence rate
Mobility for residential areas (% changed)
Month
Jun
Jul
Aug

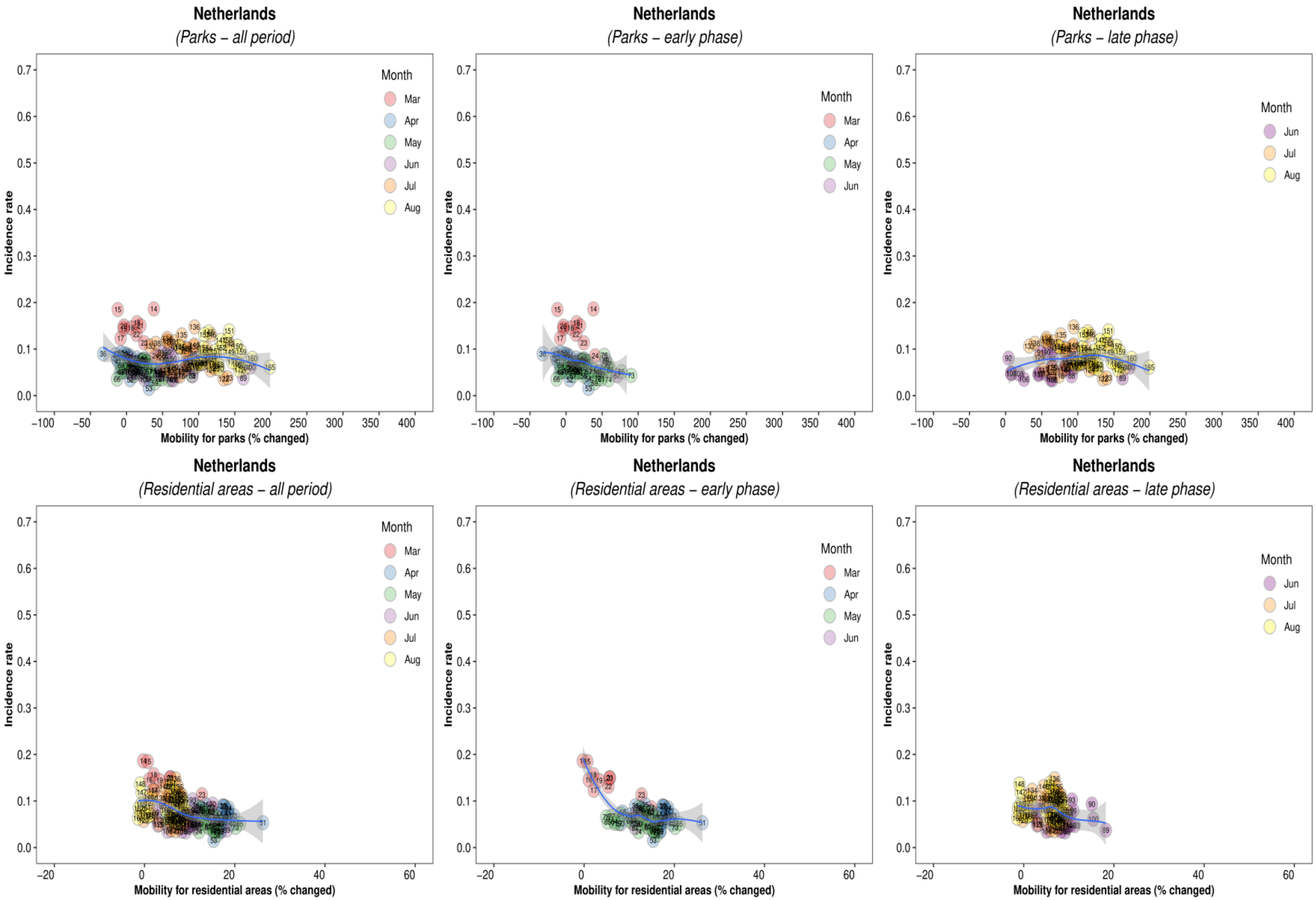
Netherlands
(Parks – all period)
Netherlands
(Parks – early phase)
Netherlands
(Parks – late phase)
Netherlands
(Residential areas – all period)
Netherlands
(Residential areas – early phase)
Netherlands
(Residential areas – late phase)
Incidence rate
Mobility for parks (% changed)
Mobility for residential areas (% changed)
Month
Mar
Apr
May
Jun
Jul
Aug

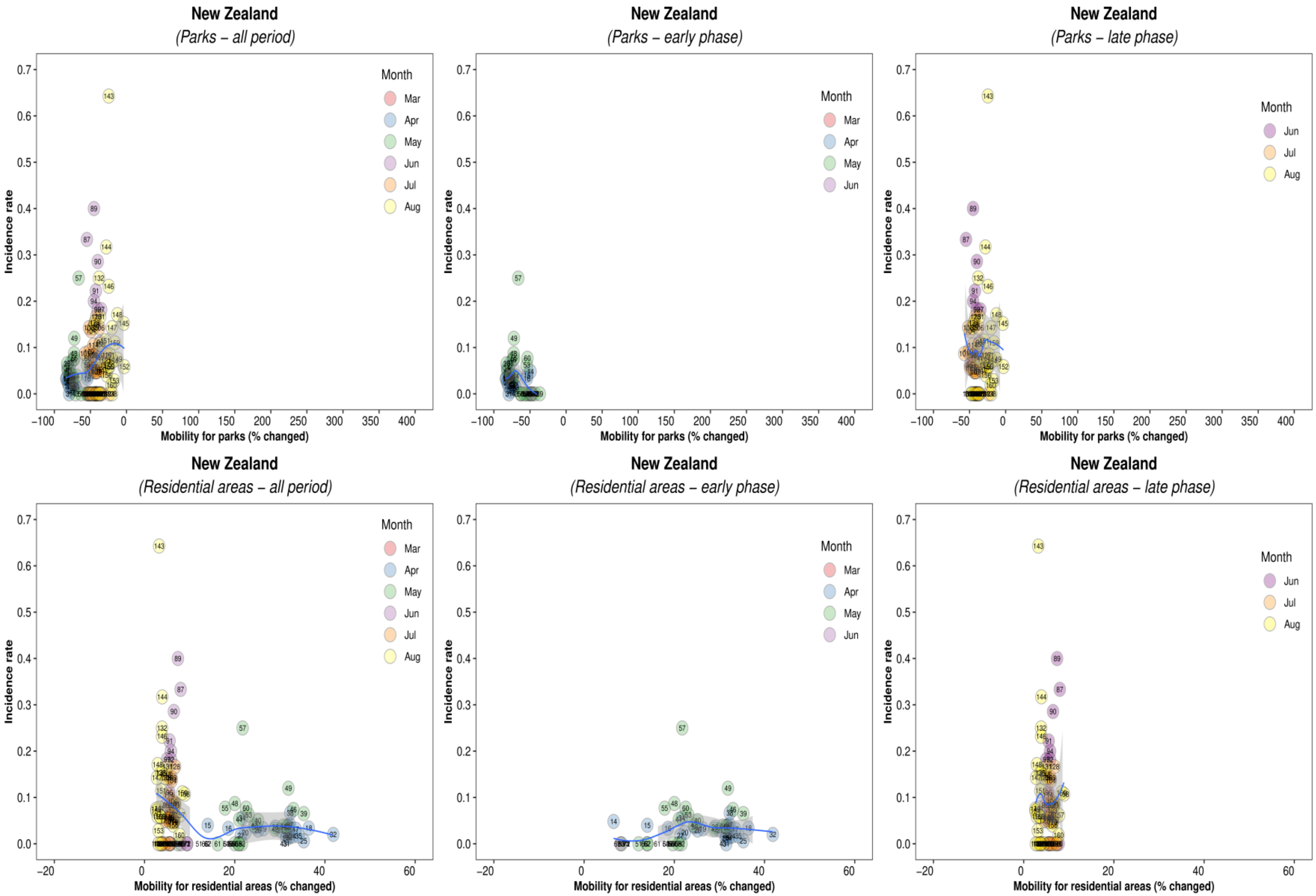
New Zealand
(Parks – all period)
New Zealand
(Parks – early phase)
New Zealand
(Parks – late phase)
New Zealand
(Residential areas – all period)
New Zealand
(Residential areas – early phase)
New Zealand
(Residential areas – late phase)
Incidence rate
Mobility for parks (% changed)
Mobility for residential areas (% changed)
Month
Mar
Apr
May
Jun
Jul
Aug

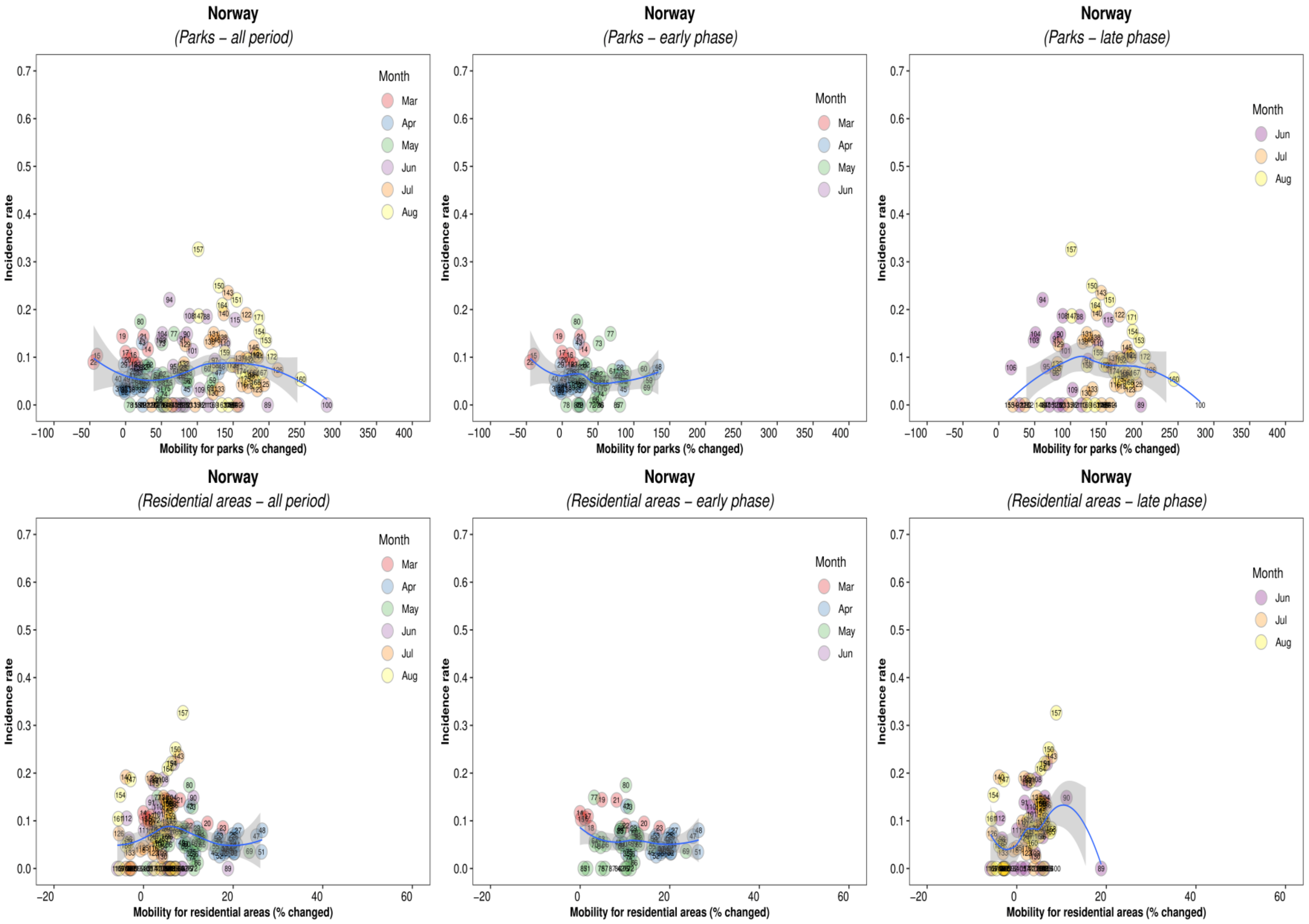

Norway
(Parks – all period)
Norway
(Parks – early phase)
Norway
(Parks – late phase)
Norway
(Residential areas – all period)
Norway
(Residential areas – early phase)
Norway
(Residential areas – late phase)
Incidence rate
Mobility for parks (% changed)
Mobility for residential areas (% changed)
Month
Mar
Apr
May
Jun
Jul
Aug

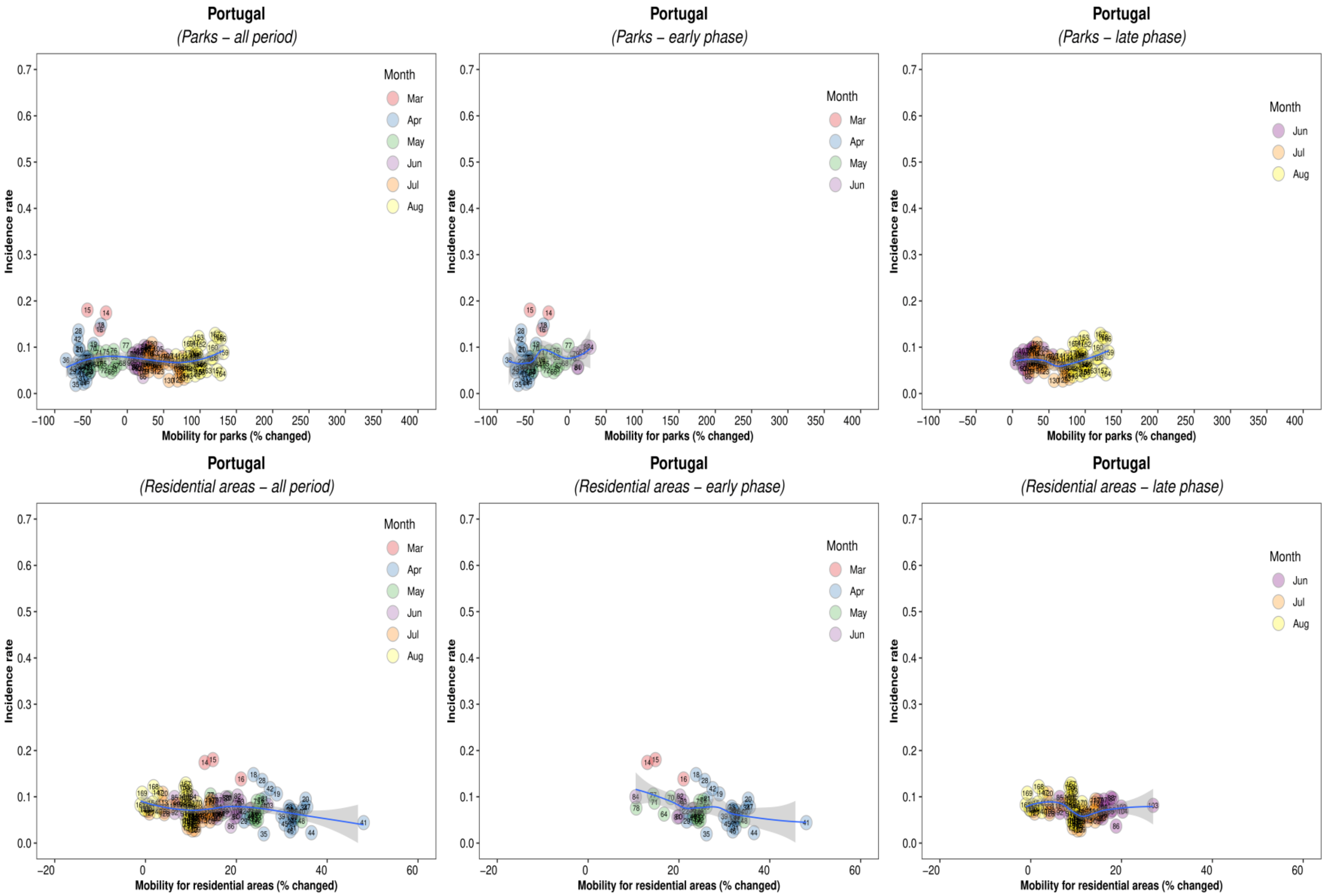
Portugal
(Parks – all period)
Portugal
(Parks – early phase)
Portugal
(Parks – late phase)
Portugal
(Residential areas – all period)
Portugal
(Residential areas – early phase)
Portugal
(Residential areas – late phase)
Incidence rate
Mobility for parks (% changed)
Mobility for residential areas (% changed)
Month
Mar
Apr
May
Jun
Jul
Aug

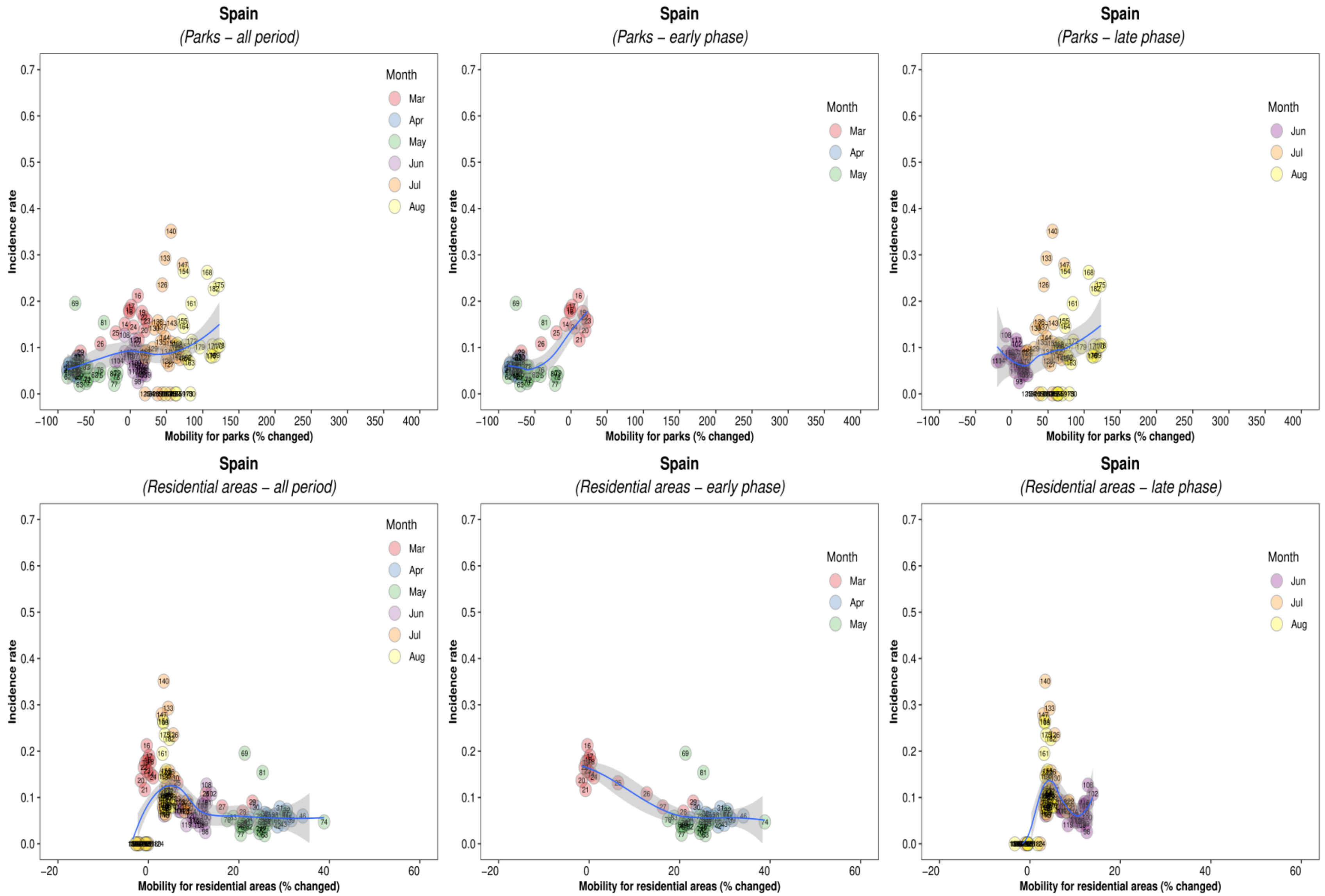
Spain
(Parks – all period)
Spain
(Parks – early phase)
Spain
(Parks – late phase)
Spain
(Residential areas – all period)
Spain
(Residential areas – early phase)
Spain
(Residential areas – late phase)
Incidence rate
Mobility for parks (% changed)
Mobility for residential areas (% changed)
Month
Mar
Apr
May
Jun
Jul
Aug

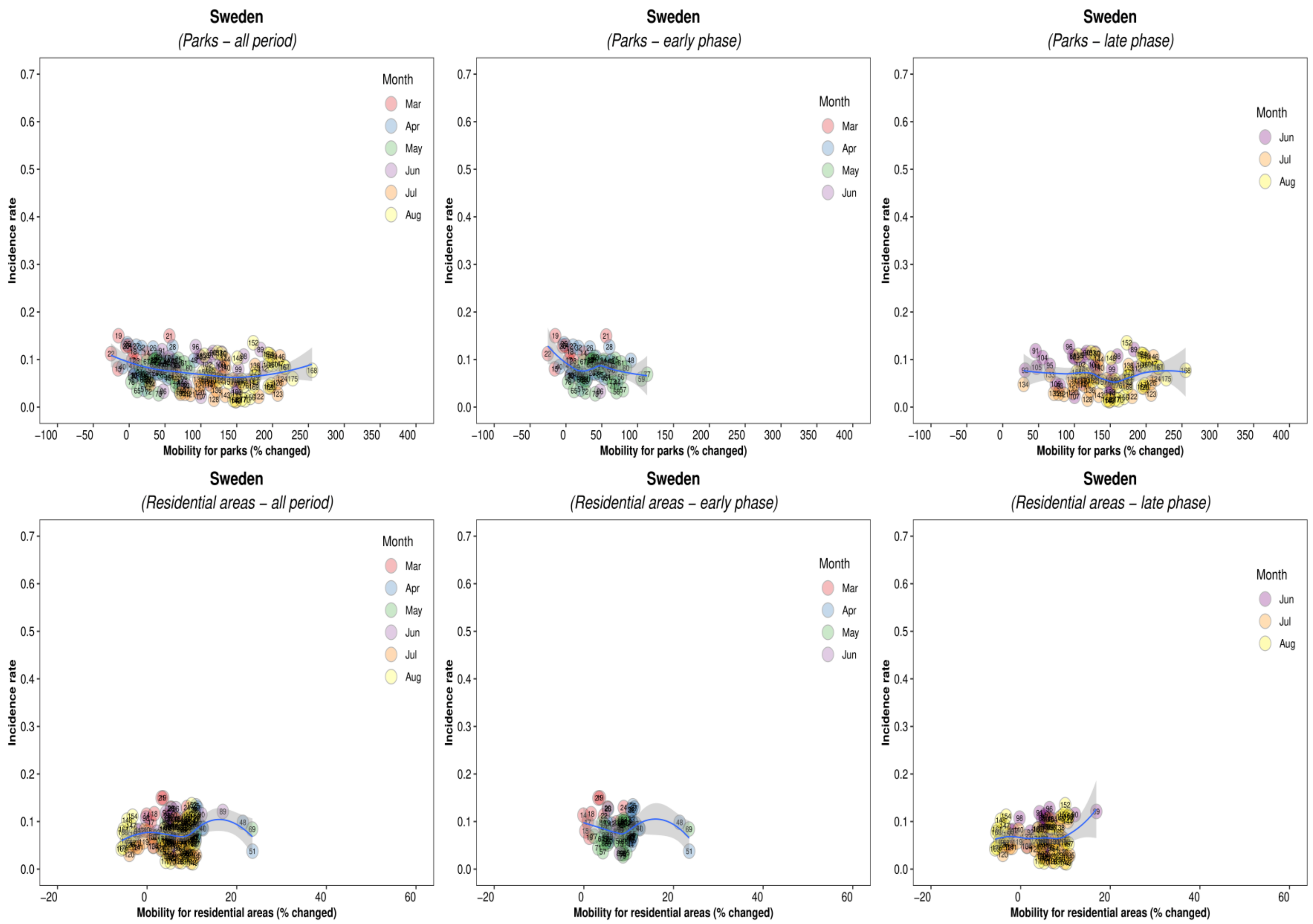

Sweden
(Parks – all period)
Sweden
(Parks – early phase)
Sweden
(Parks – late phase)
Sweden
(Residential areas – all period)
Sweden
(Residential areas – early phase)
Sweden
(Residential areas – late phase)
Incidence rate
Mobility for parks (% changed)
Mobility for residential areas (% changed)
Month
Mar
Apr
May
Jun
Jul
Aug

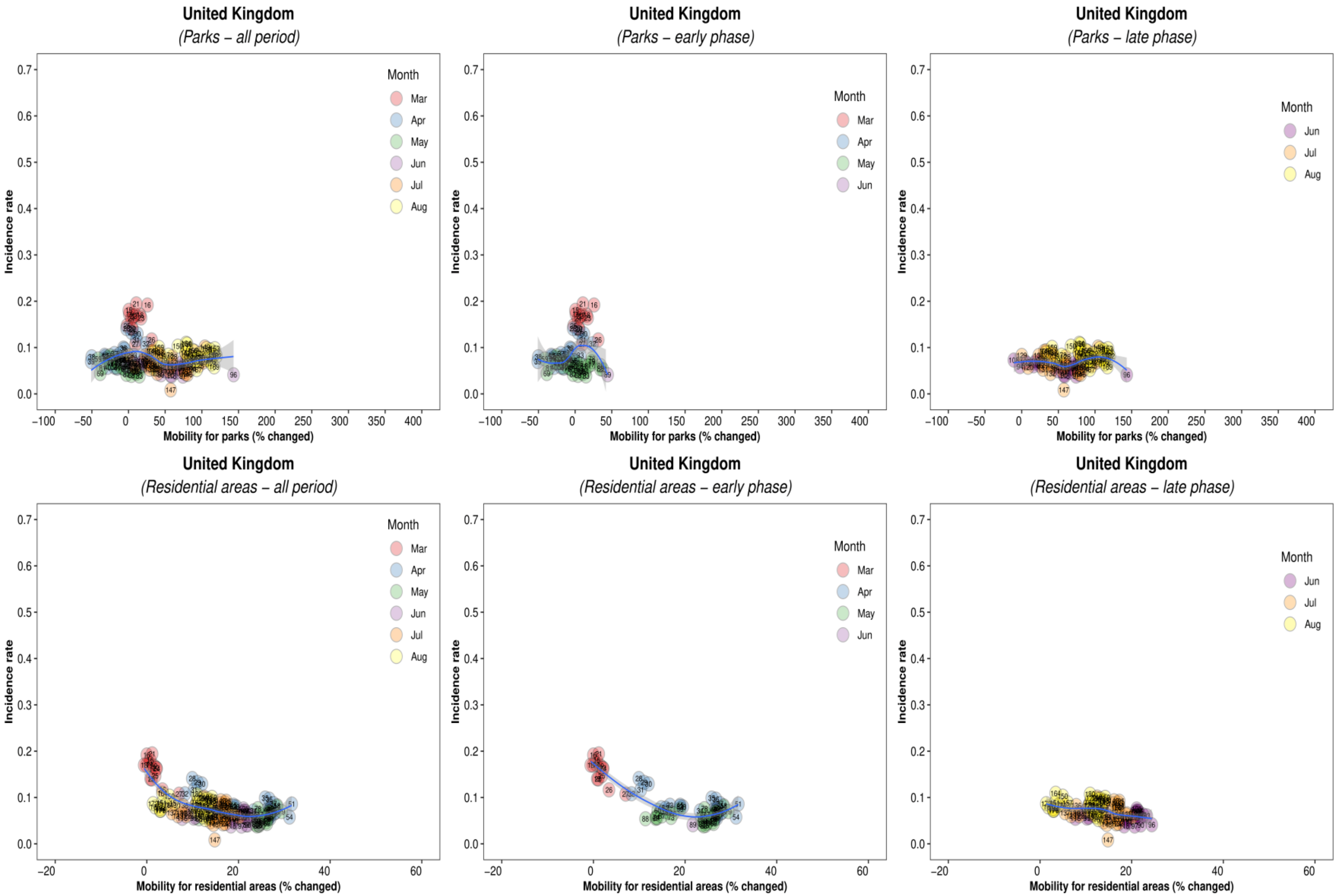
United Kingdom
(Parks – all period)
Incidence rate
Mobility for parks (% changed)
Month
Mar
Apr
May
Jun
Jul
Aug
United Kingdom
(Parks – early phase)
Incidence rate
Mobility for parks (% changed)
Month
Mar
Apr
May
Jun
United Kingdom
(Parks – late phase)
Incidence rate
Mobility for parks (% changed)
Month
Jun
Jul
Aug
United Kingdom
(Residential areas – all period)
Incidence rate
Mobility for residential areas (% changed)
Month
Mar
Apr
May
Jun
Jul
Aug
United Kingdom
(Residential areas – early phase)
Incidence rate
Mobility for residential areas (% changed)
Month
Mar
Apr
May
Jun
United Kingdom
(Residential areas – late phase)
Incidence rate
Mobility for residential areas (% changed)
Month
Jun
Jul
Aug

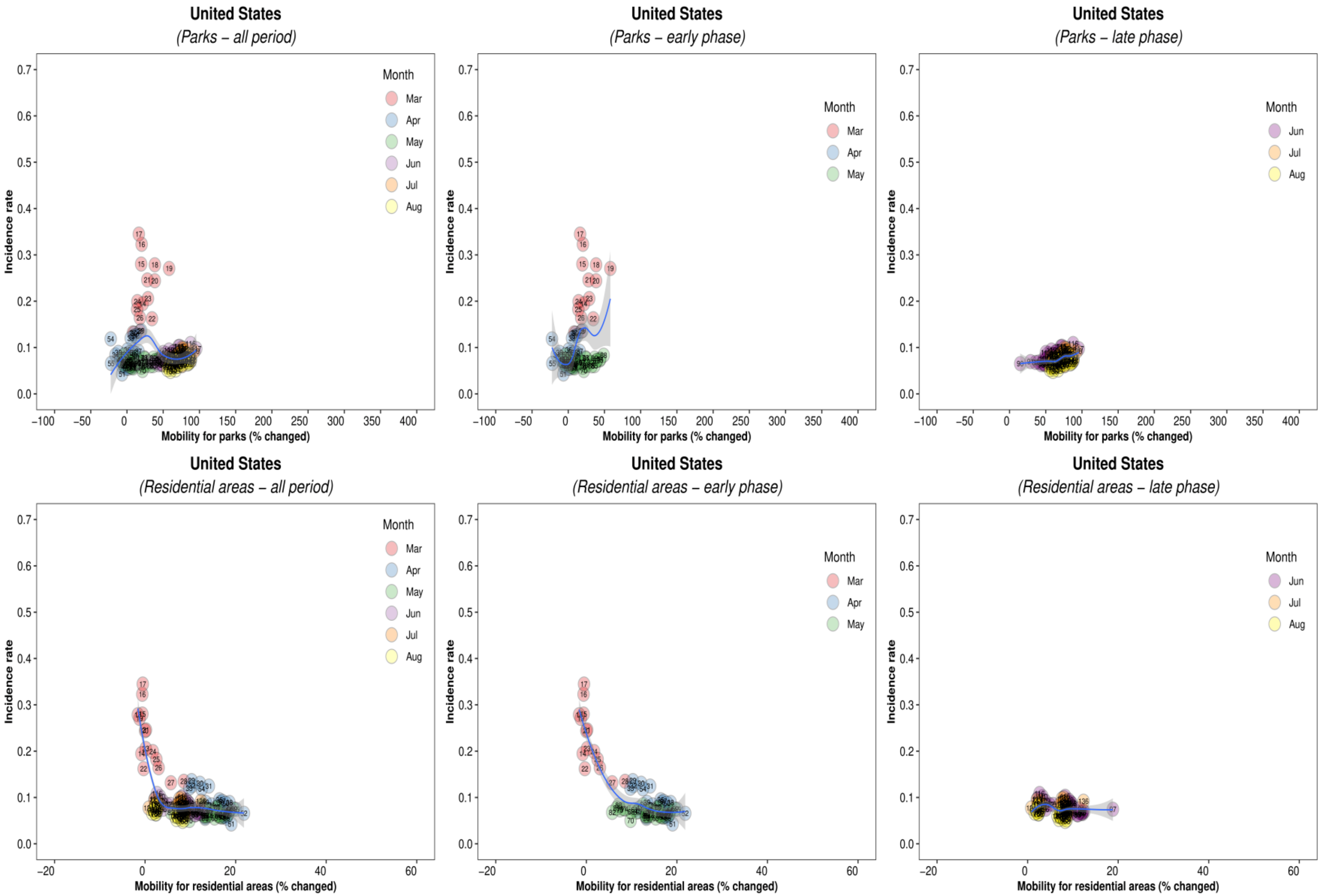
United States
(Parks – all period)
United States
(Parks – early phase)
United States
(Parks – late phase)
United States
(Residential areas – all period)
United States
(Residential areas – early phase)
United States
(Residential areas – late phase)
Incidence rate
Mobility for parks (% changed)
Mobility for residential areas (% changed)
Month
Mar
Apr
May
Jun
Jul
Aug

Supplementary Figure 2B. Association between new daily incidence rates of COVID-19 and mobility changes in 36 countries by early and late phase and other places (parks and residential areas) visited. Asia, Eastern Europe, Latin America, and Caribbean.

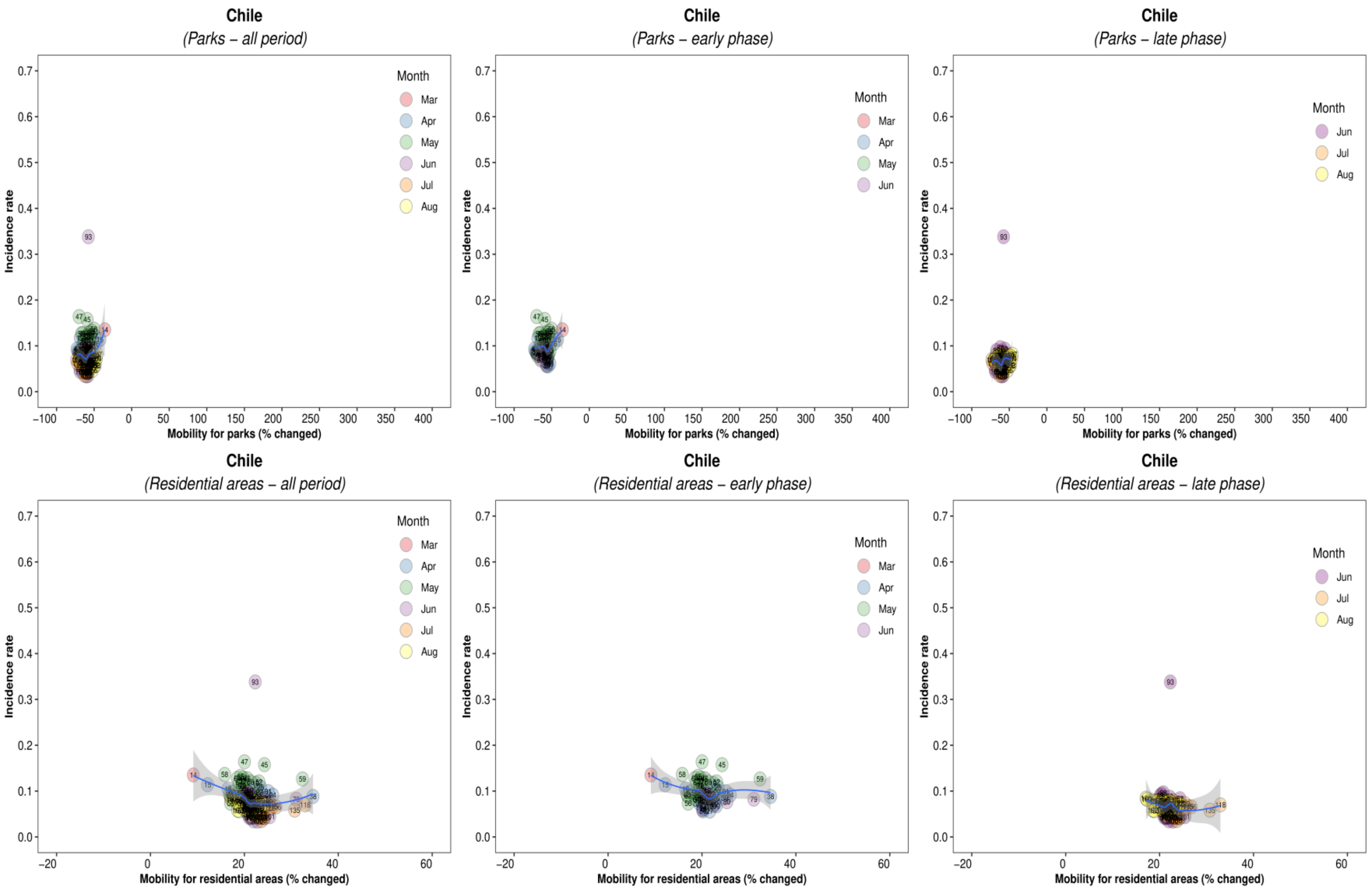

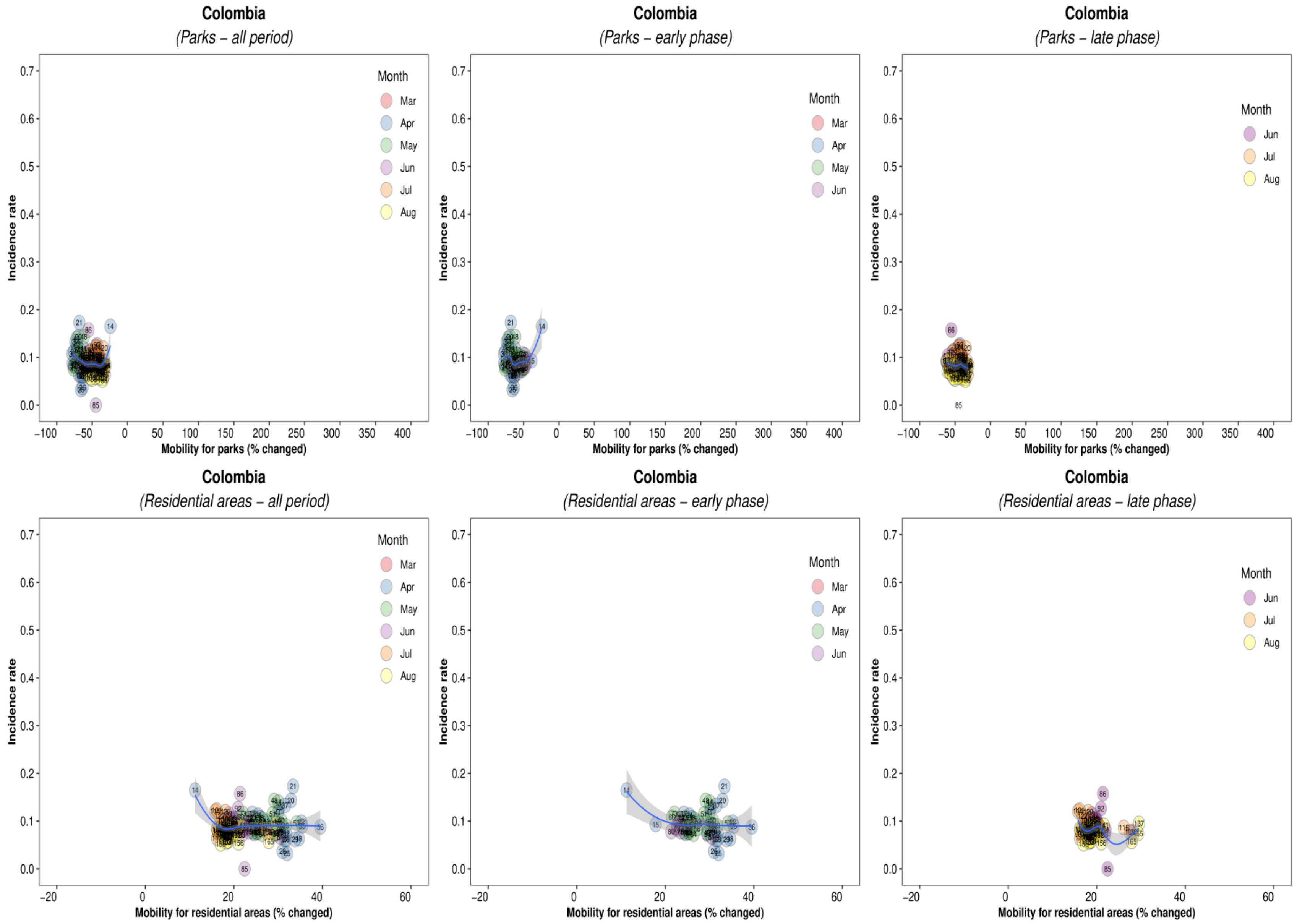
Colombia
(Parks – all period)
Colombia
(Parks – early phase)
Colombia
(Parks – late phase)
Incidence rate
Mobility for parks (% changed)
Month
Mar
Apr
May
Jun
Jul
Aug
Colombia
(Residential areas – all period)
Colombia
(Residential areas – early phase)
Colombia
(Residential areas – late phase)
Mobility for residential areas (% changed)

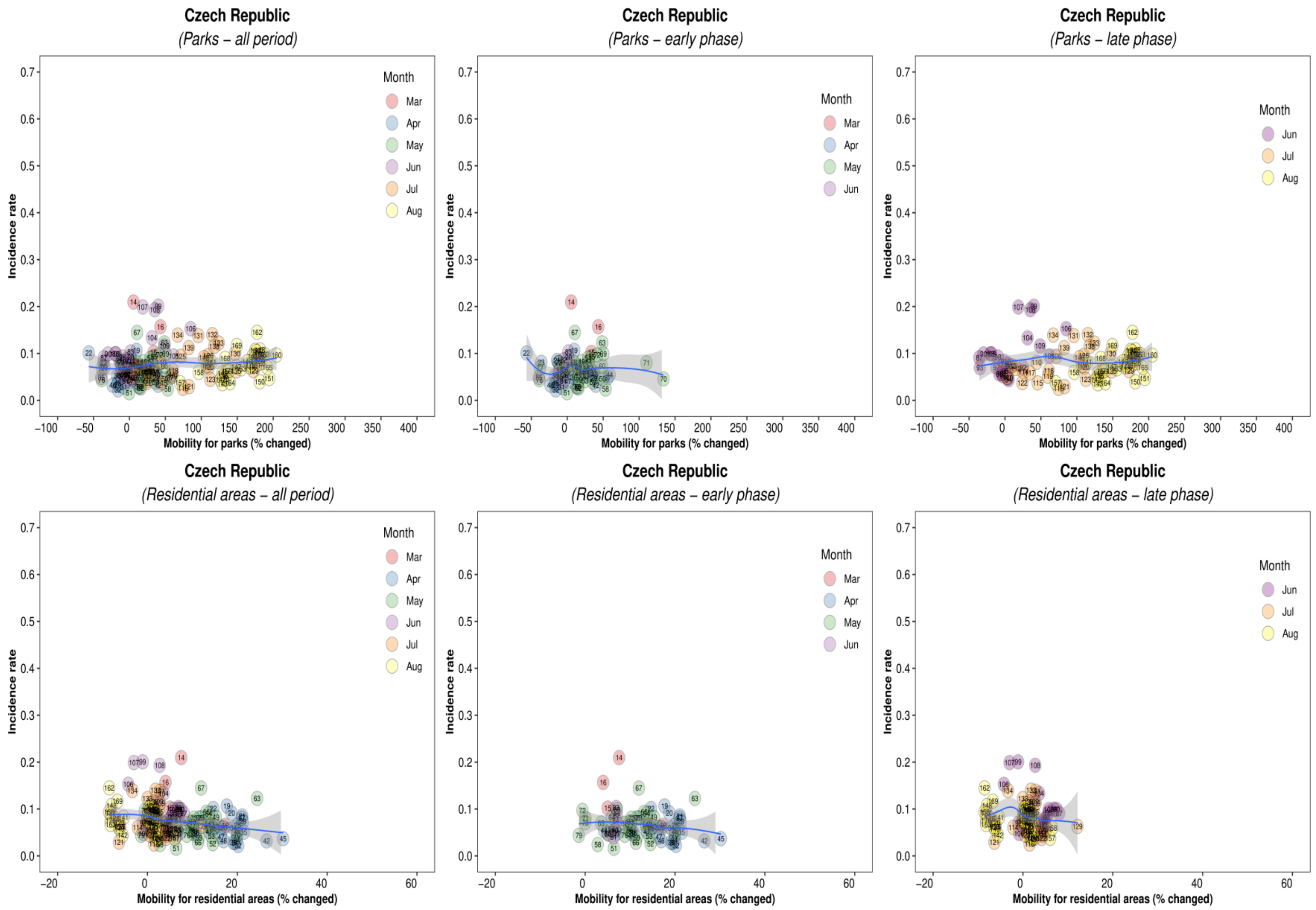
Czech Republic
(Parks – all period)
Czech Republic
(Parks – early phase)
Czech Republic
(Parks – late phase)
Czech Republic
(Residential areas – all period)
Czech Republic
(Residential areas – early phase)
Czech Republic
(Residential areas – late phase)
Incidence rate
Mobility for parks (% changed)
Mobility for residential areas (% changed)
Month
Mar
Apr
May
Jun
Jul
Aug
0.0
0.1
0.2
0.3
0.4
0.5
0.6
0.7
−100
−50
0
50
100
150
200
250
300
350
400
−20
0
20
40
60

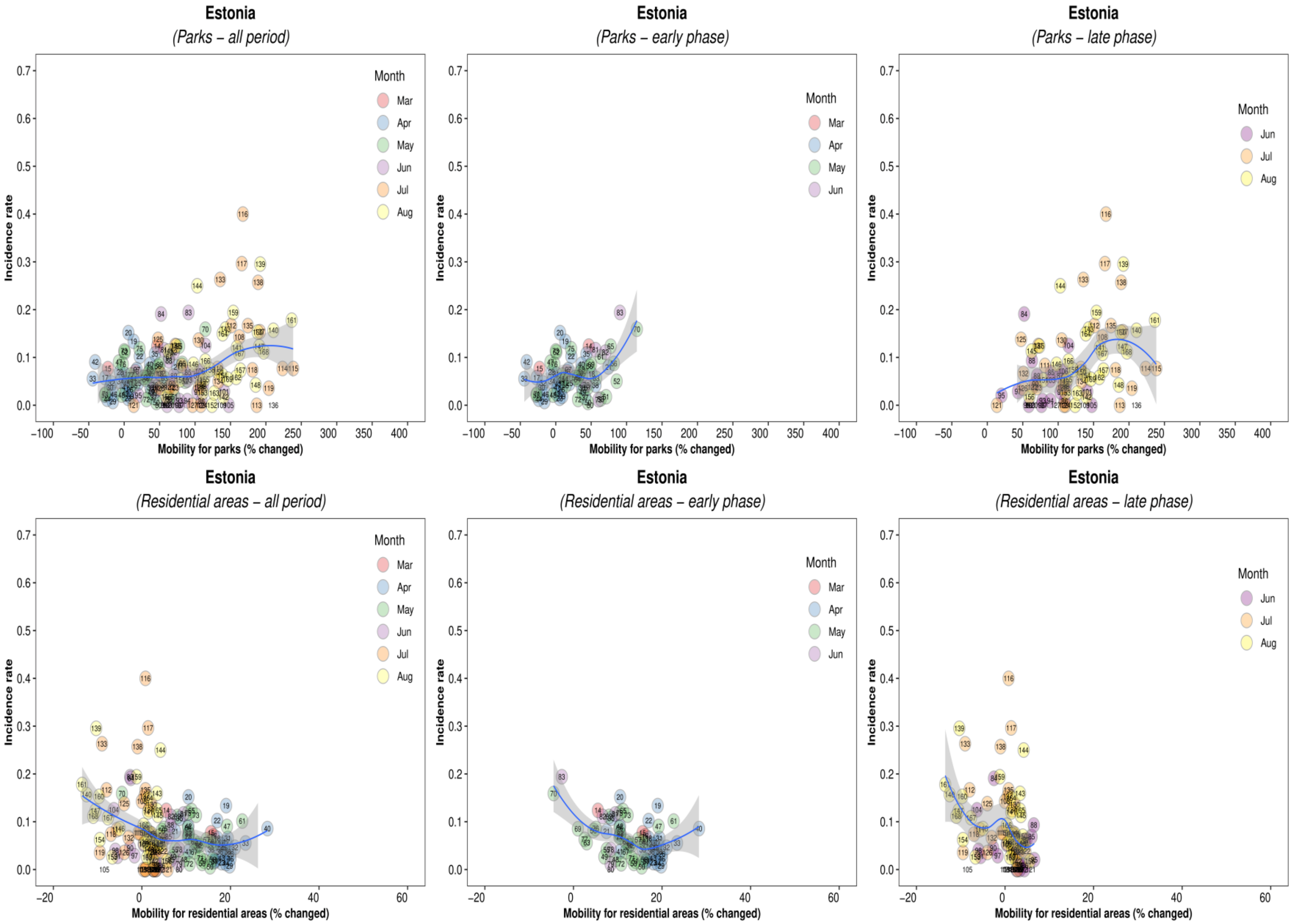
Estonia
(Parks – all period)
Estonia
(Parks – early phase)
Estonia
(Parks – late phase)
Estonia
(Residential areas – all period)
Estonia
(Residential areas – early phase)
Estonia
(Residential areas – late phase)
Incidence rate
Mobility for parks (% changed)
Mobility for residential areas (% changed)
Month
Mar
Apr
May
Jun
Jul
Aug

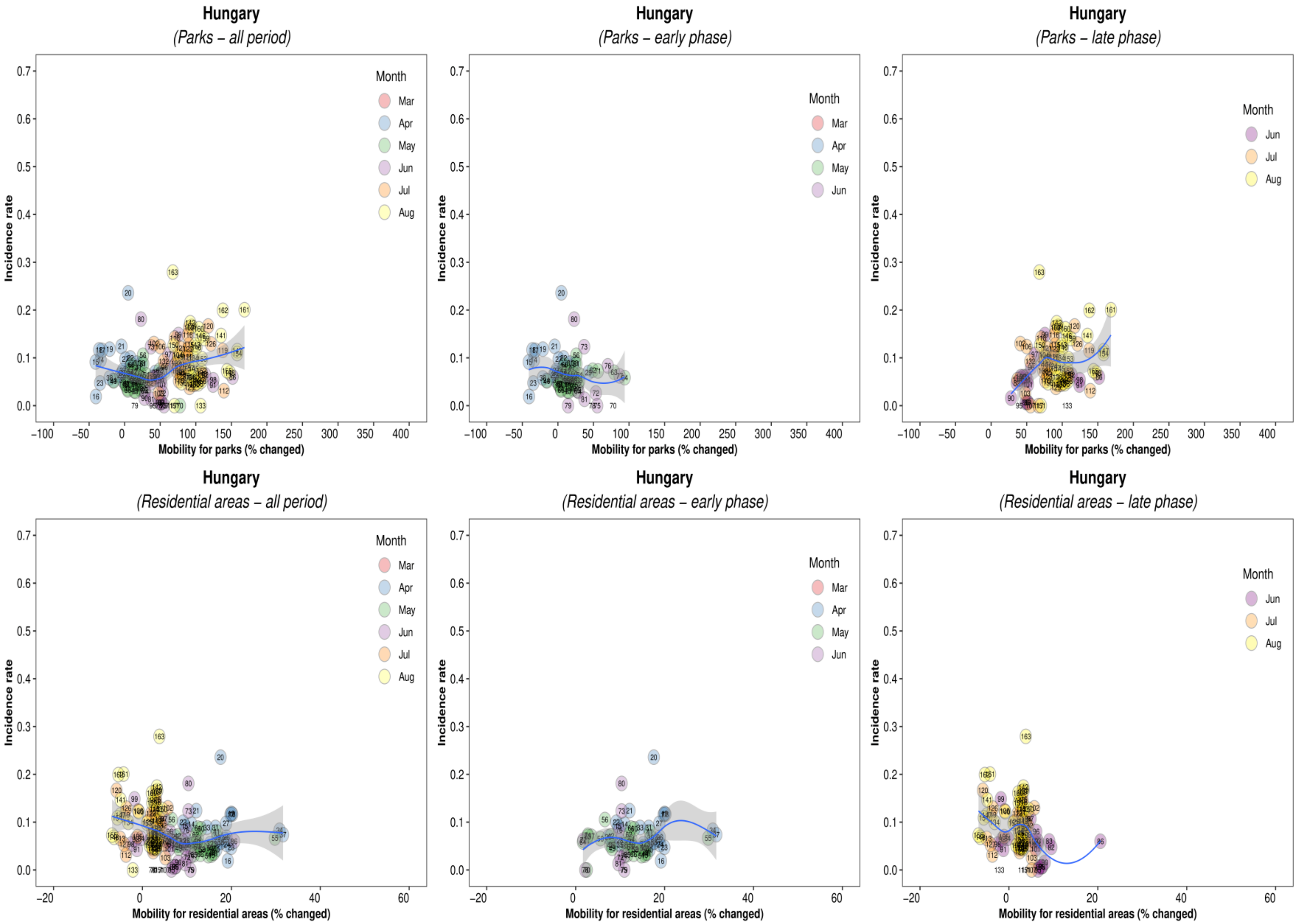
Hungary
(Parks – all period)
Hungary
(Parks – early phase)
Hungary
(Parks – late phase)
Hungary
(Residential areas – all period)
Hungary
(Residential areas – early phase)
Hungary
(Residential areas – late phase)
Incidence rate
Mobility for parks (% changed)
Mobility for residential areas (% changed)
Month
Mar
Apr
May
Jun
Jul
Aug

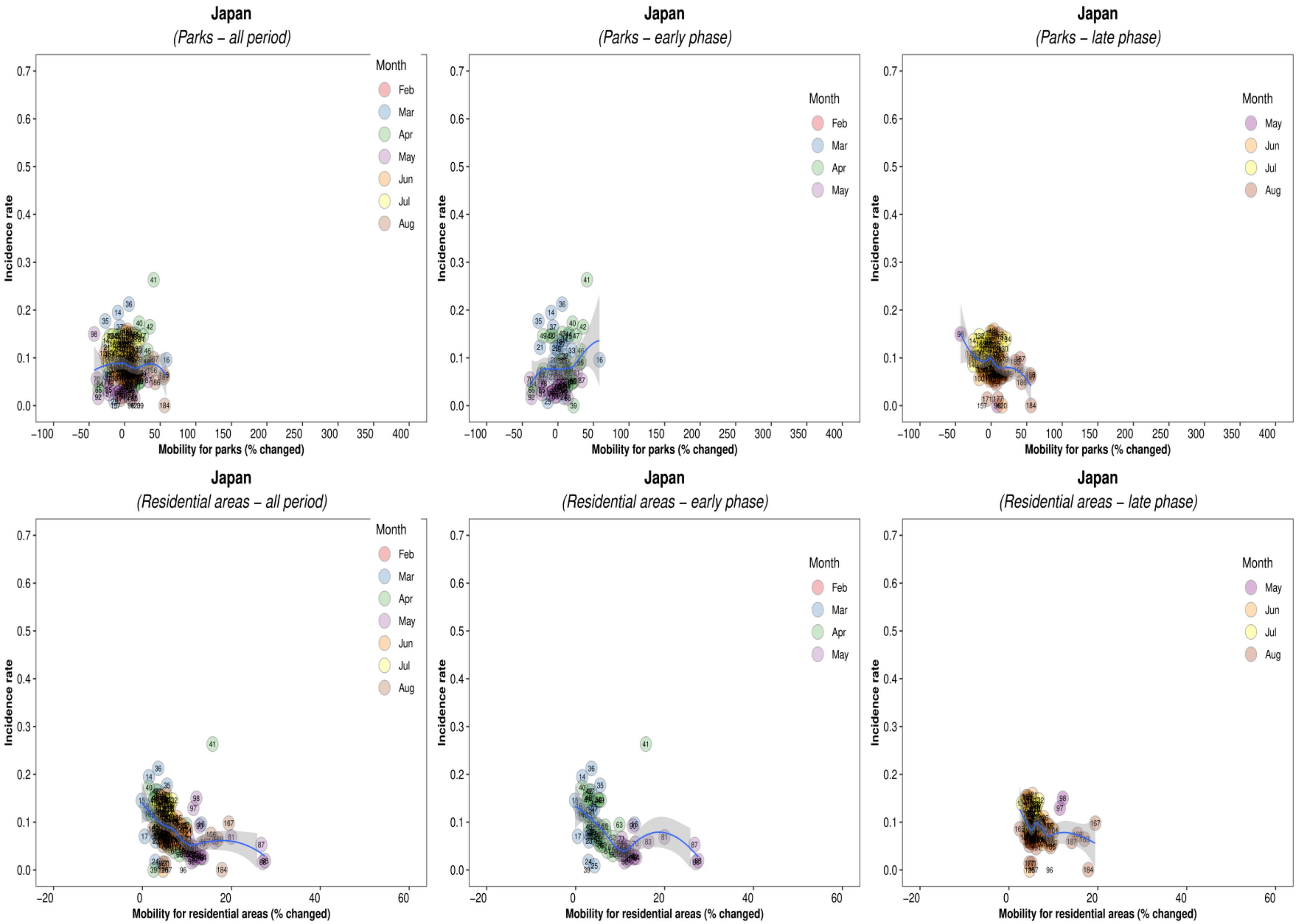
Japan
(Parks – all period)
Incidence rate
Mobility for parks (% changed)
Month
Feb
Mar
Apr
May
Jun
Jul
Aug
Japan
(Parks – early phase)
Incidence rate
Mobility for parks (% changed)
Month
Feb
Mar
Apr
May
Japan
(Parks – late phase)
Incidence rate
Mobility for parks (% changed)
Month
May
Jun
Jul
Aug
Japan
(Residential areas – all period)
Incidence rate
Mobility for residential areas (% changed)
Month
Feb
Mar
Apr
May
Jun
Jul
Aug
Japan
(Residential areas – early phase)
Incidence rate
Mobility for residential areas (% changed)
Month
Feb
Mar
Apr
May
Japan
(Residential areas – late phase)
Incidence rate
Mobility for residential areas (% changed)
Month
May
Jun
Jul
Aug

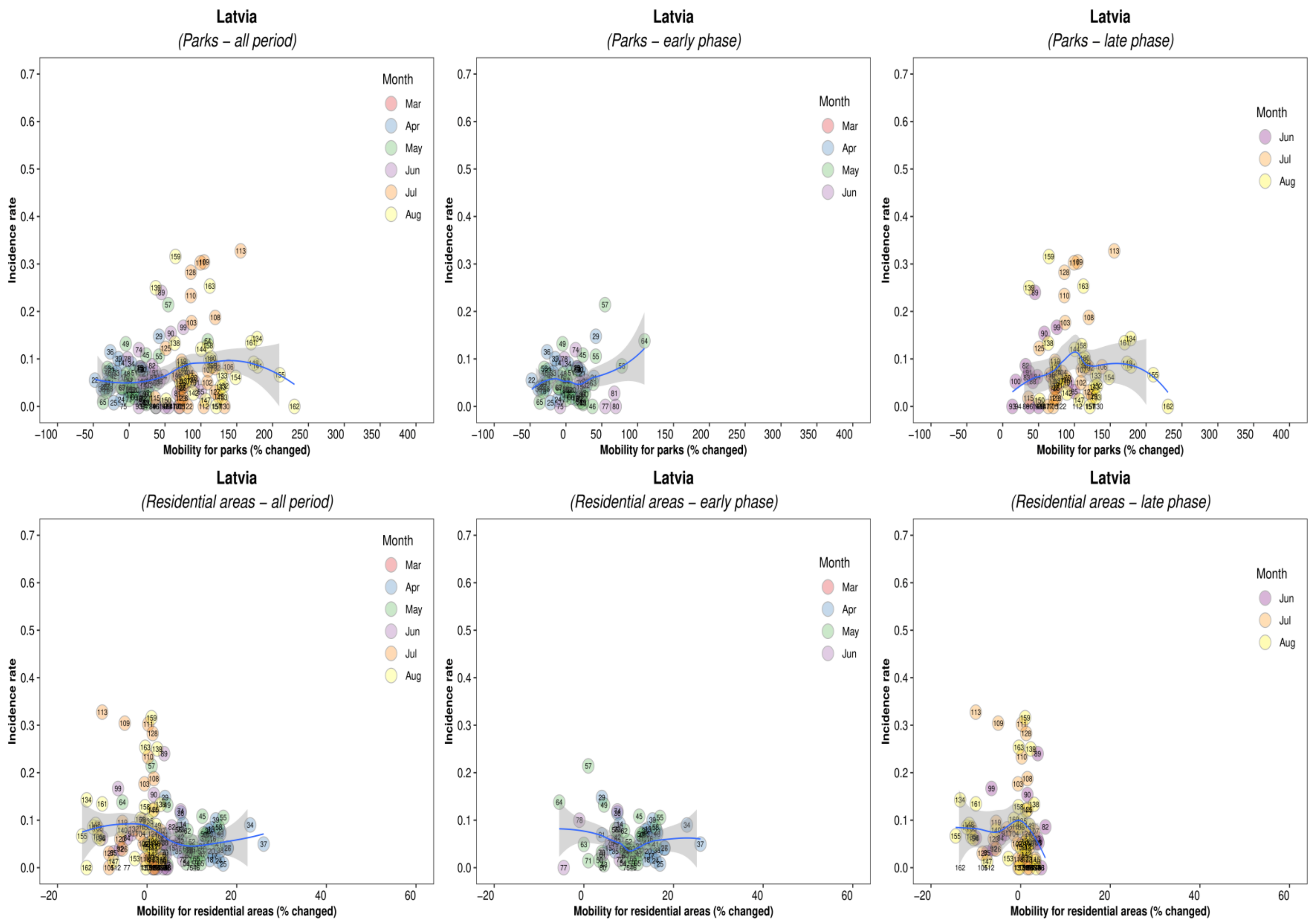
Latvia
(Parks – all period)
Latvia
(Parks – early phase)
Latvia
(Parks – late phase)
Latvia
(Residential areas – all period)
Latvia
(Residential areas – early phase)
Latvia
(Residential areas – late phase)
Incidence rate
Mobility for parks (% changed)
Mobility for residential areas (% changed)
Month
Mar
Apr
May
Jun
Jul
Aug

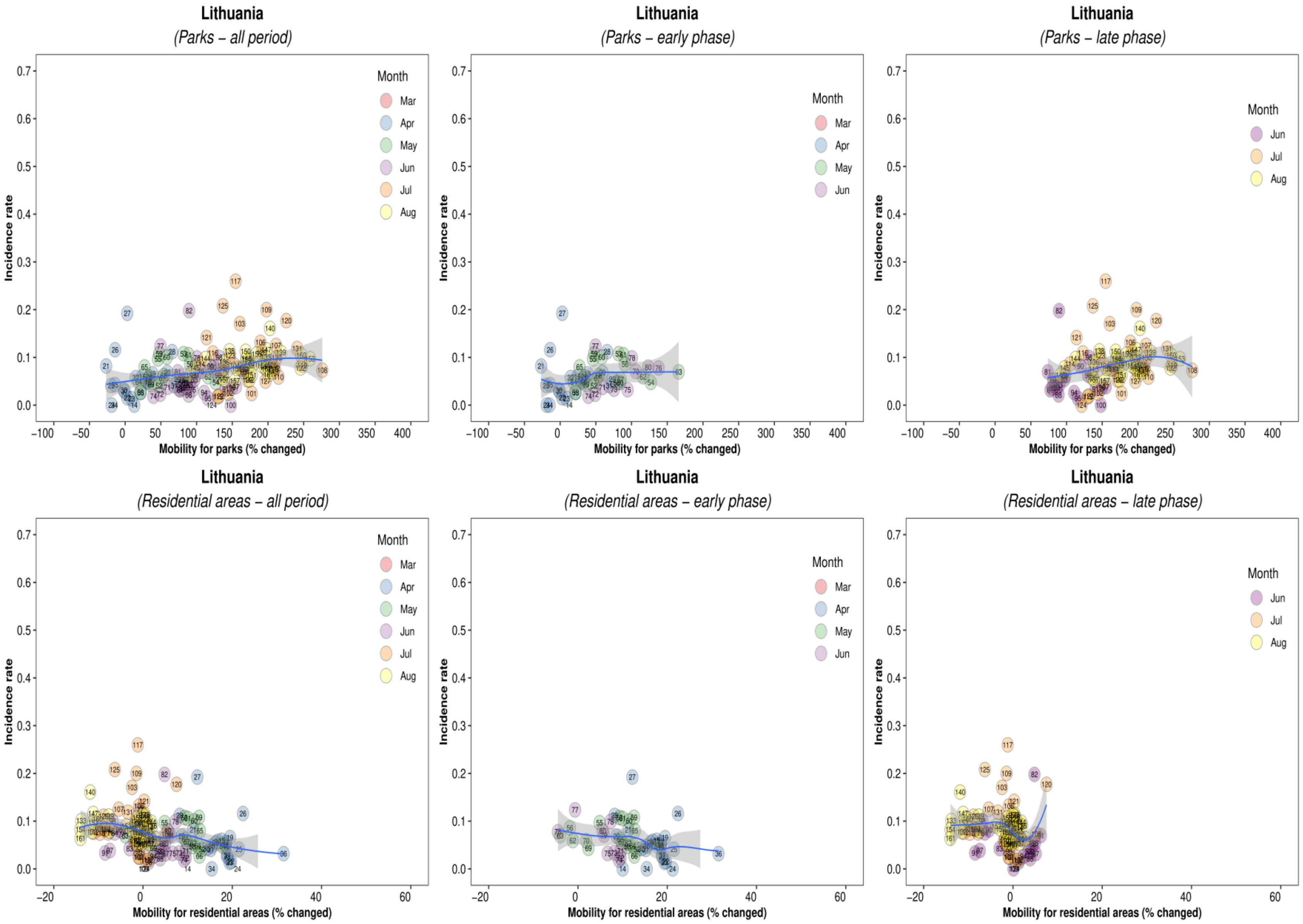
Lithuania
(Parks – all period)
Lithuania
(Parks – early phase)
Lithuania
(Parks – late phase)
Lithuania
(Residential areas – all period)
Lithuania
(Residential areas – early phase)
Lithuania
(Residential areas – late phase)
Incidence rate
Mobility for parks (% changed)
Mobility for residential areas (% changed)
Month
Mar
Apr
May
Jun
Jul
Aug

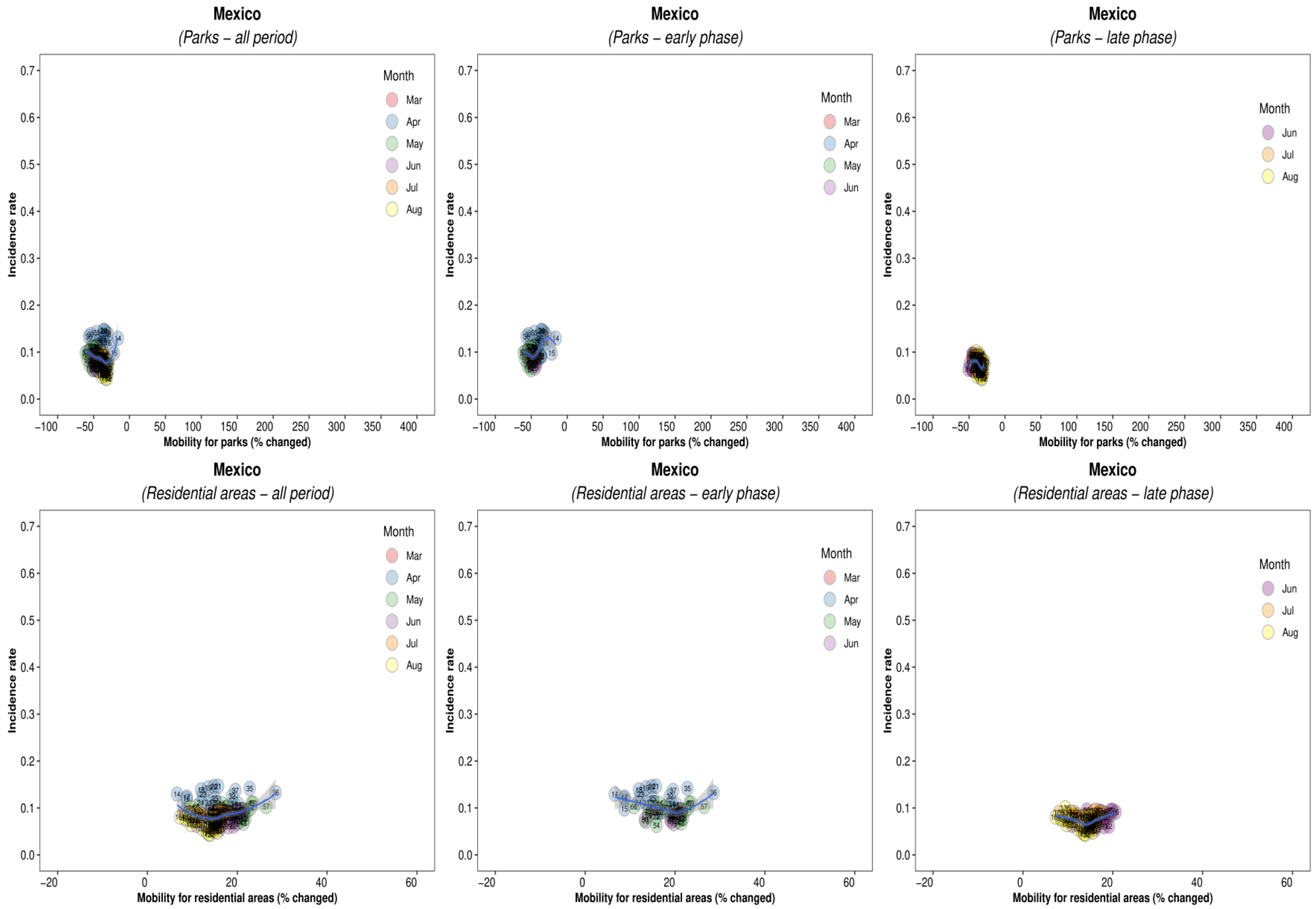
Mexico
(Parks – all period)
Mexico
(Parks – early phase)
Mexico
(Parks – late phase)
Mexico
(Residential areas – all period)
Mexico
(Residential areas – early phase)
Mexico
(Residential areas – late phase)
Incidence rate
Mobility for parks (% changed)
Mobility for residential areas (% changed)
Month
Mar
Apr
May
Jun
Jul
Aug

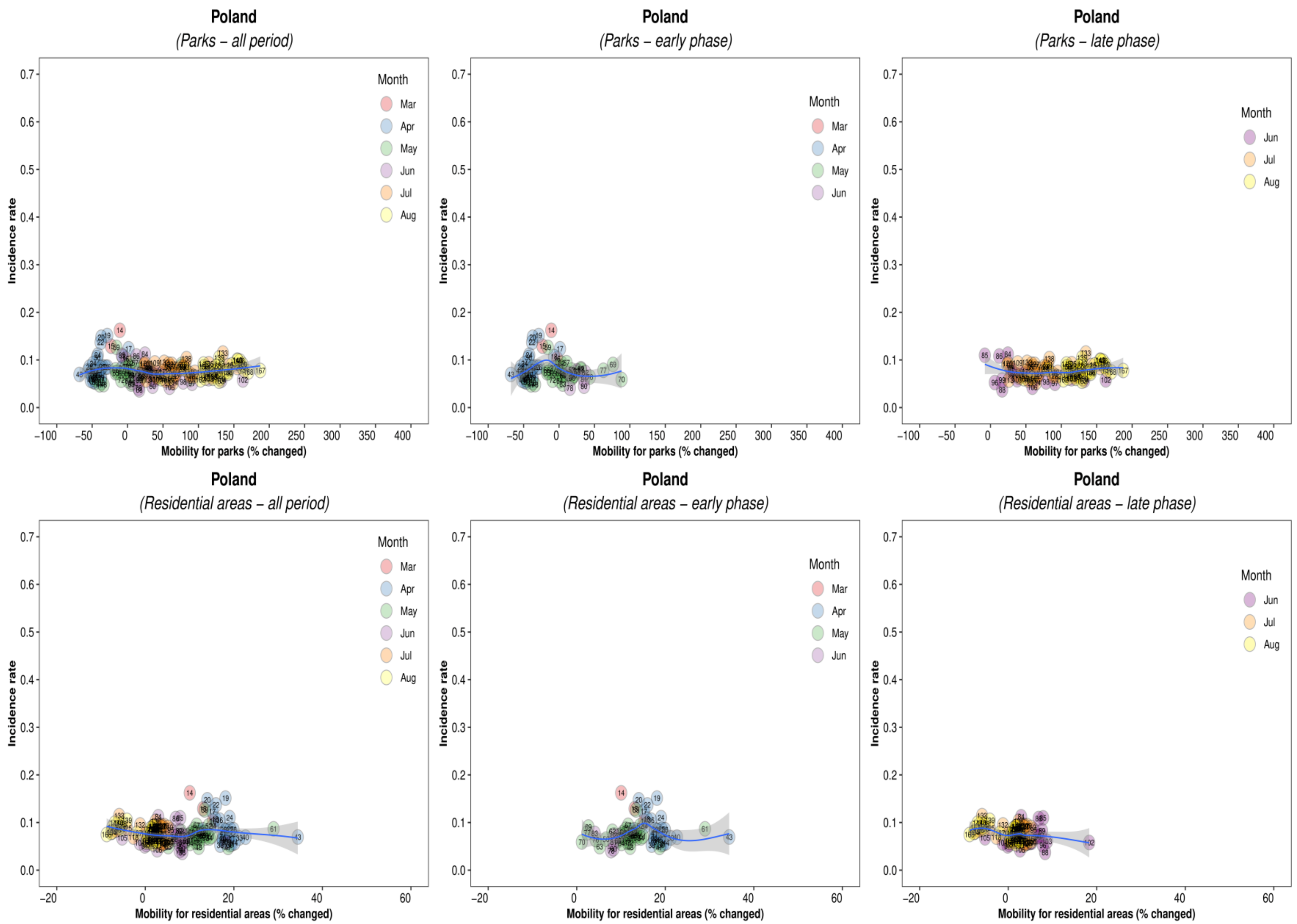
Poland
(Parks – all period)
Poland
(Parks – early phase)
Poland
(Parks – late phase)
Incidence rate
Mobility for parks (% changed)
Month
Mar
Apr
May
Jun
Jul
Aug
Poland
(Residential areas – all period)
Poland
(Residential areas – early phase)
Poland
(Residential areas – late phase)
Mobility for residential areas (% changed)

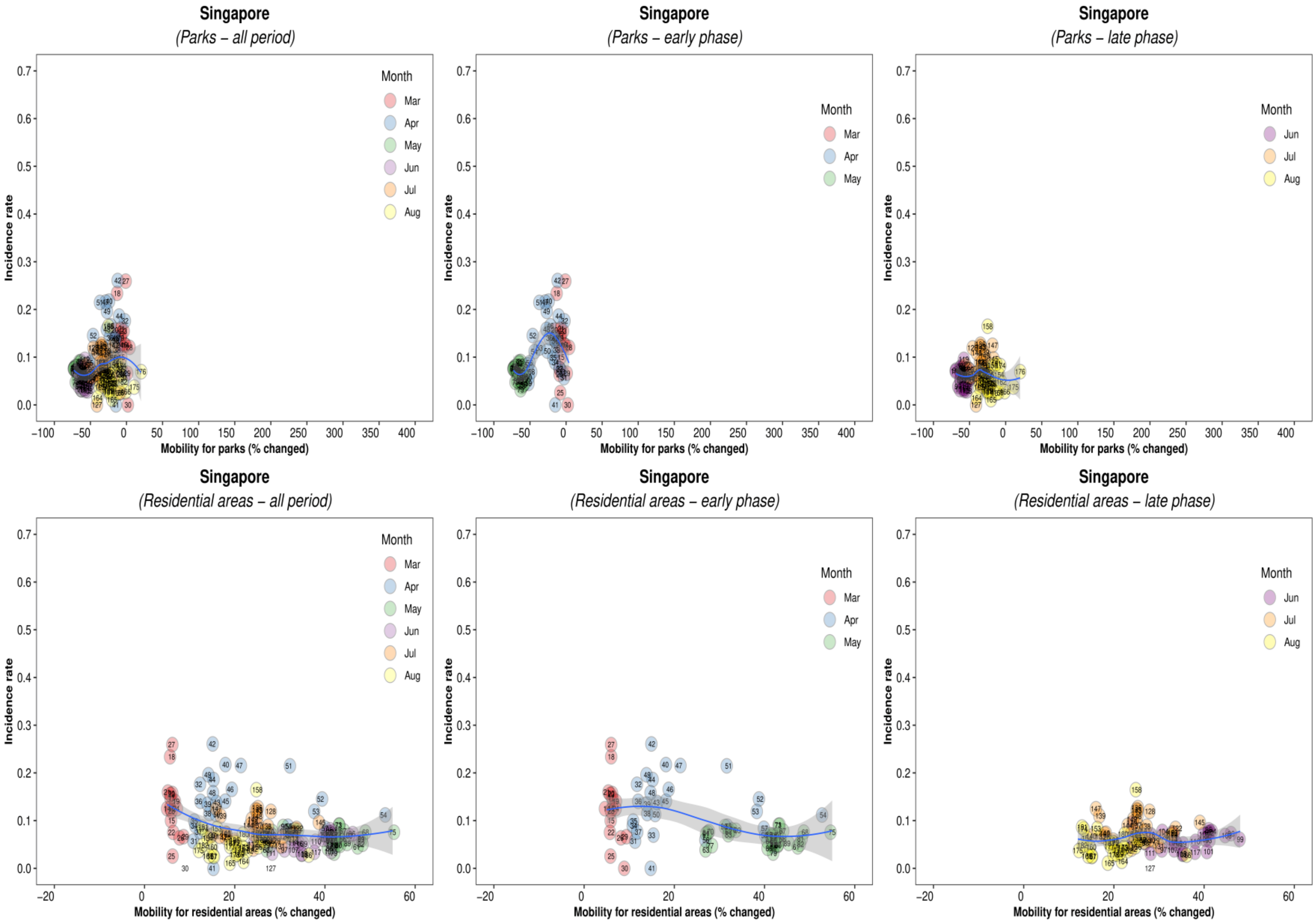

Singapore
(Parks – all period)
Month
Mar
Apr
May
Jun
Jul
Aug
Incidence rate
Mobility for parks (% changed)
Singapore
(Parks – early phase)
Singapore
(Parks – late phase)
Singapore
(Residential areas – all period)
Mobility for residential areas (% changed)
Singapore
(Residential areas – early phase)
Singapore
(Residential areas – late phase)

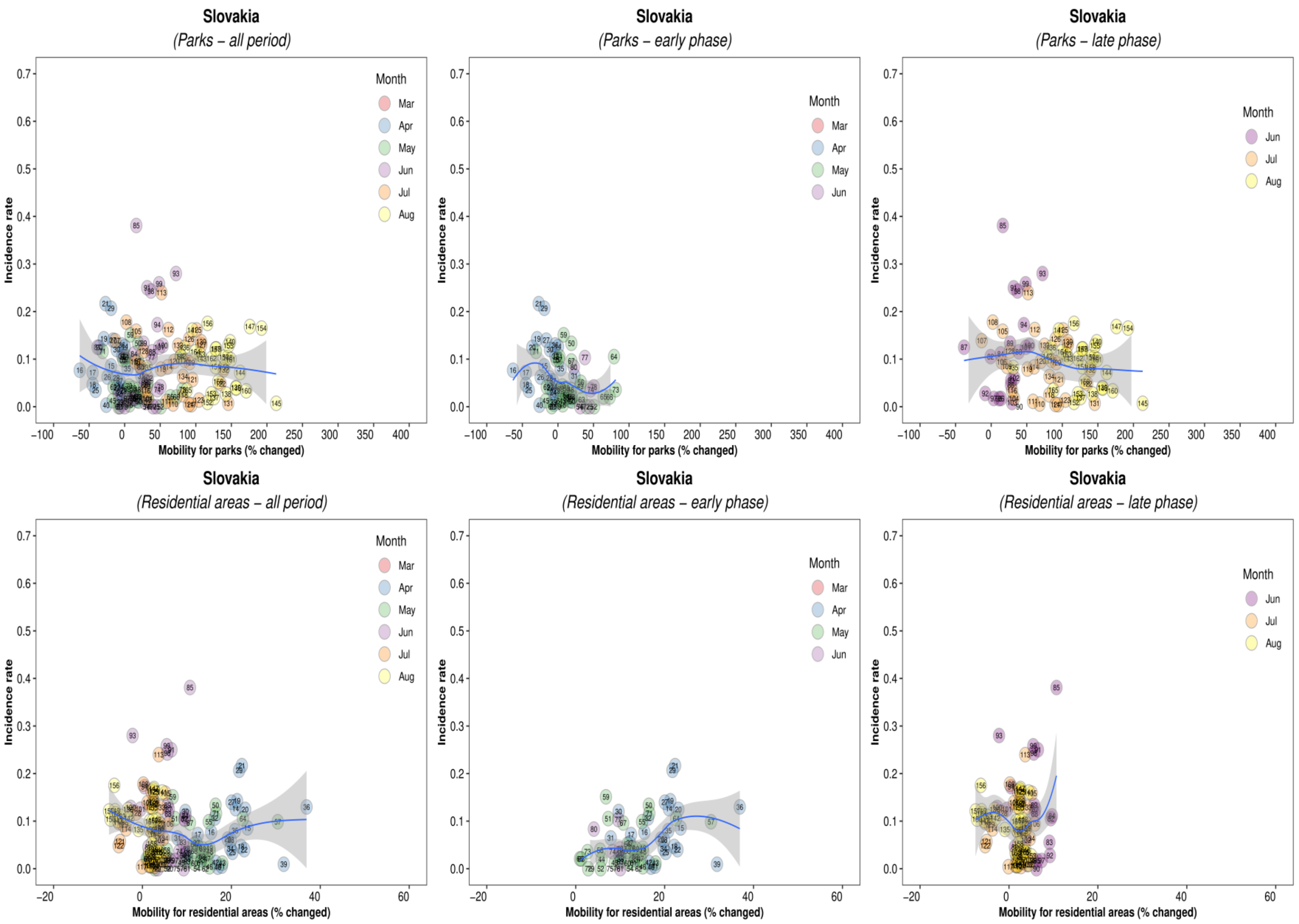
Slovakia
(Parks – all period)
Slovakia
(Parks – early phase)
Slovakia
(Parks – late phase)
Slovakia
(Residential areas – all period)
Slovakia
(Residential areas – early phase)
Slovakia
(Residential areas – late phase)
Incidence rate
Mobility for parks (% changed)
Mobility for residential areas (% changed)
Month
Mar
Apr
May
Jun
Jul
Aug

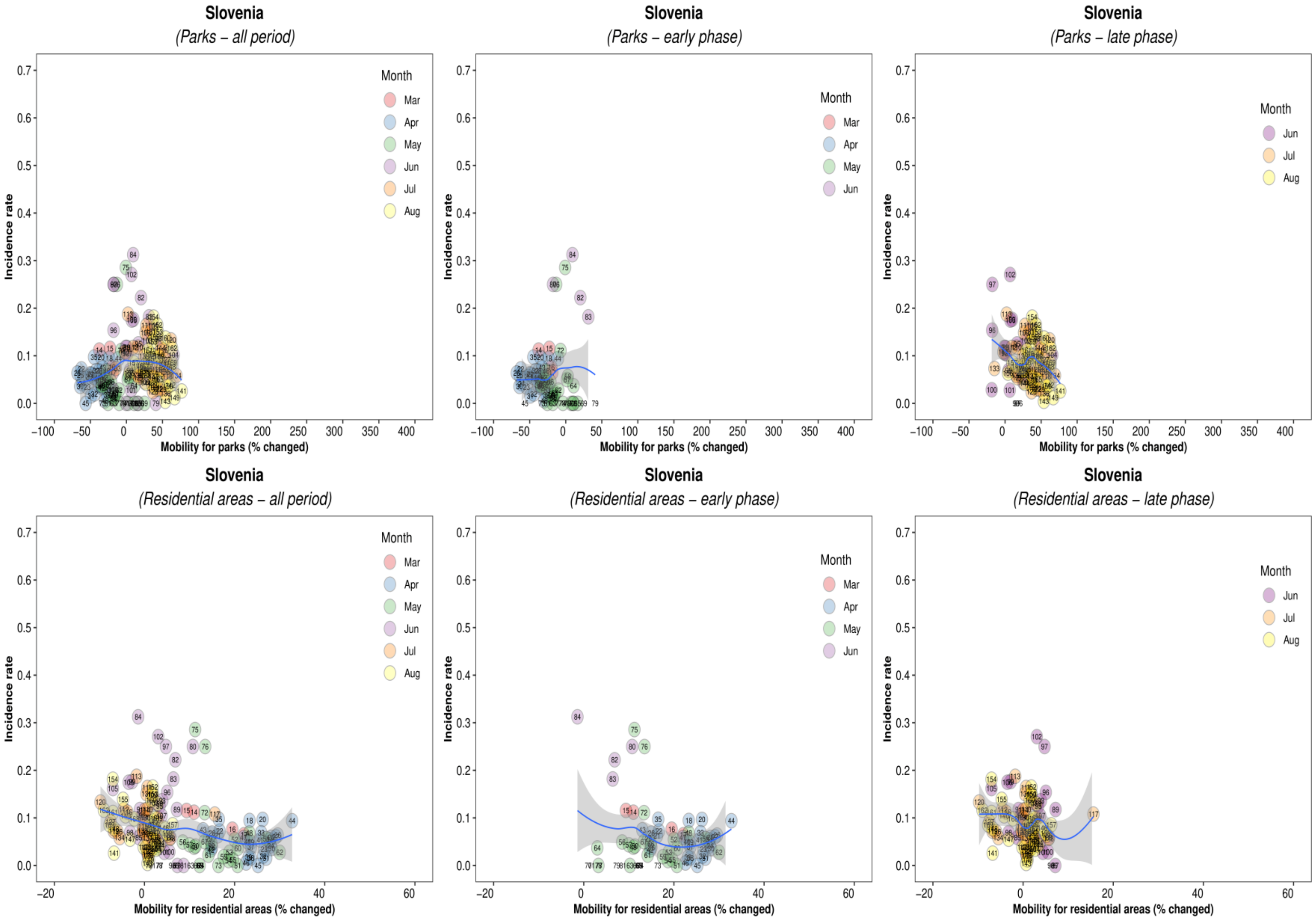
Slovenia
(Parks – all period)
Slovenia
(Parks – early phase)
Slovenia
(Parks – late phase)
Slovenia
(Residential areas – all period)
Slovenia
(Residential areas – early phase)
Slovenia
(Residential areas – late phase)
Incidence rate
Mobility for parks (% changed)
Mobility for residential areas (% changed)
Month
Mar
Apr
May
Jun
Jul
Aug

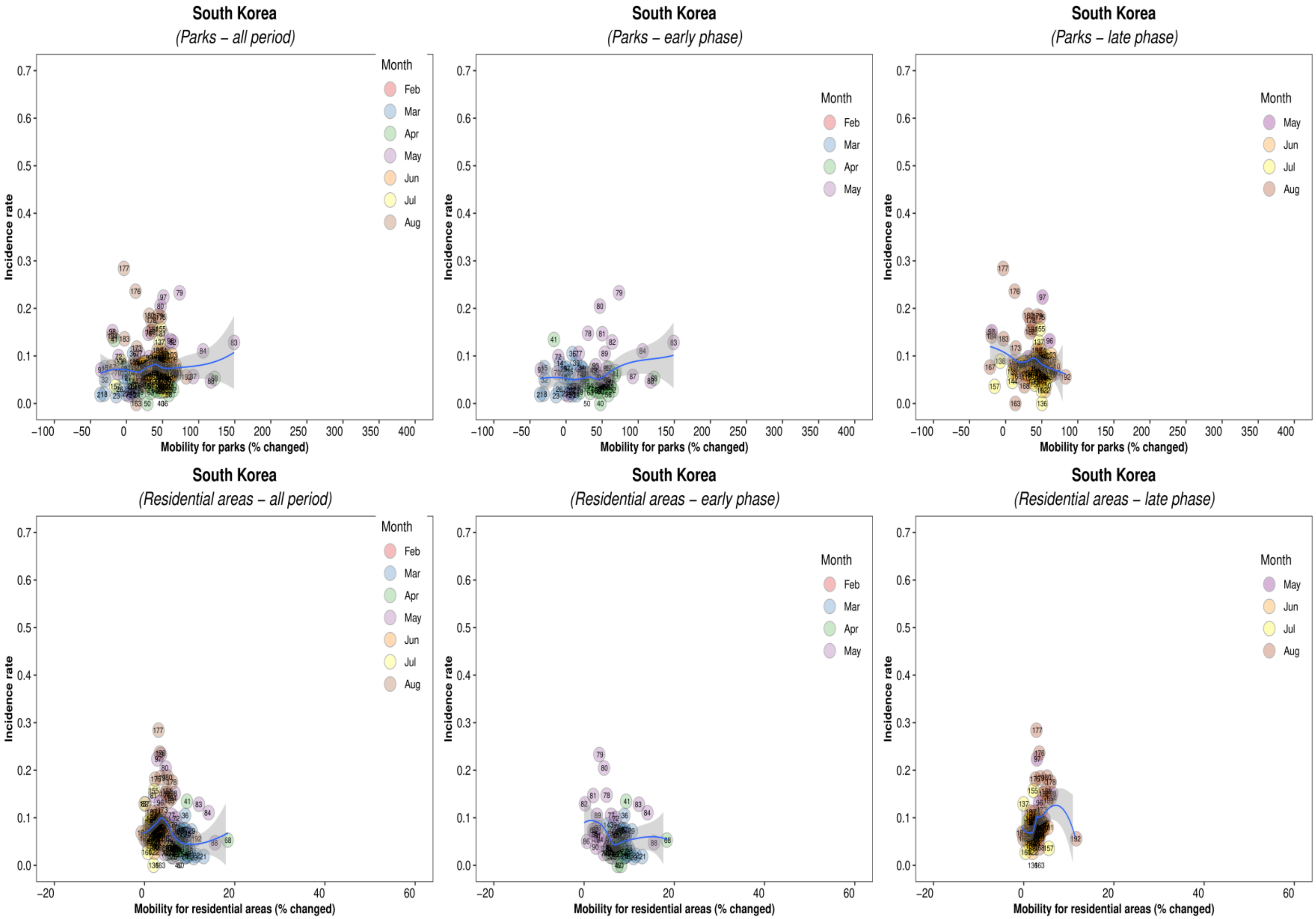
South Korea
(Parks – all period)
South Korea
(Parks – early phase)
South Korea
(Parks – late phase)
Incidence rate
Mobility for parks (% changed)
Month
Feb
Mar
Apr
May
Jun
Jul
Aug
South Korea
(Residential areas – all period)
South Korea
(Residential areas – early phase)
South Korea
(Residential areas – late phase)
Mobility for residential areas (% changed)

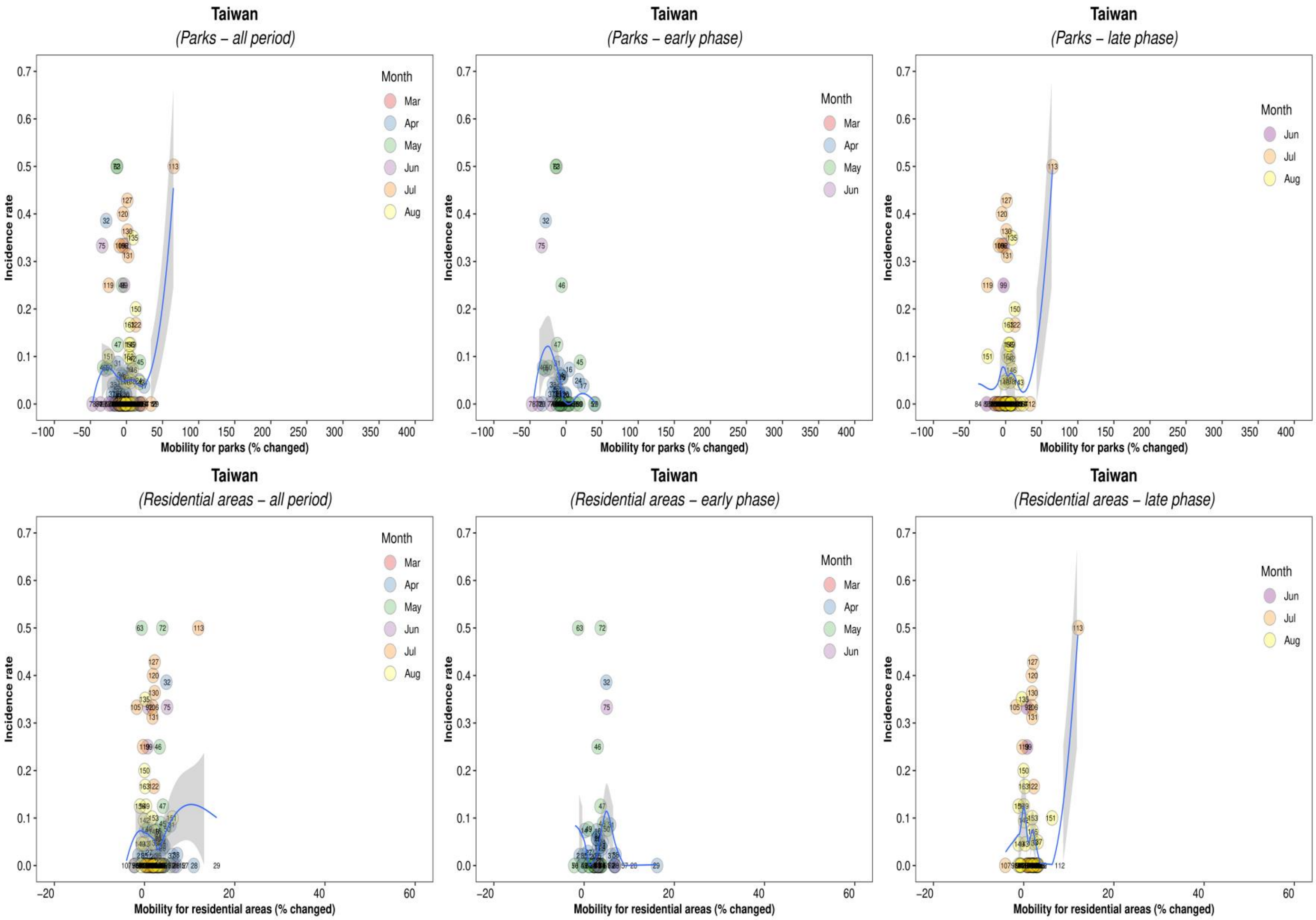

Taiwan
(Parks – all period)
Taiwan
(Parks – early phase)
Taiwan
(Parks – late phase)
Taiwan
(Residential areas – all period)
Taiwan
(Residential areas – early phase)
Taiwan
(Residential areas – late phase)
Incidence rate
Mobility for parks (% changed)
Mobility for residential areas (% changed)
Month
Mar
Apr
May
Jun
Jul
Aug

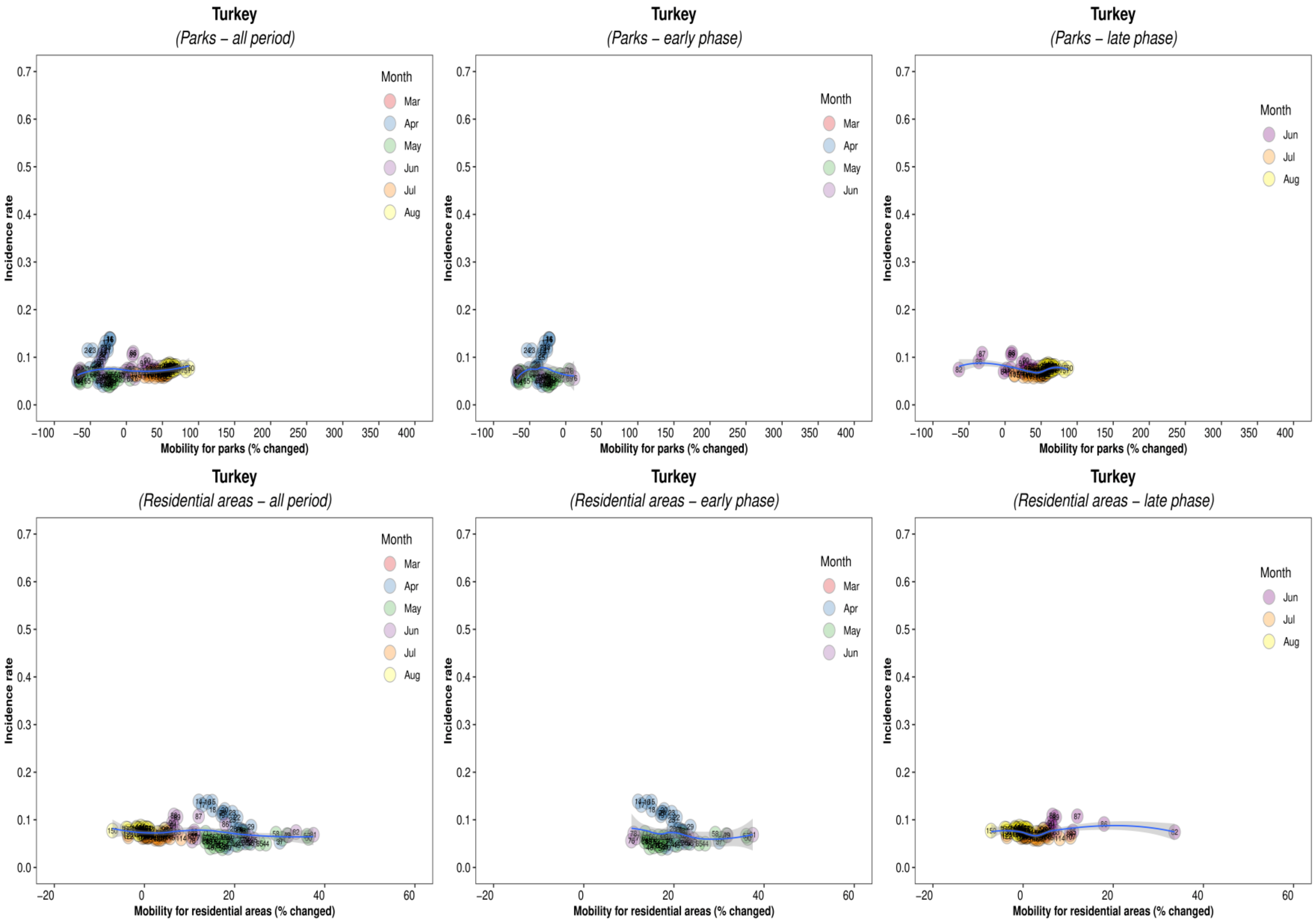
Turkey
(Parks – all period)
Turkey
(Parks – early phase)
Turkey
(Parks – late phase)
Turkey
(Residential areas – all period)
Turkey
(Residential areas – early phase)
Turkey
(Residential areas – late phase)
Incidence rate
Mobility for parks (% changed)
Mobility for residential areas (% changed)
Month
Mar
Apr
May
Jun
Jul
Aug

Supplementary Figure 3. Association between new daily incidence rates of COVID-19 and mobility changes in 36 countries, for grocery and pharmacy visits.

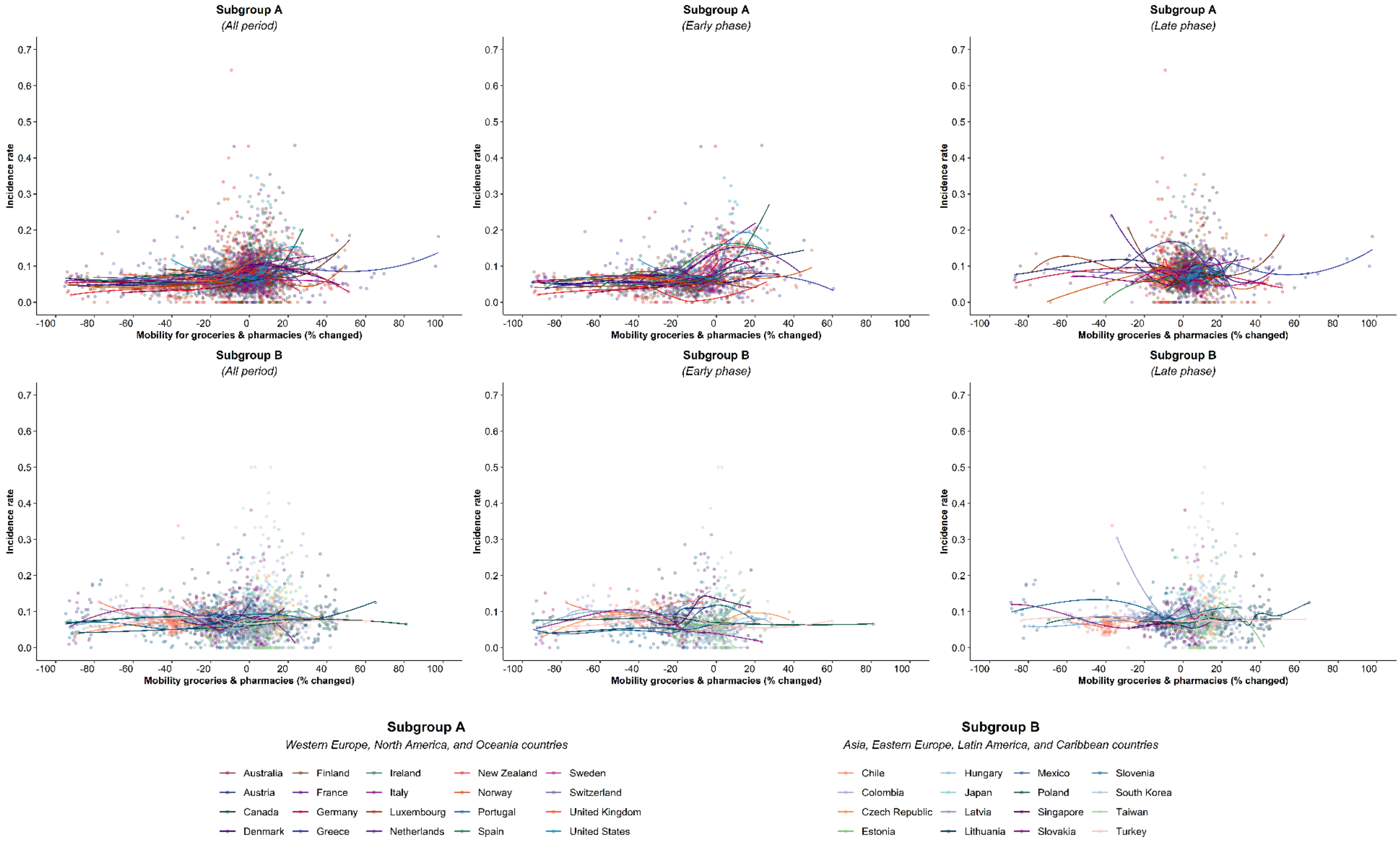


Footnote 1. The mobility change measurement period was from the day of the 100th case in each country through August 31, 2020.
Footnote 2. Pandemic phase was defined for each country by the median of the date when the 100$^{th}$ case was detected to the end of the study period: early phase for the period before the median date and late phase for the period after the median date.

Supplementary Figure 4. Association between new daily incidence rates of COVID-19 and mobility changes for 36 countries based on alternative lag days

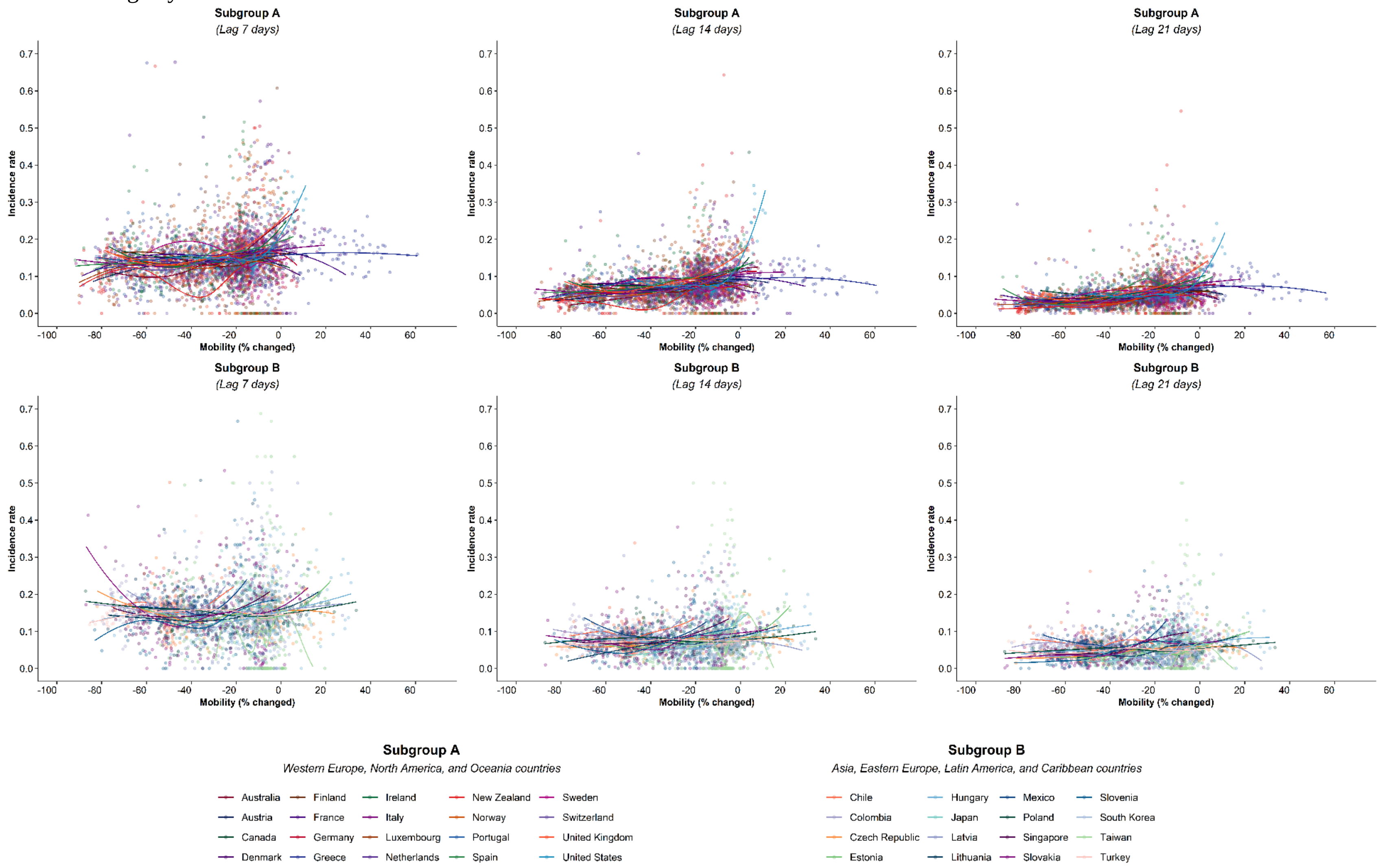


Footnote 1. The mobility change measurement period was from the day of the 100th case in each country through August 31, 2020.
